# Distinct Roles of Therapeutic Expectations and Dissociative Symptoms in Antidepressant Response During Ketamine Treatment in Routine Care

**DOI:** 10.64898/2026.04.23.26351276

**Authors:** Willys Cantenys, Zeynep Yoldas, Luc Masset, Alix Romier, Léonie Samion, Marion Imbault, Thai-Moc Tram Tran, Flavia Megda Garcia, Samar Soltani, Martina Marradi, Hanh-Dan Ho, Antoinette Gohier, Chahinaz Doulazmi, Marianne Chesneau, Cécile Jadaan, Joy Bulut, Ndilyam Djonouma, Salma Charradi, Anne Claret Tournier, Philippe Fossati, Liane Schmidt

**Author notes:** Corresponding Author: Liane Schmidt, PhD, Address: 47 boulevard de l’Hôpital, 75013 Paris, France.

## Abstract

**Background:** Ketamine is a rapid-acting antidepressant that produces acute dissociative symptoms. In routine care, the respective contributions of therapeutic expectations and dissociative symptoms to antidepressant response, and the directionality of their associations with depressive symptom change, remain poorly characterized.

**Methods:** We conducted a retrospective longitudinal observational cohort study of 100 adults with major depressive disorder or bipolar depression receiving six open-label intravenous racemic ketamine infusions over 3 weeks. Therapeutic expectations were rated at baseline and before each infusion. Post-infusion, dissociative symptoms (CADSS) were assessed first, followed by depressive symptom severity (MADRS) within the same session. Linear mixed-effects, mediation, and random intercept cross-lagged panel models (RI-CLPM) were used to distinguish within-person from between-person effects.

**Results:** Depressive symptoms improved across the induction course, with 45% of participants meeting response criteria. At each session, therapeutic expectations consistently predicted post-infusion improvement in depressive symptoms at the within-person level, independently of dissociative symptoms. Moreover, expectations became stronger across sessions in treatment responders. Dissociative symptoms were associated with improvement when examined alone, but were not observed after adjustment for expectations, and were not linked to improvement in depression at the within-person level from session to session. They were associated with greater overall antidepressant benefit at the between-person level. A notable indirect pathway was identified at the between-person level, where expectations determined changes in depression through initial dissociative symptoms and early depressive symptom reductions. This pathway explained 3.2% of the total effect of expectations on improvement in depression by the end of treatment.

**Conclusions:** Therapeutic expectations and dissociative symptoms contributed to antidepressant response through distinct pathways: expectations functioned at the individual level as a dynamic within-person driver, whereas dissociative propensity served on the group level as a stable between-person marker of outcome, highlighting complementary clinical targets to optimize treatment response in routine care.

## Introduction

Major depressive disorder affects about 330 million people worldwide and remains a leading cause of years lived with disability, contributing to premature mortality and overall disease burden (1). Despite available monoaminergic antidepressants, up to one-third of patients do not achieve adequate remission, highlighting the need to better understand the factors that shape treatment response (2).

Antidepressant effectiveness depends not only on pharmacological mechanisms (efficacy) but also on psychological and contextual factors that influence how treatment is perceived and evaluated (3). Among these factors, patients’ therapeutic expectations—beliefs about treatment effectiveness and perceived improvement—are associated with clinical improvement across pharmacologic, psychotherapeutic, and somatic treatments (4–8). This association may be part of a feedback loop in which early symptom changes shape patients’ beliefs about treatment effectiveness, which, in turn, determine adherence and responsiveness to subsequent medical treatments and interventions (9). The growing recognition of the role of therapeutic expectations in antidepressant clinical trials (3,10) underscores the need to better understand how expectations are formed and change over the course of treatment.

These questions are particularly relevant for novel, fast-acting antidepressants such as ketamine, an N-methyl-D-aspartate (NMDA) receptor antagonist. Subanesthetic doses of ketamine have rapid antidepressant effects in treatment-resistant depression (11,12) that may be highly sensitive to expectation (13,14). Moreover, beyond its molecular effects, ketamine induces transient alterations in perception and sense of self, manifesting as changes in bodily sensations, distortions of spatial or temporal perception, and feelings of detachment from oneself or the external world, typically described as dissociative symptoms (15,16).

Although these dissociative symptoms are well described, their importance for antidepressant treatment response remains debated (14,17–23). While some studies have reported associations between dissociative symptom levels and antidepressant response (18–20), other investigations have not found consistent evidence for this relationship (21–23). Interventional studies further suggest that when dissociative symptoms are masked by full anesthesia or when psychoactive placebos provide other active-treatment cues, ketamine’s antidepressant advantages over control are diminished or absent (24,25).

However, the respective contributions of therapeutic expectations and dissociative symptoms to antidepressant response in routine care, as well as the interaction and directionality of their associations with changes in depressive symptoms across repeated infusions, remain poorly characterized. It remains unclear whether dissociative symptoms directly predict antidepressant response or whether they mediate the association of therapeutic expectations with depressive symptoms.

When patients undergo antidepressant treatment, they are given information about the treatment from clinicians and patient leaflets (4,26). Through this information, they form therapeutic expectations that can influence how they experience dissociative symptoms (14), suggesting overall that these two factors do not operate independently but interact to determine antidepressant response.

We hypothesized here that, during ketamine treatment, therapeutic expectations may shape the intensity of ketamine-induced dissociative symptoms, which act as early cues of pharmacologic activity and contribute indirectly to clinical improvement (14,27). To characterize the directionality and magnitude of these associations, we examined (i) whether pre-infusion therapeutic expectations predict the intensity of ketamine-induced dissociative symptoms and subsequent antidepressant response, (ii) whether dissociative symptoms predict post-infusion antidepressant response, and (iii) whether these associations reflect dynamic processes unfolding within individual patients across infusions or stable differences between patients.

To test these hypotheses, we retrospectively analyzed routinely collected clinical data on therapeutic expectations, dissociative symptoms, and depressive symptoms across a 6-infusion course of racemic ketamine in patients with moderate to severe depression, using a multilevel analytical strategy designed to characterize both the overall structure and the within-person directionality of these associations across the induction course.

## Methods

### Ethical considerations

The study protocol was approved by the Sorbonne University ethics committee (approval number CER-2025-CANTENYS-KP-00431) and conducted in accordance with the Declaration of Helsinki. The committee approved secondary use of deidentified routine clinical data with an opt-out procedure; patients were informed and could decline use of their data.

### Participants

This retrospective observational study included data from 100 patients with moderate to severe depression who received ketamine antidepressant treatment between June 2021 and July 2025. All patients were treated in the Department of Adult Psychiatry at Pitié-Salpêtrière Hospital. Data were deidentified and extracted from electronic medical records by the clinical team using secure internal systems. Eligible patients had a DSM-5–based clinical diagnosis of major depressive disorder or bipolar depression, received at least one intravenous ketamine infusion, and had available baseline and at least one post-infusion assessment. Among the 100 patients, 37 were men, and 63 were women, with a mean age of 45.7 years (range, 19–77 years). Patients were diagnosed with either major depressive disorder (n = 80) or bipolar depression (n = 20). Baseline clinical characteristics, comorbidities, and concomitant treatments are summarized in Table 1.

**Table 1.**
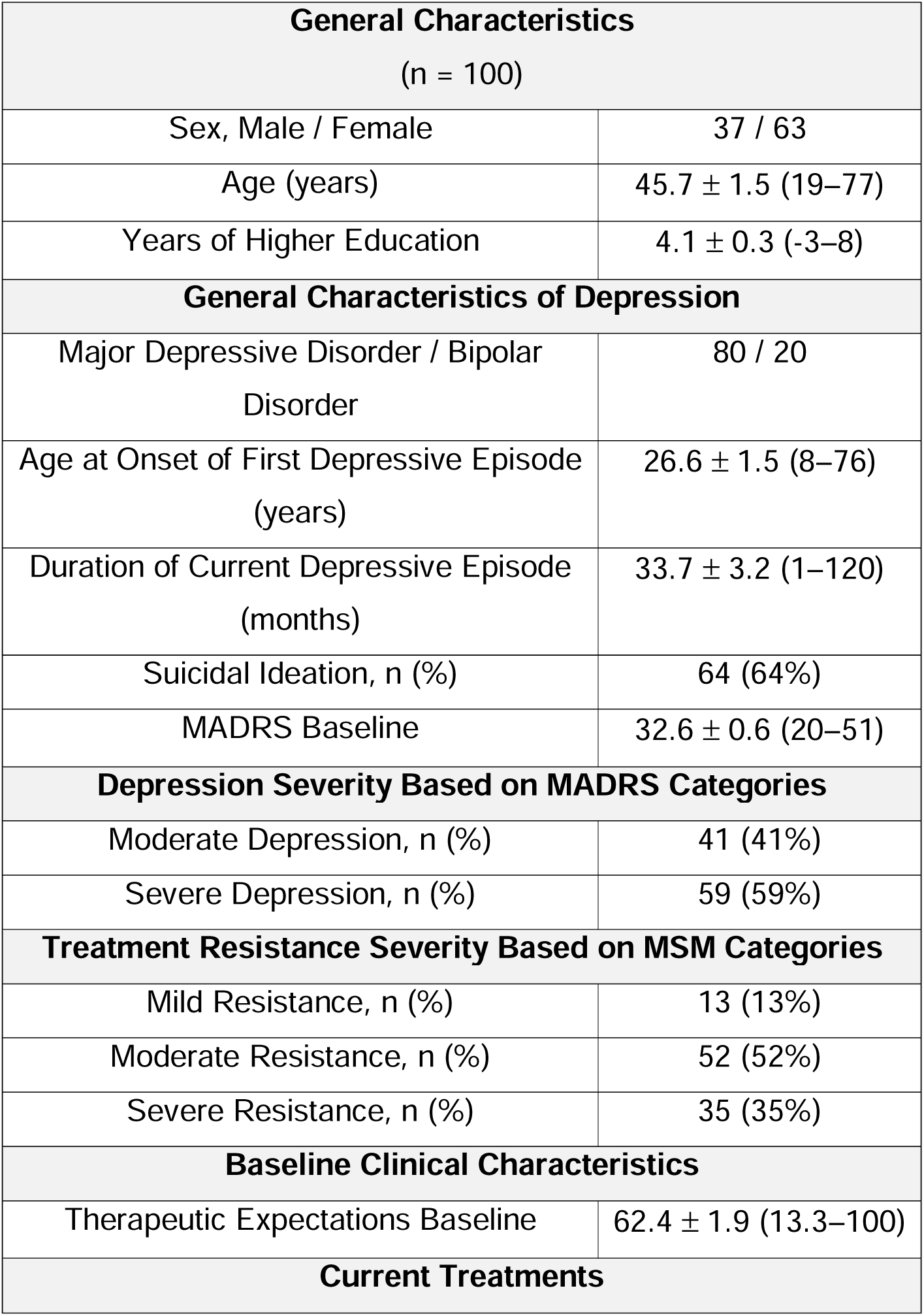
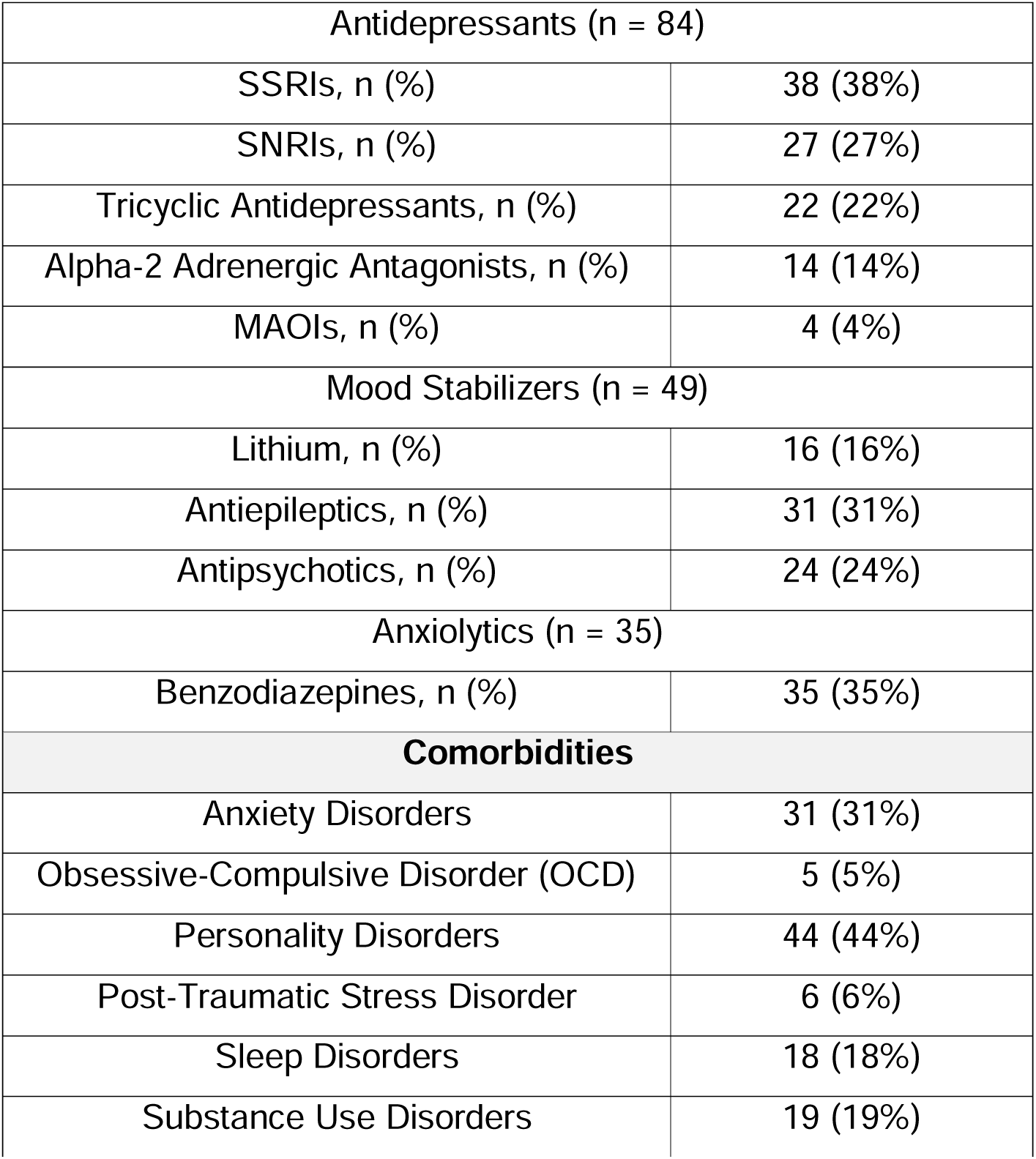
Sociodemographic and clinical data. **Note**. The duration of the current depressive episode was quantified in months. Age at illness onset was defined as the patient’s age at first clinically diagnosed depressive episode. Educational level was operationalized as years relative to completion of secondary education (reported as years of higher education; 0 = high school completion; negative values indicate lower educational attainment). Disease severity was categorized according to Montgomery-Åsberg Depression Rating Scale (MADRS) thresholds: moderate severity, scores 20–31; severe severity, scores 32–60. Treatment resistance was stratified using the Maudsley Staging Method (MSM), with mild resistance defined as scores of 3 to 6, moderate resistance as scores of 7 to 10, and severe resistance as scores of 11 to 15. Data are presented as mean ± SE (range). MADRS indicates Montgomery-Åsberg Depression Rating Scale; MSM, Maudsley Staging Method; SSRI, selective serotonin reuptake inhibitor; SNRI, serotonin and norepinephrine reuptake inhibitor; MAOI, monoamine oxidase inhibitor.

### Study design

Longitudinal data were retrospectively extracted from standardized clinical assessments focusing on dissociative symptoms and therapeutic expectations, measured repeatedly throughout the treatment course in routine care. The program was delivered as routine clinical care, not for research purposes. Patients received 6 open-label intravenous racemic ketamine infusions over 3 weeks, with infusions scheduled twice weekly. Dosages were individualized through stepwise titration (0.5–1.0 mg/kg) based on clinical response and tolerability. Analyses focused on the initial 6 infusions because all patients are offered this induction phase in routine care. Continuation beyond this period depends on clinical response and may introduce selection bias; analyses were therefore restricted to the induction phase. Details of the administration protocol are provided in Methods S1 and Figure S1 in the Supplement (28).

### Data quality

Clinical data were abstracted retrospectively from medical records and entered into a dedicated study database by the clinical team using standardized extraction procedures. Depressive symptom severity (MADRS) was clinician-rated by trained psychiatrists or supervised residents as part of routine care. Ratings were obtained at each assessment time point and verified against source documentation. Missing assessments were recorded along with the reason for missingness (eg, scheduling constraints or early discontinuation). Longitudinal analyses used linear mixed-effects models estimated by maximum likelihood, which accommodate incomplete repeated-measures data under a missing-at-random assumption conditional on observed data. Missing data handling and diagnostics are detailed in Methods S2 and Figure S2 in the Supplement.

### Outcome Measures

The Montgomery-Åsberg Depression Rating Scale (MADRS) was used to assess depressive symptom severity (29), given its sensitivity to rapid symptom changes over short intervals (30).

Dissociative symptoms during ketamine treatment were assessed using the 23-item Clinician-Administered Dissociative States Scale (CADSS; 0–4 per item) (31). Higher scores indicate greater intensity of dissociative symptoms. Total CADSS scores and depersonalization, derealization, and amnesia subscales were examined as measures of acute dissociative symptoms during infusions (32) (Methods S3).

Therapeutic expectations were assessed using a 3-item patient-reported composite score (Methods S4; Figure S3). Items were rated on a 0 to 100 visual analog scale and captured perceived effectiveness of ketamine for depression, expected recovery, and expected posttreatment depression severity (reverse scored). The expectation score was the mean of the 3 items, with higher scores indicating greater therapeutic expectations.

In this study, *therapeutic expectations* refer to patients’ anticipated response and prospective beliefs about treatment effectiveness in routine care (i.e., response expectations) (10). Therapeutic expectations do not involve expected *treatment efficacy*, which refers to antidepressant effects observed under ideal randomized clinical trial conditions (33).

### Statistical Analysis

Statistical analyses were performed using MATLAB 2024b (Statistics and Machine Learning Toolbox, the CANlab Mediation Toolbox) (34,35). Analyses followed a sequential, multilevel strategy: longitudinal and session-level models first characterized population-level associations, mediation analyses then formalized their directional structure at the between-person level, and random intercept cross-lagged panel models finally decomposed these associations into within-person dynamics and stable between-person differences.

### Population-level and session-level analyses

Longitudinal, session-level, and categorical models were used to examine the associations between expectations and dissociative symptoms with MADRS scores over the 3-week course. Dissociative symptoms were modeled using total CADSS and depersonalization, derealization, and amnesia subscales. Dose sensitivity analyses were restricted to 0.5 mg/kg sessions (Methods S5). Baseline correlations and collinearity checks were assessed at T0 (Methods S6). Detailed specifications of these longitudinal, session-level, and categorical models are provided (Methods S7 in the Supplement).

#### (1) Longitudinal analysis of depressive symptom trajectories

First, longitudinal associations between therapeutic expectations, dissociative symptoms, and MADRS scores were assessed across infusions 1–6 using linear mixed-effects models (LMEs). LME1 included fixed effects for time, expectations, CADSS, and their 2 and 3-way interactions, adjusted for age, education, and sex, with random intercepts and random slopes for time. LME2 and LME3 were reduced versions of LME1, including expectations (LME2) or dissociative symptoms (LME3), respectively, and their interactions with time, using the same covariates and random-effects structure. Full model equations and diagnostics are provided in Methods S8–S9 and Figures S4–S6. All variables were standardized (z scores) using the overall mean and standard deviation across all observations. Models were estimated using maximum likelihood estimation (MLE).

#### (2) Session-level associations of therapeutic expectations and dissociative symptoms

Session-level analyses examined whether therapeutic expectations or dissociative symptoms measured at each time point were associated with end-of-course improvement (T0–T6). End-of-course improvement was defined as the percent change in MADRS from baseline (T0) to each patient’s last MADRS assessment during the induction phase (T_Endpoint_). Thirteen separate linear regression models were fit—7 for expectations and 6 for dissociative symptoms—to estimate the change in percent improvement per standard deviation increase in the predictor variable, adjusting for age, education, and sex. Statistical significance was assessed using Bonferroni correction for 13 comparisons; models including CADSS subscales are described in Methods S5.

#### (3) Categorical analysis of therapeutic expectations and dissociative symptoms across response groups

For categorical analyses, patients were classified as low (<25% reduction in MADRS), moderate (25%–75%), or high (≥75%) responders (Figure S7). Two LMEs (LME4 and LME5) modeled expectations and CADSS scores, respectively, as outcomes. Fixed effects were included for time (i.e., number of infusions), response group, and their interaction, and random intercepts and random slopes for time, adjusting for age, sex, and education. Models were estimated using restricted maximum likelihood (REML) in MATLAB (fitlme) (Tables S6–7, Methods S10, Figures S8–S9). These analyses characterized overall and session-level associations but did not formally test their directional structure, which was addressed in subsequent mediation analyses.

### Multivariate Effects: Mediation Analyses

To examine how therapeutic expectations are formally linked to end-of-course improvement, a 3-path mediation analysis was conducted (36). The model tested the hypothesis that baseline therapeutic expectations (x) statistically predict the intensity of dissociative symptoms during the first ketamine infusion (m1), which then determine early positive shifts in depressive symptoms at the start of the treatment (m2), and translate into overall end-of-course improvement in depression.

The model was specified as follows:

(1) Path a1: the univariate effect of baseline therapeutic expectations (x) on dissociative symptoms (m1).

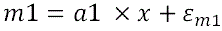
(2) Path a2: the univariate effect of dissociative symptoms during the first ketamine infusion (m1) on depressive symptoms after the first ketamine infusion (m2), controlling for the impact of expectations (x) and the direct effect (d) of expectations on such early clinical improvement.

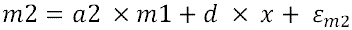
(3) Path b: the effect of early clinical improvement (m2) on end-of-course improvement (y), controlling for the direct effect (c’) of expectations on end-of-course improvement and the direct effect (f) of dissociative symptoms during the first infusion (m1) on end-of-course improvement (y).

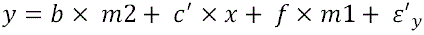

Single-level mediation, the indirect effect, was assessed by:

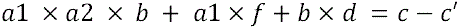

The serial regressions were controlled for individual differences in age, sex, and level of education among patients. Because early and end-of-course improvement were defined relative to the baseline MADRS score, the model also controlled for variability in baseline depression severity across patients. All variables (x, m1, m2, y, covariates) were standardized into z-scores. All path regressions were tested for significance using nonparametric bootstrap resampling (10,000 samples, with replacement), and 2-sided P values were derived from bootstrapped confidence intervals. The magnitude of the indirect effect was additionally quantified using κ² (kappa-squared), which expresses the indirect effect as a proportion of its structural maximum, independently of scale and model complexity; conventional thresholds define small (κ*²* ≥ 0.01), medium (κ*²* ≥ 0.09), and large (κ*²* ≥ 0.25) effects (37).

### Multivariate Cross-Lagged Analyses: Within-Person Dynamics and Between-Person Differences

Mediation analyses, operating at the between-person level and anchored to the first infusion, do not capture how the directional associations between therapeutic expectations, dissociative symptoms, and depressive symptoms unfold across the six infusion sessions within individual patients. Random Intercept Cross-Lagged Panel Models (RI-CLPM; (38)) were therefore estimated to characterize these directional associations, separating within-person effects, which reflected session-to-session deviations from each patient’s own average, from between-person effects, which reflected the overall differences between patients captured by the random intercepts.

#### Model specification

Three complementary RI-CLPMs were estimated: a trivariate model (RI-CLPM1) including therapeutic expectations, dissociative symptoms, and depressive symptoms, and two bivariate models (RI-CLPM2: expectations and depressive symptoms; RI-CLPM3: dissociative and depressive symptoms) used as robustness checks (see Methods S11).

RI-CLPM1 tested whether therapeutic expectations predict both dissociative symptoms (β*_ec_*) and depressive symptom severity (β*_ed_*) within each session, and whether dissociative symptoms additionally predict depressive symptom severity (β*_cd_*) beyond expectations, while controlling for reverse temporal paths (β*_de_,* β*_ce_,* β*_dc_*) following:

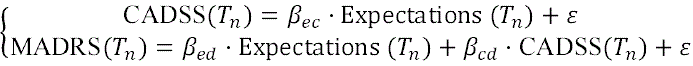

Within-session directional paths (lag0) are labeled by source and target variable initials, e (Expectations), c (CADSS), d (MADRS), such that β*_ec_*, β*_ed_*, and β*_cd_* denote Expectations→CADSS, Expectations→MADRS, and CADSS→MADRS, respectively. The assessment sequence was fixed: therapeutic expectations were assessed before infusion; post-infusion, dissociative symptoms were systematically assessed by the patient (CADSS) prior to depressive symptom severity rated by the clinician (MADRS), within a 1–4 hour post-infusion window once acute dissociative symptoms had clinically dissipated. This fixed sequential ordering renders contemporaneous directional paths appropriate rather than bidirectional covariances (39,40). Inter-session reverse paths follow the same convention, β*_de_* (MADRS→Expectations), β*_ce_* (CADSS→Expectations), and β*_dc_* (MADRS→CADSS), reflecting associations from session T_n_ to session T_n+1_, and were included to evaluate the directionality of cross-lagged effects.

Age, sex, and education were regressed on all random intercepts; baseline MADRS was additionally regressed on the random intercept of depressive symptoms (RId) to account for stable between-person differences in baseline depressive symptom severity.

#### Model estimation

Model parameters were estimated using maximum likelihood with full-information maximum likelihood for missing data and robust standard errors (MLR estimator, Yuan-Bentler correction) in R (lavaan 0.6-21; (41)).

The estimation of within-session path parameters (β*_ed_,* β*_ec,_*β*_de_, and* β*_ce_*) was unconstrained at each session T1–T6. Inter-session autoregressive paths were constrained to equality across transitions, consistent with the assumption of temporal stationarity in within-person dynamics (38). The effect of depressive symptoms on dissociative symptoms quantified by the parameter β*_dc_* was also constrained to equality, as ketamine-induced dissociative symptom intensity is primarily determined by acute pharmacological exposure rather than prior depressive symptom severity (17) (see Methods S11 and Tables S10–S12 for full specification and model results). All cross-lagged paths are estimated on the within-person component (42). Model fit was acceptable for the trivariate RI-CLPM1 (CFI = 0.91, RMSEA = 0.08) (43,44).

#### Parameter comparisons

Directional hypotheses were tested using the Generalized Order-Restricted Information Criterion Approximation (GORICA (45); restriktor v0.6-30), an information-criterion approach that quantifies relative support for order-restricted hypotheses under structural equation models. Notably, we tested whether patients who expected greater therapeutic benefit experienced better outcomes, or whether those who showed greater depressive improvement formed stronger therapeutic expectations, by comparing |β*_ed_*| > |β*_de_*|.

Likewise, we tested whether those who expected more therapeutic benefit displayed stronger dissociative symptoms, or whether stronger dissociative symptoms led to higher therapeutic expectations, by comparing |β*_ec_*| > |β*_ce_*|.

For each hypothesis, the GORICA weight (*w*) represents the relative support for the hypothesized order constraint compared with its complement (range, 0–1), and can be interpreted as the relative weight of evidence favoring that hypothesis within the candidate set. The associated evidence ratio (w[H]/w[complement]) quantifies the strength of support, with values greater than 1 indicating stronger support for the hypothesis.

## Results

### Longitudinal associations of therapeutic expectations and dissociative symptoms

As shown in Figure 2A, MADRS scores, mean (SE), decreased from 32.6 (0.55) at baseline (T0) to 24.4 (0.76) after the first infusion (T1), 19.6 (0.86) after the third infusion (T3), and 17.1 (0.97) after the sixth infusion (T6). This decrease was supported by a significant fixed effect of time in the longitudinal mixed-effects models (e.g., LME1: β = –0.27; *SE* = 0.05; *P* < .001; *95% CI*, –0.36 to –0.18; Table S1). After the sixth infusion, 45% of participants (n = 45) met response criteria (≥ 50% reduction in MADRS), including 12 patients (12%) who achieved remission (MADRS < 7).

**Figure 1.**
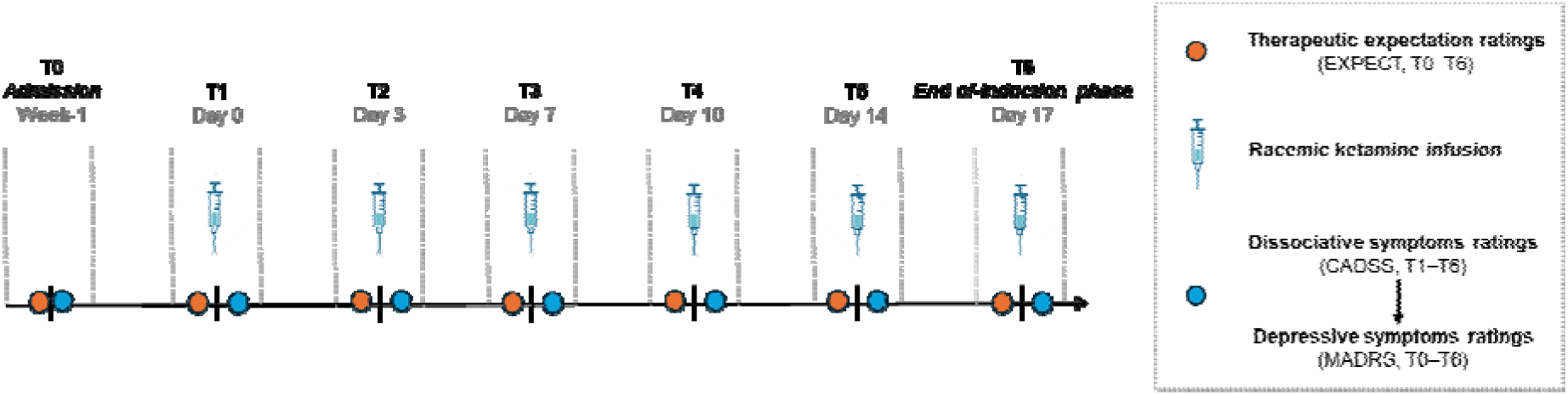
Study design. Six open-label intravenous racemic ketamine infusions (T1–T6) were administered twice weekly for three weeks. Therapeutic expectations were rated by patients at baseline (prior to the first infusion, T0) and immediately before each infusion. Post-infusion, dissociative symptoms were first assessed using the Clinician-Administered Dissociative States Scale (CADSS), followed by depression symptom severity assessed using the Montgomery–Åsberg Depression Rating Scale (MADRS) by a trained psychiatrist, within the same session: dissociative symptoms were assessed before depressive symptom severity, both within a 1-4 hour post-infusion window once acute dissociative symptoms had clinically dissipated. T0 corresponded to the pretreatment baseline assessment completed within the 2 weeks preceding the first infusion; T1–T6 corresponded to post-infusion assessments following infusions 1–6; T_Endpoint_ corresponded to the last available post-session MADRS assessment during the 3-week induction phase. Dosage was adjusted, when clinically indicated, from 0.5 to 0.75 mg/kg and, if needed, to 1.0 mg/kg in 55 of 100 patients.

**Figure 2.**
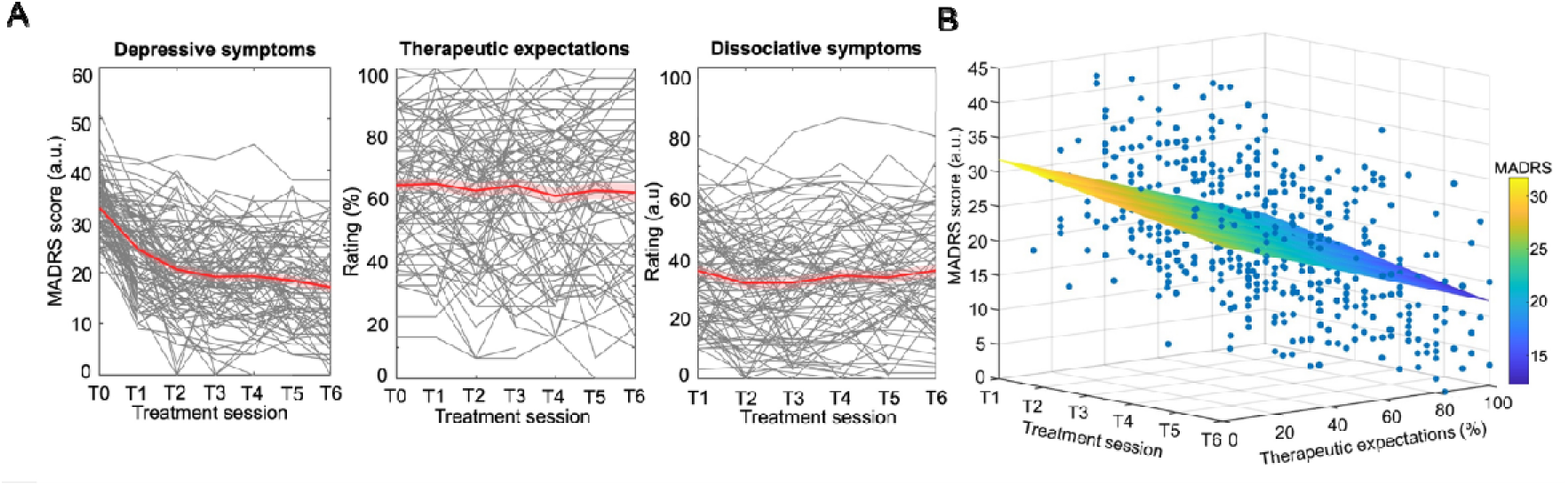
Longitudinal trajectories of depressive symptoms, therapeutic expectations, and dissociative symptoms during ketamine induction. **(A)** Line plots show the trajectories of depressive symptom severity (MADRS total score, arbitrary units [a.u.]), therapeutic expectations (0–100), and dissociative symptoms (CADSS total score, a.u.) from admission (T0) through six subsequent racemic ketamine infusions (T1–T6). Individual lines in gray represent individual patients, and the red, bold line represents the mean, with error bars indicating standard deviations. **(B)** The 3-dimensional mesh illustrates the main effects of treatment over time, as well as the main effect of therapeutic expectations on depression severity. The x-axis indicates the treatment time point (from first to sixth racemic ketamine infusion), the y-axis represents therapeutic expectations, and the z-axis depicts depression severity measured by MADRS scores. Data points in blue denote individual patients at each treatment and testing time point. The surface color mapping corresponds to MADRS scores on the z-axis.

Therapeutic expectations were associated with overall lower MADRS scores (LME1: β = –0.34; *SE* = 0.05; *P* < .001; *95% CI*, –0.43 to –0.24; Table S1), including in models adjusting for dissociative symptom intensity (LME2: β = –0.32; *SE* = 0.05; *P* < .001; *95% CI*, –0.42 to –0.23; Table S2). Interactions with time and dissociative symptoms were not significant (Table S1).

Dissociative symptoms were also associated with lower MADRS scores in models without expectations (LME3: β = –0.11; *SE* = 0.05; *P* = .03; Table S3), but not after adjustment for expectations (LME1: β = –0.06; *SE* = 0.05; *P* = .20; *95% CI*, –0.15 to 0.03; Table S1). In subscale analyses, depersonalization was associated with lower MADRS scores (LME3_depersonalization_: β = –0.13; *SE* = 0.05; *P* = .005; *95% CI*, –0.22 to – 0.04; Table S3), whereas derealization showed a trend (LME3_derealization_: β = –0.08; *SE* = 0.05; *P* = .10; *95% CI*, –0.17 to 0.02; Table S3) and became significant in fixed-dose analyses (LME3_derealization_: β = –0.12; *SE* = 0.05; *P* = .02; *95% CI*, –0.23 to –0.02; Table S15). Amnesia was not associated with MADRS scores (LME3_amnesia_: β = – 0.02; *SE* = 0.04; *P* = .68; *95% CI*, –0.11 to 0.07; Table S3); other fixed-dose results were similar (Tables S13–15).

### Session-specific associations of therapeutic expectations and dissociative symptoms

Therapeutic expectations were significantly associated with greater end-of-course improvement in depression at each treatment session (β range, 9.01–15.86; all *P* < .0038, Bonferroni-corrected; Figure 3A and Table S4). Dissociative symptoms were also associated with end-of-course improvement at the first (β = 8.68; *P* = .002, Bonferroni-corrected) and third infusion (β = 9.94; *P* < .001; Bonferroni-corrected), and to a lesser extent at the other time points (β range, 6.09–8.02; *P* < .05, not adjusted for multiple comparisons). These results indicate that a one-standard deviation increase in expectations was associated with approximately 20% to 35% greater end-of-course improvement in MADRS, compared with 13% to 23% for dissociative symptoms.

**Figure 3.**
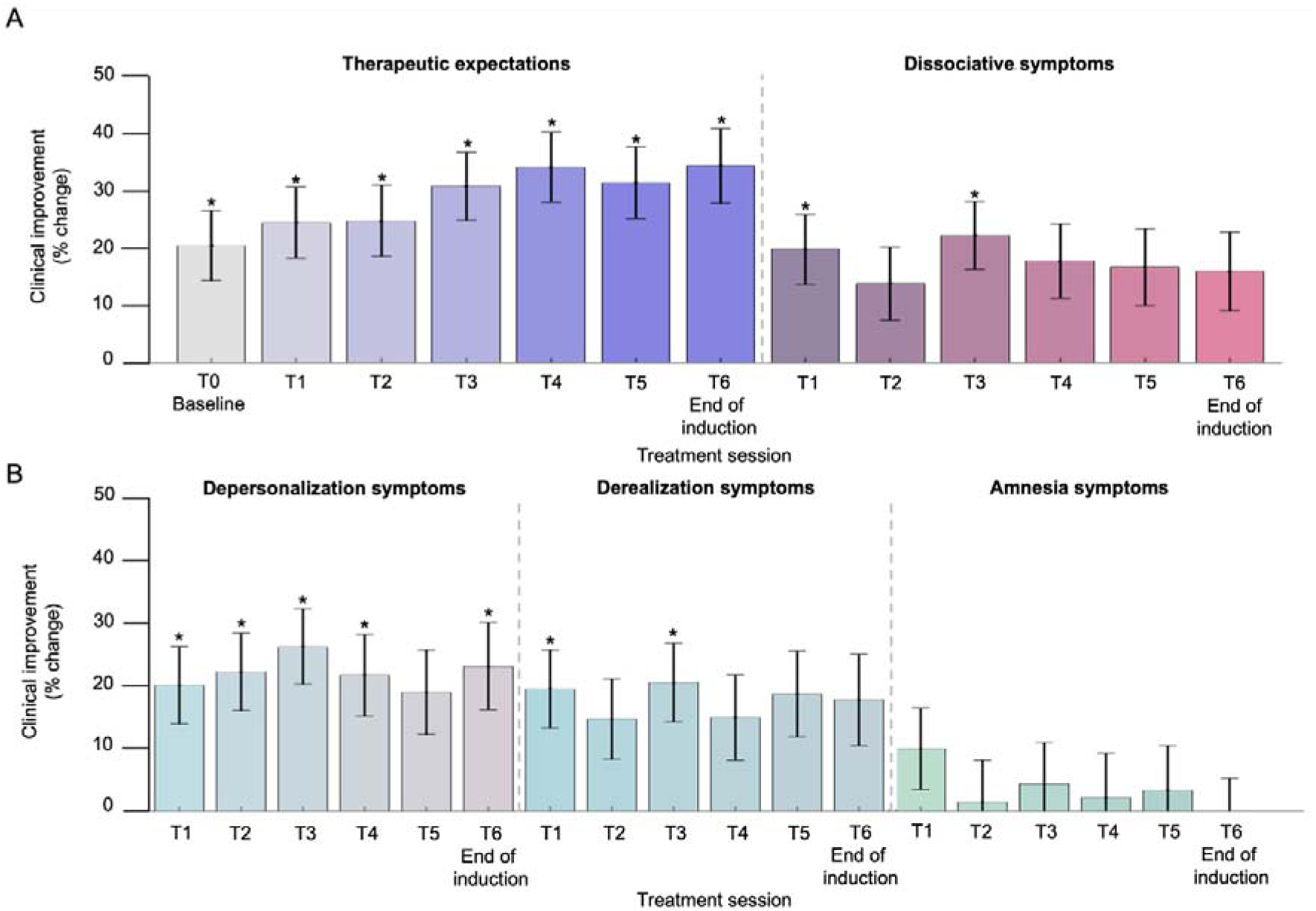
Session-specific effects of therapeutic expectations and dissociative symptoms. **(A)** Percent change in depressive symptoms, measured by the Montgomery-Åsberg Depression Rating Scale (MADRS), per standard deviation increase in patient-reported therapeutic expectations or dissociative symptoms at each testing session (T0–T6; n = 100; 624 observations). **(B)** Percent change in depressive symptoms per standard deviation increase in dissociative symptom dimensions, depersonalization, derealization, and amnesia assessed following an infusion (T1–T6, n = 100; 524 observations). Error bars indicate standard errors (*SEs*). Asterisks indicate significance after Bonferroni correction (\**P < .0038 in **A**; *P < .0028 in **B**)*.

Similar results were observed for depersonalization and derealization, although their temporal patterns differed. Depersonalization symptoms were significantly associated with greater depressive improvement from the first through the sixth infusion, except at the fifth session (β range, 8.60–11.44; *P* < .0028, Bonferroni-corrected; Table S5). Derealization symptoms were associated with improvement at the first and third sessions only (β range, 8.32–8.95; *P* < .0028, Bonferroni-corrected; Table S5). Amnesia was not significantly associated with end-of-course improvement (*P* = .13–.83, not adjusted for multiple comparisons) (Figure 3B and Tables S4 and S5 in the Supplement). Results were similar in fixed-dose analyses (Tables S16 and S17).

### Categorical associations of treatment responsiveness with therapeutic expectations and dissociative symptoms

Therapeutic expectations and dissociative symptoms differed across response groups, with moderate (25%–75%) and high (≥75%) responders showing higher scores than low responders (expectations: β = 0.65; *SE* = 0.12; *P* < .001; *95% CI*, 0.41 to 0.89; dissociation: β = 0.48; *SE* = 0.13; *P* < .001; *95% CI*, 0.22 to 0.75; Tables S6 and S7; Figure 4A–C). A significant group-by-time interaction for expectations (β = 0.19; *SE* = 0.06; *P* = .001; *95% CI*, 0.07 to 0.30) indicated increasing expectations with subsequent infusions among high responders (Figure 4B). Significant associations with response group were observed for depersonalization (β = 0.57; *SE* = 0.13; *P* < .001; *95% CI*, 0.33 to 0.82; Table S7) and derealization (β = 0.40; *SE* = 0.13; *P* = .003; *95% CI*, 0.14 to 0.67; Table S7), but not for amnesia (β = 0.13; *SE* = 0.14; *P* = .32; *95% CI*, –0.13 to 0.40; Table S7). Findings were maintained in fixed-dose analyses (Tables S18 and S19).

**Figure 4.**
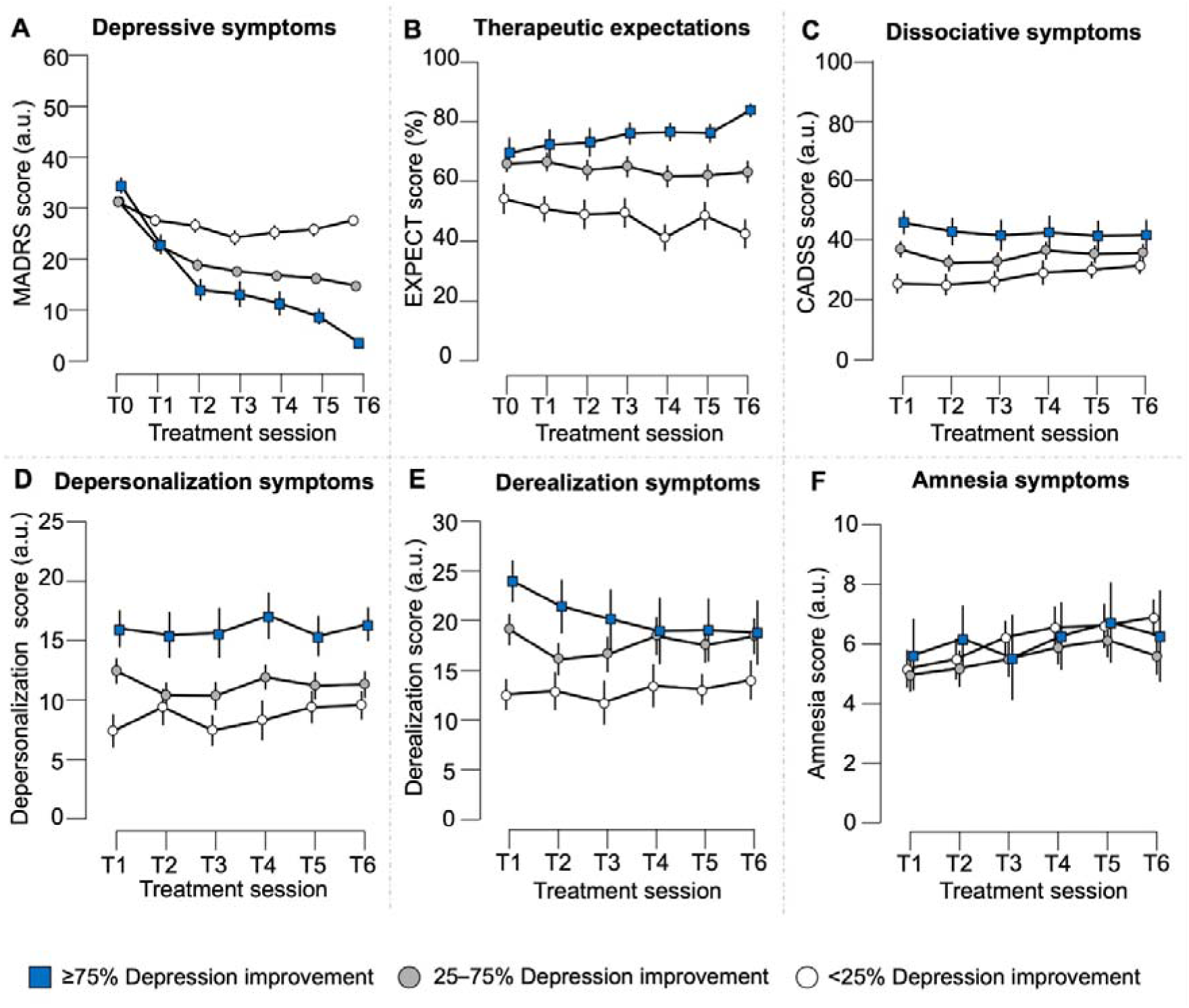
Trajectories of depression, expectations, and dissociation by end-of-induction response group. Mean depression severity (MADRS score; panel A), therapeutic expectations (panel B), and dissociative symptoms (total CADSS score; panel C) and CADSS dimensions—depersonalization, derealization, and amnesia (panels D–F)—are shown at each time point during the 6-infusion induction phase, stratified by end-of-induction clinical response group (low responders: <25%, moderate responders: 25–75%, high responders: ≥75% end-of-induction MADRS improvement; n = 100). Error bars indicate within-group standard errors (*SEs*). MADRS, Montgomery-Åsberg Depression Rating Scale; CADSS, Clinician-Administered Dissociative States Scale; Therapeutic expectation ratings (0–100); a.u., arbitrary scale units.

### Multivariate effects: Mediation analyses

A 3-path mediation analysis tested whether dissociative symptoms and early improvement sequentially mediated the association between therapeutic expectations and end-of-course improvement (path c total effect: β = 0.39; *SE* = 0.09; *P* < .001; Table S8). Higher expectations were associated with greater dissociative symptoms during the first infusion (path a1: β = 0.20; *SE* = 0.09; *P* = .040), greater dissociative symptoms were associated with greater early improvement (path a2: β = 0.19; *SE* = 0.10; *P* = .050), and greater early improvement was associated with greater end-of-course improvement (path b: β = 0.33; *SE* = 0.10; *P* = .003). The direct association between expectations and end-of-course improvement remained significant after accounting for mediators (direct path c′: β = 0.24; *SE* = 0.11; *P* = .022), indicating partial mediation. The indirect 3-path effect was statistically significant (indirect path a1a2b: β = 0.01; *SE* = 0.01; *P* = .037; 3.2% of total effect), with κ*²* = 0.34, reflecting a high proportion of the attainable maximum for this mediation structure. Among dissociative dimensions, only derealization showed a significant indirect effect (*P* = .048), while depersonalization and amnesia did not (Table S9). These findings were replicated in analyses restricted to 0.5 mg/kg sessions, with a significant indirect effect for the overall CADSS score (*P* = .045) and a trend toward significance for derealization (*P* = .069; Tables S20 and S21).

### Multivariate Cross-Lagged Analyses: Within-Person Dynamics and Between-Person Differences

The trivariate RI-CLPM (Figure 6A) disentangled session-by-session directional dynamics among therapeutic expectations, dissociative symptoms, and depressive symptoms from inter-individual differences sustained across the treatment course.

**Figure 5.**
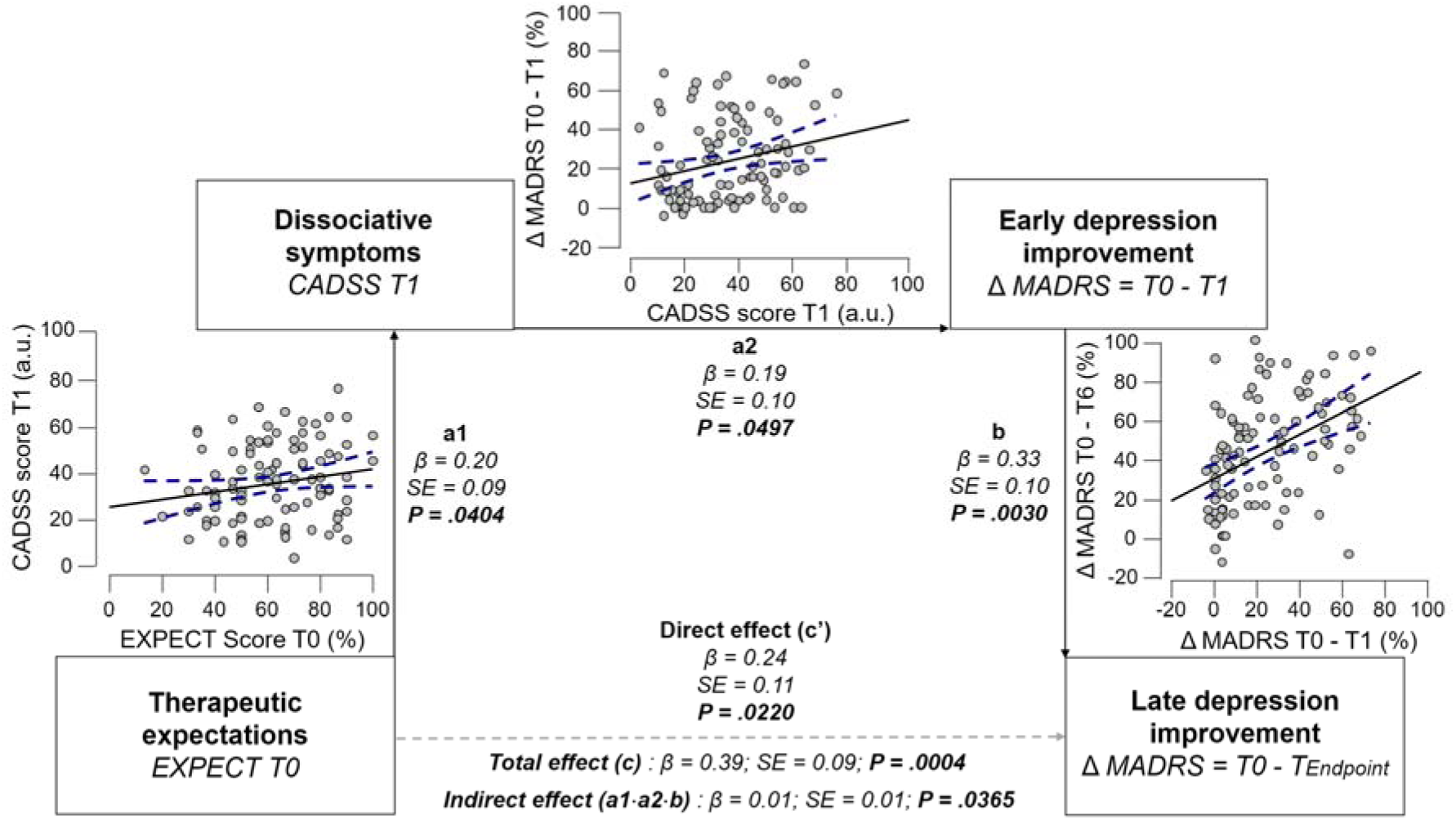
Three-path mediation model linking therapeutic expectations, dissociative symptoms, and depression improvement. This figure illustrates the 3-path mediation model, estimated across all patients regardless of ketamine dose and titration status (n = 100), in which baseline therapeutic expectations were related to end-of-induction depression improvement (ΔMADRS T0–T_Endpoint_) through two sequential mediators: dissociative symptoms at the first infusion (CADSS T1) and early depression improvement (ΔMADRS T0–T1). T_Endpoint_ indicates each participant’s last available assessment during the 6-infusion induction phase. Path coefficients (β) are indicated with standard errors (*SEs*) and P values *(P*). The scatterplots display raw values with each point representing a patient. MADRS, Montgomery-Åsberg Depression Rating Scale; CADSS, Clinician-Administered Dissociative States Scale.

**Figure 6.**
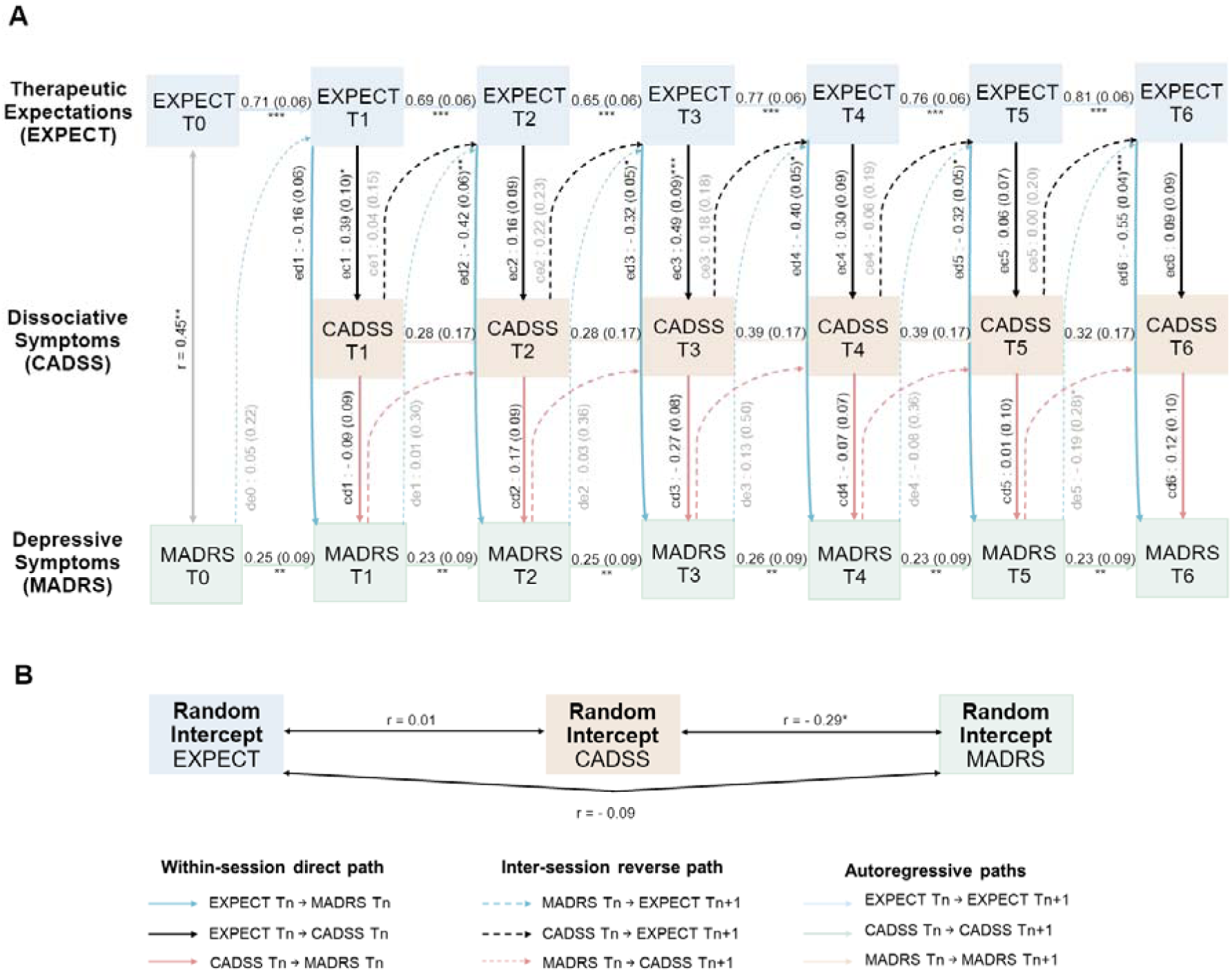
Random intercept cross-lagged panel model of therapeutic expectations, dissociative symptoms, and depressive symptom severity across six ketamine infusions. **(A)** This panel displays the within-person cross-lagged structure of the trivariate RI-CLPM (RI-CLPM1: therapeutic expectations, dissociative and depressive symptoms total scores). The assessment sequence was fixed: therapeutic expectations were assessed before infusion (EXPECT); post-infusion, dissociative symptoms were systematically assessed by the patient (CADSS) prior to depressive symptom severity rated by the clinician (MADRS), within a 1–4 hour post-infusion window once acute dissociative symptoms had clinically dissipated. Directional paths represent the three pre-specified within-session hypotheses: pre-infusion therapeutic expectations predicting post-infusion dissociative symptom intensity (black solid vertical arrows) and depressive symptom severity (blue solid vertical arrows), and dissociative symptoms predicting depressive symptom severity within the same session (red solid vertical arrows). Inter-session reverse paths, included to evaluate the directionality of cross-lagged effects by testing whether prior levels of depressive symptoms or dissociative symptoms at session T_n_ predicted subsequent therapeutic expectations at session T_n+1_, are represented as vertical dashed arrows. Autoregressive paths, reflecting the within-person stability of each variable from session T_n_ to session T_n+1_, are represented as horizontal arrows color-coded to their respective variable: light blue for therapeutic expectations (EXPECT), light green for depressive symptoms (MADRS), and beige for dissociative symptoms (CADSS). Path labels: e, therapeutic expectations (EXPECT); d, depressive symptoms (MADRS); c, dissociative symptoms (CADSS). Within-session paths: ec (EXPECT T_n_ → CADSS T_n_), ed (EXPECT T_n_ → MADRS T_n_), cd (CADSS T_n_ → MADRS T_n_). Inter-session reverse paths: ce (CADSS T_n_ → EXPECT T_n+1_), de (MADRS T_n_ → EXPECT T_n+1_), dc (MADRS T_n_ → CADSS T_n+1_). Path coefficients are standardized, with standard errors in parentheses; asterisks indicate significance levels (* *P* < .05, ** *P* < .01, *** *P* < .001). Path coefficients for within-session directional paths and inter-session reverse paths are displayed in black and gray, respectively, reflecting their status as primary hypotheses and directional control paths. Baseline covariance at T0 and correlations among random intercepts are displayed as bidirectional associations. **(B)** This panel displays the between-person structure, showing correlations among the three random intercepts (RI-EXPECT, RI-CADSS, RI-MADRS), reflecting stable individual differences across the treatment course. Correlation coefficients are reported as Pearson’s r, with asterisks indicating statistical significance (\**P* < .05, ** *P* < .01, *** *P* < .001). EXPECT, therapeutic expectations; CADSS, Clinician-Administered Dissociative States Scale; MADRS, Montgomery-Åsberg Depression Rating Scale.

#### Effect of therapeutic expectations on depressive symptoms

At the within-person level, session-specific therapeutic expectations negatively predicted post-infusion MADRS scores (β*_ed_*) across infusions (T2–T6: β*std* = –0.32 to –0.55, all *P* ≤ .044; T1: β*std* = –0.16*, P* = .346), uniformly negative and predominantly significant. The within-session effect of expectations on depressive symptoms (|β*_ed_*|) exceeded in magnitude its inter-session reverse effect of symptoms on expectations (|β*_de_*|) across five of the six transitions (range: β = –0.29 to 0.43, all *P* ≥ .387; except T5→T6: β = –0.63, *P* = .025). GORICA provided moderate support for this pattern (w = 0.66, ratio = 1.92×). In the bivariate RI-CLPM2 model, excluding dissociative symptoms, β*_ed_* coefficients were of comparable or slightly larger magnitude (β*std* = – 0.34 to –0.62), with stronger GORICA support for the within-session dominance (w = 0.85, ratio = 5.57×), confirming the robustness of this association (Table S11).

#### Effect of therapeutic expectations on dissociative symptoms

Therapeutic expectations positively predicted post-infusion dissociative symptom intensity (β*_ec_*), with coefficients positive at all sessions but reaching significance at T1 and T3 only (β*std* range: 0.06 to 0.49; T1: *P* = .012; T3: *P* < .001; T2, T4–T6: *P* ≥ .089). The within-session expectation-to-dissociation pathway (|β*_ec_*|) consistently exceeded its inter-session reverse effect (|β*_ce_*|) across transitions (range: β = –0.12 to 0.39, all *P* ≥ .093; GORICA: w = 0.89, ratio = 7.71×).

#### Effect of dissociative symptoms on depressive symptoms

The within-person effect of dissociative symptoms to depressive symptoms (β*_cd_*) was non-significant at all sessions (β*_cd_* range = –0.27 to 0.17, *P* ≥ .105). This result was confirmed by RI-CLPM3, excluding potential statistical suppression by therapeutic expectations (Table S12). Likewise, the reverse pathway estimating the effect of depression severity on dissociative symptoms was non-significant (β*_dc_* = 0.01, *P* = .961).

#### Between-person effects

At the between-person level, the covariance between the random intercepts of depressive symptoms and dissociative symptoms was significantly negative (RI-CLPM1: *r* = –0.29, *P* = .030; RI-CLPM3: *r* = –0.33, *P* = .026), indicating that patients who reported higher dissociative symptoms overall tended to show lower depressive symptom severity across the entire treatment course (Figure 6B).

## Discussion

In this 3-week longitudinal observational study of intravenous ketamine in routine care, therapeutic expectations and dissociative symptoms were associated with greater end-of-course antidepressant response. Patients who improved the most also reported increasingly positive expectations throughout treatment. Mediation analyses indicated that this association was partially mediated by dissociative symptom intensity and early clinical improvement after the first infusion. Multivariate analyses showed evidence for distinct roles of therapeutic expectations and dissociative symptoms for antidepressant responsiveness. While therapeutic expectations were a within-person driver at each session, dissociative symptoms were a between-person predictor of depression improvement over the entire treatment course, with less session-specific effectiveness. Together, these findings offer clinically relevant insights into how psychological and pharmacological factors interact during ketamine treatment in routine care.

The patients had moderate to severe depression at baseline; 45% met response criteria after six racemic ketamine infusions, and 12% achieved remission. These rates are consistent with response and remission rates reported in controlled trials and real-world ketamine studies, supporting external validity (46–48). Concomitant monoaminergic antidepressant treatments were largely stable during the induction phase, minimizing pharmacological confounding.

Consistent with prior work in antidepressant and other clinical trials (3,26), therapeutic expectations were strongly associated with antidepressant response following the ketamine induction phase. These results extend prior work by showing that therapeutic expectations increased with the number of infusions only among treatment responders, while they remained unchanged among non-responders. This argues for reciprocal associations between responsiveness and expectations. Experimental findings from placebo analgesia and fast-acting placebo effects of antidepressants have shown that perceived treatment effectiveness can serve as a learning signal that reinforces therapeutic expectations (13,49,50). Here, we show that this pattern is also observed in routine care. Our findings indicate that patients’ perceptions of treatment effectiveness are an important psychological factor associated with changes in clinical symptoms over time.

We took an additional step by examining the role of dissociative symptoms, which were linked to both antidepressant response and therapeutic expectations. Different dissociative dimensions showed distinct patterns of association: higher dissociative intensity, particularly depersonalization and derealization, was more common in patients with greater antidepressant response. Depersonalization and derealization involve temporary shifts in perception and self-related processing that may reflect the context in which therapeutic expectations and antidepressant response co-occur (14,16). Conversely, the amnesia component was not related to treatment response. This result suggests limited recall of the immediate experience, which may reduce the extent to which this aspect of dissociation can be integrated into therapeutic and expectation-related processes.

Our single-level mediation results provide further evidence for an indirect effect of baseline therapeutic expectations on depressive symptoms via the experience of dissociative symptoms and early treatment effects. This indirect effect was statistically significant, but of modest clinical significance (3.2% of the total effect). This result stands at odds with the idea that dissociations play a clinically meaningful mechanistic role for antidepressant treatment responsiveness (17,18,20). They rather suggest that patients with higher expectations tended to experience more intense dissociative symptoms at the first infusion, consistent with the hypothesis that pre-treatment expectations shape the perceived intensity of subjective drug effects (51). Dissociative symptoms may therefore be associated with cognitive-affective changes relevant to depression, including alterations in self-related and emotional processing (49–51). In this context, therapeutic expectations may structure the appraisal of these altered experiences, consistent with the mediation finding that dissociative symptoms at the first infusion do not exert an independent effect on clinical improvement but instead reflect part of the expectation–outcome association. One limitation of our single-level mediation analysis is that it assessed overall associations at the group level. We cannot infer individual patient– and session-specific effects or their contribution to improvement in depression. Thus, the single-level mediation analyses identified potential targets for intervention. On the contrary, our multivariate cross-lagged analyses allowed us to shed light on the individual, session-to-session directionality of effects, and which effects were more plausible. For example, whether patients who expected greater treatment effectiveness experienced greater improvement, or whether those who improved formulated stronger expectations.

Our findings from three different, but complementary, random intercept cross-lagged models disentangled whether changes in expectations, dissociative symptoms, or depressive symptoms occurred first. Across the full induction course, the directional dominance of expectations over depressive symptoms was about 2 (trivariate RICLM) to 6 (bivariate RICLM) times stronger than the reverse effect. Likewise, the directional dominance of expectations on dissociative symptoms was about 8 times stronger than its reverse effect. These findings validate the assumptions of the mediation model. Interestingly, the reverse effect of depressive symptoms on therapeutic expectations became significant by the end of the induction course. This result is consistent with our earlier finding that therapeutic expectations increased with the number of infusions only among responders, and suggests that expectations and improvement in depressive symptoms may mutually reinforce each other in treatment responders.

Our finding that therapeutic expectations also predicted acute dissociative symptom intensity, with coefficients remaining positive across sessions but reaching statistical significance only at T1 and T3, favors the hypothesis that patients’ beliefs about treatment effectiveness partly shape the perceived intensity of the treatment’s acute subjective effects (51–54). Expectancy-based influences on subjective experiences may be particularly salient during early treatment exposures, when patients rely more heavily on prior beliefs to interpret an unfamiliar pharmacological state, as reflected in ketamine-induced dissociative and perceptual alterations (26,53,55). As they progress in their treatment, the accumulation of direct experience may gradually update these appraisals, reducing the relative contribution of expectations to the evaluation of dissociative symptoms. (26,51,55). In contrast, dissociative symptoms showed no within-person association with subsequent depressive symptom severity from session to session. This finding may account for inconsistencies across studies regarding the association between dissociative symptoms and antidepressant response (18–23), which often confound between-person dissociative symptoms and session-specific within-person fluctuations (38). At the between-person level, however, patients with higher dissociative symptoms showed more depressive symptom improvement across the entire treatment course. This finding suggests that for some patients, dissociative symptoms may help predict improvement during certain sessions, but not consistently across all sessions. However, across the entire induction course, patients with higher overall levels of dissociative symptoms tended to benefit most from ketamine antidepressant treatment. These results are in line with the mediation results, which also reflected this between-person effect. In line with the literature, experiencing dissociative symptoms may thus represent an overall source of variance between patients in antidepressant effects (17).

Together, these findings suggest that therapeutic expectations and dissociative symptoms have distinct effects: Expectations function as a dynamic within-person driver of antidepressant response, whereas dissociation serves as a between-person marker of overall clinical improvement. These distinct profiles highlight complementary clinical targets: optimizing therapeutic expectations at each infusion may enhance antidepressant response, while identifying patients who display dissociative symptoms may help guide patient stratification and supportive care (4,8,56).

Future work should examine affective evaluations of the dissociative experience following ketamine treatment. In the present study, dissociative symptoms were measured with the CADSS, which does not capture other experiential dimensions, such as emotional responses or non-dissociative perceptual changes, that may also shape treatment outcomes. The emotional aspect of dissociative experiences can vary between sessions, which may explain why there are no consistent effects of dissociative symptoms on depressive symptoms within a person from one session to the next. Likewise, broader contextual and social factors, including elements of the therapeutic setting and clinician–patient relationship, were not formally assessed (55). These affective and interpersonal influences warrant closer investigation.

Investigating expectation effects within routine medical care is a relatively rare approach, offering the opportunity to observe real-world, longitudinal patterns across successive treatment visits. However, this design also leads to missing assessments due to scheduling constraints or early discontinuation. Procedural consistency was supported by a standardized protocol that included structured preparation and post-session debriefings (28).

In conclusion, this observational study among patients with moderate to severe depression provides convergent evidence that therapeutic expectations and acute dissociative symptoms were associated with antidepressant response over the course of treatment. Within a broader “set and setting” framework (55), our study primarily captures the “set” of ketamine’s antidepressant effects, notably the role of patients’ expectations in how they experience and appraise its pharmacological effects. Overall, these findings clarify how psychological and pharmacological factors may interact during antidepressant treatment in routine care, which is helpful for understanding and optimizing treatment effectiveness and patient stratification.

## Article Information

### Corresponding Author

Willys Cantenys, MD–PhD student, and Liane Schmidt, PhD, Paris Brain Institute, Pitié-Salpêtrière Hospital, 47 Blvd de l’Hôpital, Paris, France.

### Author Contributions

Dr. Cantenys and Dr. Schmidt were responsible for accessing the complete dataset and ensuring the accuracy and integrity of the data analyses. Drs. Fossati and Schmidt share equal senior authorship and jointly supervised the study.

*Study concept and design*: Cantenys, Fossati, Schmidt.

*Data acquisition: Cantenys,* Yoldas, Masset, Romier, Samion, Imbault, Tran, Megda Garcia, Soltani, Marradi, Ho, Gohier, Doulazmi, Chesneau, Jadaan, Bulut, Djonouma, Charradi, Claret Tournier.

*Statistical analysis*: Cantenys, Schmidt.

*Drafting of the manuscript*: Cantenys, Fossati, Schmidt.

*Critical revision of the manuscript for important intellectual content*: Cantenys, Fossati, Schmidt.

*Administrative, technical, or material support*: Cantenys, Masset, Claret Tournier, Fossati, Schmidt.

*Study supervision*: Fossati, Schmidt.

### Additional Contributions

We thank the medical and nursing staff, including Yvette Byll and all nurses at the Adult Psychiatry department of Pitié-Salpêtrière Hospital. We acknowledge the valuable contribution of Flavia Megda Garcia † and respectfully honor her memory.

## Data Availability

Fully anonymized data were used in this study, with no possibility of re-identification. The datasets are not publicly available because they derive from retrospective clinical data collected in routine care and are subject to ethical and institutional restrictions. Data may be made available from the corresponding author upon reasonable request, subject to approval by the relevant institution and applicable regulations.

## Acknowledgment of AI Use

The authors used *Nature Research Assistant* (© 2025 Springer Nature; accessed December 2025) to assist with language editing, improving clarity, rephrasing, and structuring portions of the manuscript. The authors reviewed and edited the AI-assisted text and take full responsibility for the integrity and accuracy of the manuscript content.

