## Supplementary Material for "Distinct Roles of Therapeutic Expectations and Dissociative Symptoms in Antidepressant Response During Ketamine Treatment in Routine Care"

**Supplemental Online Content**

**Methods**

**Methods S1.** Intravenous Ketamine Administration Protocol

**Methods S2.** Data Quality and Management of Missing Data

**Methods S3.** Details on the Assessment of Dissociative Symptoms – CADSS Score

**Methods S4.** Details on the Assessment of Therapeutic Expectations

**Methods S5.** Sensitivity Analyses at Constant Ketamine Dosage

**Methods S6**. Baseline Associations between Depression Severity, Therapeutic Expectations, and Dissociative Symptoms

**Methods S7.** Detailed Specification of Univariate Linear Models

**Methods S8.** Overall Model Fit Diagnostics LME1 to LME3

**Methods S9**. Cross-Validation of LME1

**Methods S10.** Overall Model Fit Diagnostics LME4 and LME5

**Methods S11.** Cross-Lagged Mixed Models

**Figures**

**Figure S1.** Distribution of Titrated Ketamine Dosages

**Figure S2**. Completeness matrices for MADRS, CADSS, and EXPECT ratings

**Figure S3.** Longitudinal Stability and Internal Consistency of Therapeutic Expectations

**Figure S4.** Quantile–Quantile (Q–Q) plots of standardized residuals for LME1 to LME3

**Figure S5.** Residuals versus Fitted Values for Longitudinal LME1 to LME3

**Figure S6.** Correlation of Predicted MADRS Scores from LME1 and Observed MADRS Scores

**Figure S7.** Distribution of treatment responses

**Figure S8.** Quantile–Quantile (Q–Q) plots of standardized residuals for LME4 and LME5

**Figure S9.** Residuals versus Fitted Values for LME4 and 5

**Tables**

**Table S1.** Longitudinal Analysis: Linear Mixed-Effects Model 1 of Depression

**Table S2**. Longitudinal Analysis: Linear Mixed-Effects Model 2 of Depression

**Table S3**. Longitudinal Analysis: Linear Mixed-Effects Model 3 of Depression

**Table S4**. Session-Level Analysis: Linear Regression Model of Depression Predicted by Therapeutic Expectations and Dissociative Symptoms

**Table S5**. Session-Level Analysis: Linear Regression Model of Depression Predicted by Therapeutic Expectations and Dissociative Symptom Dimensions

**Table S6.** Dimensional Analysis: Linear Mixed-Effects Model 4 of Therapeutic Expectations

**Table S7**. Dimensional Analysis: Linear Mixed-Effects Model 5 of Dissociative Symptoms

**Table S8.** Three-Path Mediation Model with Average Therapeutic Expectations, and Dissociative Symptom Scores

**Table S9**. Three-Path Mediation Model with Dissociative Symptom Dimensions

**Table S10.** Multivariate Cross-Lagged Analysis: Trivariate RI-CLPM of Therapeutic Expectations, Dissociative Symptoms, and Depressive Symptoms

**Table S11.** Multivariate Cross-Lagged Analysis: Bivariate RI-CLPM of Therapeutic Expectations and Depressive Symptoms

**Table S12.** Multivariate Cross-Lagged Analysis: Bivariate RI-CLPM of Dissociative Symptoms and Depressive Symptoms

**All analyses at constant ketamine dosage of 0.5 mg/kg:**

**Table S13.** Linear Mixed Effects Model 1 of Depression

**Table S14**. Linear Mixed Effects Model 2 of Depression

**Table S15**. Linear Mixed Effects Model 3 of Depression

**Table S16**. Session-Level Regressions of Therapeutic Expectations and Dissociative Symptoms

**Table S17**. Session-Level Regressions of Dissociative Symptom Dimensions

**Table S18**. Linear Mixed-Effects Model 4 of Therapeutic Expectations

**Table S19.** Linear Mixed-Effects Model 5 of Dissociative Symptoms

**Table S20.** Three-Path Mediation Model with Average Therapeutic Expectations, and Dissociative Symptom Scores

**Table S21.** Three-Path Mediation Model with Dissociative Symptoms Dimensions

**References**.

This supplementary material has been provided by the authors to give readers additional information about their work.

**Methods S1.** Intravenous Ketamine Administration Protocol

The induction phase of intravenous ketamine treatment at Pitié-Salpêtrière Hospital involves six sessions and a dose-titration approach based on clinical response assessed by the Montgomery-Åsberg Depression Rating Scale (MADRS). This phase aims for a rapid and meaningful antidepressant response. The protocol starts with a dose of 0.5 mg/kg given over the first two to three infusions. If there is no clinically significant response—defined as less than a 50% reduction in MADRS score—the dose is increased to 0.75 mg/kg. If an adequate response still does not occur after another two to three sessions at this dose, the dose is raised to 1.0 mg/kg. This personalized titration approach is designed to maximize therapeutic benefit while reducing adverse effects during the induction phase (1).

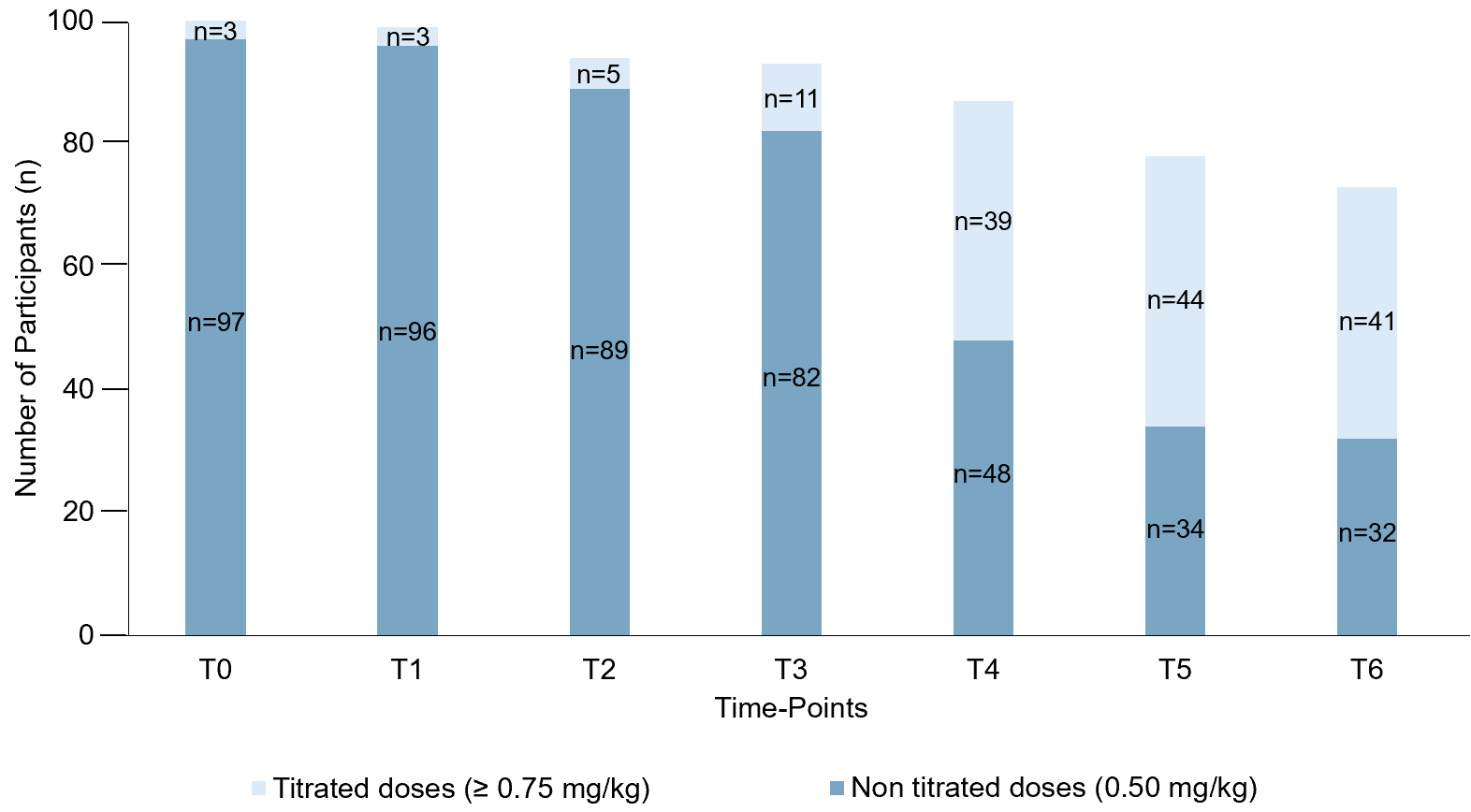
**Figure S1. Distribution of Ketamine Dosages.** Stacked bars represent the cumulative number of patients who continued receiving the standard 0.5 mg/kg ketamine dose (*dark blue*) and those who underwent dose titration to 0.75 mg/kg (*light blue*) across the six infusion sessions of the induction phase. No infusion was administered at T0 (baseline assessment only). All patients initially received 0.5 mg/kg at the first infusion (T1); titration occurred only in cases of insufficient clinical response. These data illustrate the composition of the analytic samples used in the main and sensitivity analyses: (1) all patients, irrespective of dose, and (2) those who remained at 0.5 mg/kg throughout the induction phase. The progressive decrease in total n across time points reflects expected attrition during the six-session induction phase, as not all patients completed every infusion. This unbalanced structure was handled in all models using mixed-effects analyses accounting for within-subject variance and missing-at-random data patterns.

**Methods S2.** Data Quality and Management of Missing Data

**Data completeness**

Across the seven assessment points, including baseline (T0) and six intravenous ketamine infusions (T1–T6), data were planned for 700 assessment occasions (7 sessions × 100 participants). EXPECT and CADSS ratings were available for 624 of 700 planned assessment occasions (89.1%), whereas MADRS ratings were available for 664 of 700 (94.9%). Missing data were mainly due to:

- Premature treatment discontinuation due to poor clinical response or adverse tolerability (n = 14 patients, 36 observations);
- Incomplete self-reported questionnaires (expectation or dissociation scales not completed; n = 19 patients, 40 observations).

All missing observations were prospectively recorded in the research database with notes on their clinical or procedural cause.

**Data quality control procedures**

Data were collected in real-world clinical settings at the Department of Psychiatry, Pitié-Salpêtrière Hospital. To promote data quality and integrity, **(1)** all clinical assessments were conducted by psychiatrists or psychiatry residents trained in standardized MADRS administration as part of the clinical care procedures at Pitié-Salpêtrière Hospital. **(2)** Data entry followed standardized procedures, with periodic quality checks by the study investigator. **(3)** Data consistency and completeness were reviewed monthly by the study investigator, and any discrepancies were corrected through source verification in the medical records. These measures were implemented to support data quality and integrity in this clinical dataset.

**Description and management of missing data**

Following the *Treatment and Reporting of Missing Data in Observational Studies (TARMOS)* framework (2), missing data were characterized and handled in three sequential steps:

1. **Preplanning of analysis and documentation of missingness source**

Given the retrospective observational design within routine clinical care, attrition and partial missingness were anticipated (e.g., treatment discontinuation, incomplete self-reports). Reasons for missing observations were systematically recorded at each session. The primary analyses used likelihood-based linear mixed-effects models, which incorporate all available observations. Given the modest proportion of missingness (range, 5.1%–10.9% across measures) and this likelihood-based analytical framework, multiple imputation was not pursued. The robustness of the main findings was examined in secondary analyses focusing on sessions administered at the standard 0.5 mg/kg dose (Tables S10–S18 in the Supplement).

2. **Exploration of potential causes for missing data points**

At the variable level, MADRS scores were missing for 36 of 700 observations (5.1%), while EXPECT and CADSS each had 76 missing values (10.9%). Visual inspection of patient-by-timepoint completeness matrices suggested a predominantly monotone dropout pattern, in which missingness clustered after early treatment discontinuation rather than appearing as sporadic omissions within otherwise complete trajectories (Figure S2). This pattern is consistent with the attrition typical of longitudinal real-world treatment datasets. To understand the nature of missingness, mixed-effects logistic regression models were fitted at the session level (n = 700), with a random intercept for participant (1|ID), predicting each variable's missingness indicator from baseline covariates (age, sex, education level, and MADRS baseline severity).

Depression scores measured by the Montgomery-Åsberg Depression Rating Scale (MADRS)

In mixed-effects logistic regression models with a random intercept for participant, missing MADRS ratings were not significantly associated with baseline age (*OR* = 1.00; *95% CI*, 0.97 to 1.04; *P* = .88), sex (*OR* = 1.59; *95% CI*, 0.55 to 4.63; *P* = .39), education level (*OR* = 1.33; *95% CI*, 0.76 to 2.34; *P* = .32), or MADRS baseline severity (*OR* = 1.05; *95% CI*, 0.96 to 1.15; *P* = .30). The lack of systematic demographic or clinical predictors suggests that missing MADRS observations were not strongly associated with the measured baseline covariates.

Therapeutic expectations (EXPECT) and dissociative symptoms (CADSS)

In mixed-effects logistic regression models with a random intercept for participant, missingness in CADSS/EXPECT ratings was not significantly associated with baseline age (*OR* = 0.99; *95% CI*, 0.96 to 1.01; *P* = .36), sex (*OR* = 2.01; *95% CI*, 0.83 to 4.84; *P* = .12), education level (*OR* = 1.20; *95% CI*, 0.76 to 1.90; *P* = .43), or MADRS baseline severity (*OR* = 1.04; *95% CI*, 0.96 to 1.12; *P* = .34). Importantly, missing values most often occurred after early treatment discontinuation due to poor tolerability or insufficient response, a pattern compatible with a predominantly monotone dropout process.

3. **Overall Interpretation**

Given the modest overall proportion of missing data (MADRS: 36/700 (5.1%); EXPECT and CADSS: 76/700 each (10.9%)), its mostly monotone dropout pattern, and the lack of strong associations between missingness and baseline depressive severity or sociodemographic characteristics, no imputation was performed. All longitudinal analyses were therefore conducted using likelihood-based linear mixed-effects models (estimated by REML), which use all available data and are commonly interpreted under a MAR working assumption conditional on observed covariates. Because the missing-data mechanism is not directly identifiable from the observed data, we treated MAR conditional on observed covariates as a working assumption for the primary likelihood-based mixed-effects analyses and its plausibility was assessed by examining recorded reasons for missingness, patterns of missingness, and predictors of missingness.

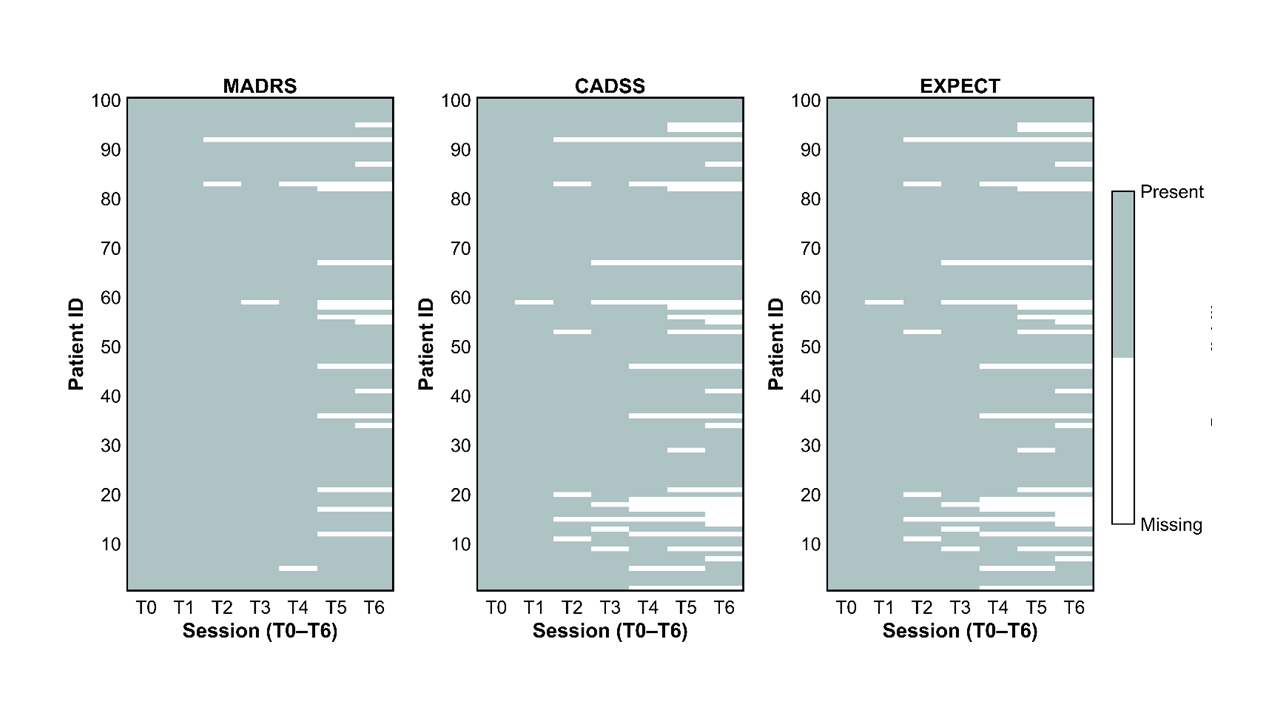

**Figure S2. Completeness matrices for MADRS, CADSS, and EXPECT ratings.** Each panel displays a completeness matrix for one outcome (MADRS, CADSS, or EXPECT) across baseline (T0) and the six intravenous ketamine infusions (T1–T6). Each row represents one patient, and each column represents an assessment time point. Colored cells denote available data, while white cells indicate missing observations.

**Methods S3**. Details on the Assessment of Dissociative Symptoms – CADSS Score

Dissociative symptoms were evaluated in the hours following each ketamine session using the Clinician-Administered Dissociative States Scale (CADSS), French version. The CADSS consists of 23 self-rated items assessing transient dissociative phenomena on a 5-point scale. Patients were instructed to indicate how they experienced the ketamine session by rating the intensity of each effect from 0 (minimum) to 4 (maximum).

The dimensional structure of the CADSS encompasses three theoretically and empirically distinct domains—depersonalization, derealization, and amnesia—that capture complementary facets of the dissociative state.

- **Depersonalization** reflects a disruption in self-perception, characterized by feelings of detachment from one’s body or sense of identity (“observer perspective”);
- **Derealization** captures perceptual distortions of the external environment, such as unreality, visual alteration, or sensory modifications;
- **Amnesia** refers to lapses in memory or continuity of experience occurring during the altered state.

This tripartite structure is consistent with both the original conceptualization of the CADSS (3) and subsequent applications in pharmacological and psychiatric contexts (4,5). In line with prior validation work for ketamine-induced dissociation (4), the CADSS has demonstrated high internal consistency (*Cronbach’s α* = 0.94) and sensitivity to dose-dependent effects of intravenous ketamine (0.5–1.0 mg/kg). In that study, dissociation increased in a dose-dependent manner, supporting the scale’s reliability and construct validity in this pharmacological context.

The CADSS dissociative dimensions were derived from raw item sums as follows: depersonalization (items 3–7, 20, 23), derealization (items 1, 2, 8–13, 16–19, 21), and amnesia (items 14, 15, 22). For each dimension, a raw total score was computed by summing the relevant items. A global CADSS total score was also calculated as the sum of all 23 raw items. All raw total scores (dimensional and global) were then z-standardized across patients before inclusion in the statistical models. Higher values indicated higher dissociative symptom levels during an infusion of ketamine at a given session.

**Methods S4**. Details on the Assessment of Therapeutic Expectations

Therapeutic expectations were evaluated using a composite score derived from the patients’ self-reported ratings regarding antidepressant effectiveness, probability of recovery, and post-treatment depression intensity. Patients responded on a visual analog scale ranging from 0% (minimum) to 100% (maximum effect). Ratings were averaged, with higher averages indicating greater therapeutic expectations The ratings comprised three equally weighted questions:

[1] "To what extent do you think ketamine treatment is effective against depression?"

[2] "To what extent do you think your depression will be cured after ketamine treatment?"

[3] "What will be the intensity of your depression after ketamine treatment?" (reverse scored).

The internal consistency of this composite score was assessed using Cronbach’s α with 95% confidence intervals (6,7). At baseline (T0), the ratings showed good internal consistency (*Cronbach’s α* = 0.82) and balanced inter-item correlations, indicating coherent but non-redundant components of expectations:

- *Efficiency* x *Recovery*: *r* = 0.62, *P* < .001
- *Efficiency* x *Intensity*: *r* = 0.55, *P* < .001
- *Recovery* x *Intensity*: *r* = 0.66, *P* < .001

Principal component analysis confirmed a unidimensional structure, explaining 74% of the total variance, with homogeneous loadings across items (0.56–0.59), supporting a single latent expectation factor.

As shown in Figure S3, across the seven assessment time points, from baseline (T0) to the sixth infusion (T6), Cronbach’s α increased from 0.82 to 0.94 (*mean*(SD) = 0.90 (0.04)), and inter-item correlations rose in parallel (*r* = 0.55–0.87, all *P* < .001), a pattern consistent with convergence of expectation subcomponents without redundancy. The unidimensional structure remained stable across time points (first component accounting for >70% of the variance). To ensure robustness, 10 000 bootstrap resamples were used to estimate non-parametric 95% confidence intervals for α. Internal consistency increased from 0.82 [*95% CI*, 0.75 to 0.87] at baseline to 0.94 [*95% CI*, 0.92 to 0.96] at the last infusion. Confidence intervals did not overlap between the early and late sessions, supporting a genuine increase in internal consistency rather than an effect of sampling variability.

Overall, the three therapeutic expectation questions consistently measured the same underlying expectation construct, each providing distinct yet complementary information about patients’ therapeutic expectations toward ketamine.

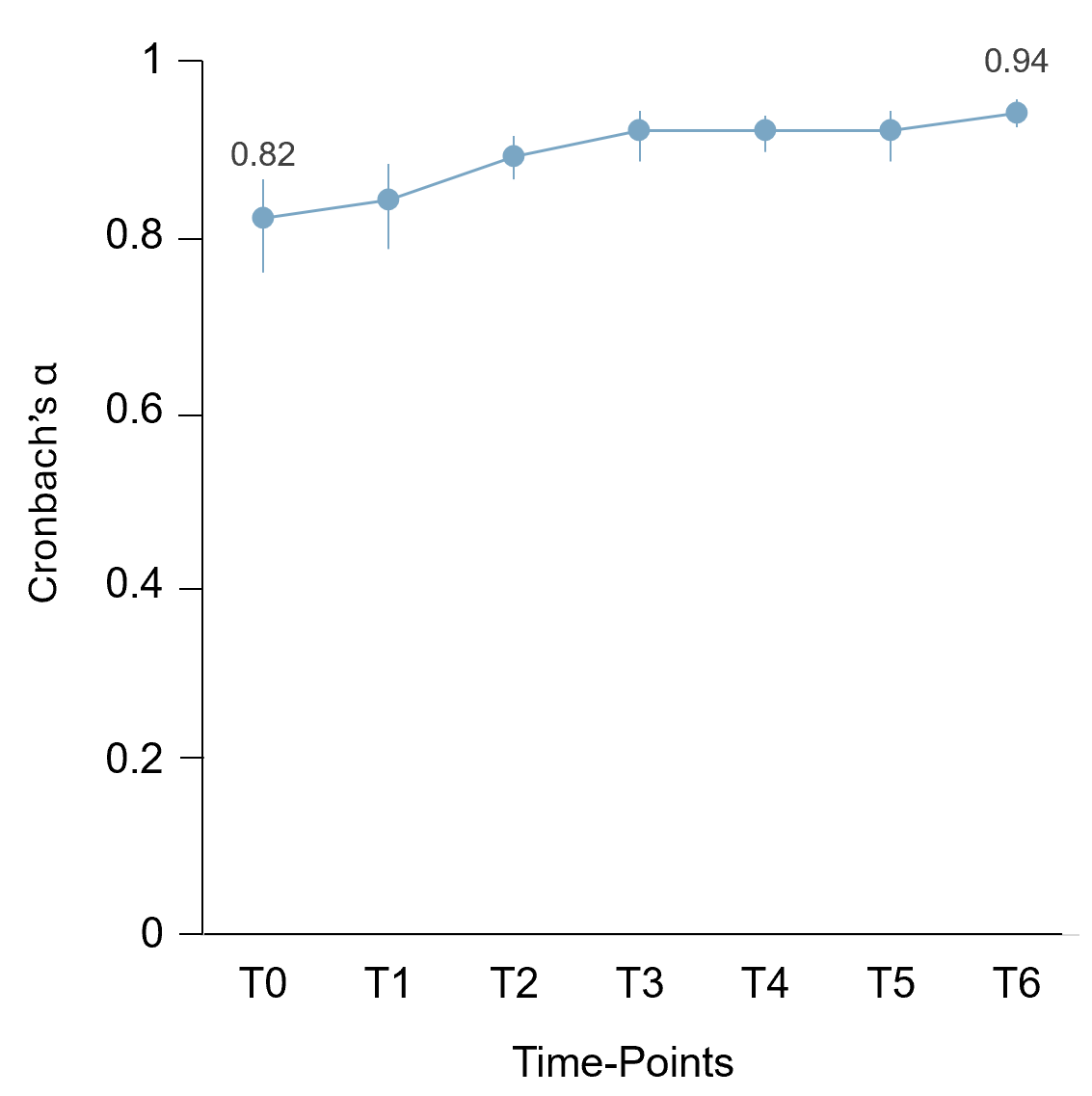
**Figure S3. Longitudinal Stability and Internal Consistency of Therapeutic Expectations.** The line plot shows Cronbach’s α values for the three therapeutic expectation items across treatment sessions (T0–T6), with error bars representing 95% confidence intervals (time points from baseline (T0) to the end-of-course session (T6)).

**Methods S5**. Sensitivity Analyses at Constant Ketamine Dosage

**Rationale**

The personalized titration approach during the induction phase led to varying cumulative ketamine exposures among patients. Most started treatment at 0.5 mg/kg, but three patients with previous ketamine treatment began directly at 0.75 mg/kg, the dose previously shown to be effective. Patients who experienced less than a 50% reduction in MADRS scores after two to three infusions were first increased to 0.75 mg/kg; if response remained insufficient after a further two to three infusions, the dose was subsequently increased to 1.0 mg/kg, based on clinical judgment (Methods S1). As a result, different treatment paths emerged for each patient, varying in both timing and dose size.

Previous studies have indicated that higher ketamine doses may produce stronger dissociative symptoms (4,8) and, in some cases, greater antidepressant effects (9). Therefore, distinguishing patients who underwent dose titration from those maintained at the standard 0.5 mg/kg dose helps to clarify whether observed long-term associations—such as those linking expectation, dissociation, and symptom improvement—reflect treatment-related processes or are partly influenced by dose-dependent pharmacodynamic effects.

The distinction between patients who underwent titration and those kept at 0.5 mg/kg aims to reduce bias related to clinical decision-making, since dose escalation is usually triggered by early nonresponse. Without considering this process, poorer early outcomes could lead to higher doses, complicating the interpretation of associations between symptom trajectories and dose adjustments.

**Systematic Replication of All Main Analyses at Constant Ketamine Dosage**

To address heterogeneity in cumulative ketamine exposure, all analyses were initially conducted on the full sample (all-patient analyses, Tables S1–S9 in the Supplement), regardless of dose, and then replicated using all sessions administered at the standard 0.5 mg/kg dose (sensitivity analyses, Tables S13–S21 in the Supplement). These replications aimed to check for the potential confounding effect of dose escalation on both antidepressant and dissociative outcomes.

All main models described in the manuscript—including linear mixed-effects models (LME1–5), timepoint-specific regression analyses, and the 3-path mediation model—were consistently replicated in both datasets to assess the robustness of findings across dosing conditions. Results are reported in Tables S13 to S21.

These convergent findings indicate that the reported relationships among expectation, dissociation, and antidepressant response were robust to dose titration. The ketamine dosage was associated with higher levels of dissociative symptoms but did not modify the pattern of associations linking dissociation and therapeutic expectations with clinical improvement trajectories.

**Methods S6**. Baseline Associations between Depression Severity, Therapeutic Expectations, and Dissociative Symptoms

To examine how the core clinical and psychological variables were related at baseline, and to assess whether the main longitudinal and session-level findings might have been influenced by pre-existing biases, a set of control analyses was conducted.

First, we analyzed the initial relationship between depressive severity and therapeutic expectations by calculating a Pearson correlation between the MADRS total score and baseline EXPECT score. This analysis revealed only a weak positive relationship (*r* = 0.20, *P* = .046; *R²* = 0.04), suggesting that expectation reflects a construct largely separate from baseline depression severity. In similar multivariable models, the variance inflation factors for MADRS and expectation were close to 1 (*VIF* = 1.04), indicating no problematic multicollinearity between these predictors.

Second, to examine whether end-of-course clinical outcomes differed by initial depression severity, we compared baseline MADRS scores across the three MADRS-based improvement categories and the three clinician-rated response categories (nonresponders, responders, and remitters) using separate one-way ANOVAs. These analyses showed no significant main effect of group on baseline severity (*F(2,97)* = 1.97, *P* = .15), suggesting that later outcome differences were unlikely to be attributable to baseline severity.

Third, we assessed whether baseline therapeutic expectations were associated with dissociative symptom severity at each infusion session. Six separate linear regression models were run with CADSS scores at each infusion (T1–T6) as the dependent variable and baseline expectations (T0) as the only predictor. Across sessions, expectation showed only small, positive, and nonsignificant relationships with dissociation (*β* = 0.03–0.16; all *P* = .07–.80; all *R²* = 0.001–0.03), with the strongest but still nonsignificant effect observed at the first infusion (T1: *β* = 0.16, *SE* = 0.09, *P* = .07; *R²* = 0.03). No association reached the Bonferroni-corrected significance level for six tests (*α* = .0083), suggesting that baseline therapeutic expectations did not significantly relate to the severity of dissociative symptoms at any infusion session.

Taken together, these control analyses show that the main relationships among therapeutic expectations, dissociative symptoms, and depressive improvement were unlikely to be primarily explained by baseline depression severity, overlapping constructs, or initial therapeutic expectations-driven differences in dissociative responses.

**Methods S7.** Detailed Specification of Univariate Linear Models

**Univariate effects: linear regression and mixed-effects analyses**

To test the hypothesis that self-reported therapeutic expectations and dissociative symptoms were associated with depressive symptoms measured during a 3-week ketamine treatment course for depression, the dataset was analyzed using complementary longitudinal, session-level, and categorical linear models. In all analyses, dissociative symptoms were analyzed using both the total CADSS score and its depersonalization, derealization, and amnesia subscales. A sensitivity analysis restricted to sessions administered at the fixed 0.5 mg/kg dose was conducted to control for potential dose effects (Methods S5). Baseline associations among depressive severity, therapeutic expectations, and dissociative symptoms, as well as group comparability and collinearity diagnostics, were evaluated in separate control analyses at T0 (Methods S6).

(1) Longitudinal analysis of depressive symptom trajectories

Longitudinal associations between therapeutic expectations, dissociative symptoms, and depressive symptom severity were examined using linear mixed-effects models as implemented in the fitlme function in MATLAB. Three longitudinal models were tested, with three separate LMEs that were fitted to depression scores (MADRS):

LME1: Joint longitudinal associations of therapeutic expectations and dissociative symptoms with depression

*MADRS_ij_ ~ (β₀ + β₁·timeᵢⱼ + β₂·CADSSᵢⱼ + β_3_·EXPECTᵢⱼ + β_4_·(time:CADSS)ᵢⱼ+ β_5_·(time:EXPECT)ᵢⱼ + β_6_·(CADSS:EXPECT)ᵢⱼ + β_7_·(time:EXPECT:CADSS)ᵢⱼ) + β_8_·ageⱼ + β_9_·educationⱼ+ β_10_·sexⱼ + (u₀ⱼ + u₁ⱼ·timeᵢⱼ) + εᵢⱼ*

LME2: Longitudinal associations of therapeutic expectations with depression

*MADRS_ij_ ~ (β₀ + β₁·timeᵢⱼ + β_2_·EXPECTᵢⱼ + β_3_·(time:EXPECT)ᵢⱼ + β_4_·ageⱼ + β_5_·educationⱼ+ β_6_·sexⱼ +(u₀ⱼ + u₁ⱼ·timeᵢⱼ) + εᵢⱼ*

LME3: Longitudinal associations of dissociative symptoms with depression

*MADRS_ij_ ~ (β₀ + β₁·timeᵢⱼ + β₂·CADSSᵢⱼ + β_3_·(time:CADSS)ᵢⱼ + β_4_·ageⱼ + β_5_·educationⱼ+ β_6_·sexⱼ + (u₀ⱼ + u₁ⱼ·timeᵢⱼ) + εᵢⱼ*

Each mixed-effects model thus estimated how MADRS scores evolved across consecutive ketamine infusions, while accounting for both between-patient differences and within-patient changes over time (time indexed by *i* and patient by *j*). The time regressor represented treatment sessions from the first to the sixth infusion. The LME1 model included EXPECT, CADSS, and their 2- and 3-way interactions with time to examine their combined and interactive associations with depressive symptom severity. All models were adjusted for age, education level, and sex. To control for individual differences in overall depression severity and its evolution from the first to the sixth ketamine infusion, each model included a random intercept (*u₀ⱼ*) and random slopes for time (*u₁ⱼ*).

All variables, dependent and independent, were standardized using z-scores based on the overall mean and standard deviation of the entire sample (i.e., patients and testing time points). The models were estimated using maximum likelihood (MLE). More details about the respective model fits and cross-validation can be found in Methods S8 and S9 and Figures S4–S6.

(2) Session-level effects of therapeutic expectations and dissociative symptoms

To complement the longitudinal analysis, session-level models examined the association between global depression improvement and session-specific therapeutic expectations or dissociative symptoms at each time point (T0: baseline, T1–T6: first to sixth infusion). Global improvement in depression was expressed as the percentage change in MADRS from baseline (T0) to each patient’s endpoint (T_Endpoint_), defined as the last available MADRS assessment during the 3-week ketamine treatment course:

$$\%\text{Improvement}_{\text{Baseline to Endpoint}}=\frac{\text{MADRS}_{\text{Baseline}}-\text{MADRS}_{\text{Endpoint}}}{\text{MADRS}_{\text{Baseline}}}.$$

A total of 13 linear regression models were fitted:
– 7 models for EXPECT ratings (baseline to session 6);
– 6 models for CADSS ratings (sessions 1 to 6).

$$\%\text{Improvement}_{\text{Baseline to Endpoint}}=\beta_{0}+\beta_{1}\text{ }\text{Rating}_{t}+\beta_{2}\text{ age}+\beta_{3}\text{ education}+\beta_{4}\text{ sex}+\varepsilon,$$

All predictors were z-scored at each time point. Coefficients therefore quantify the change in global improvement per 1–SD increase in EXPECT or CADSS. Statistical significance was assessed using Bonferroni correction for 13 comparisons.

In additional models, the three dimensions of dissociative symptoms—depersonalization, derealization, and amnesia—were also tested for their session-level associations with end-of-course depression improvement.

(3) Categorical analysis of therapeutic expectations and dissociative symptoms across response groups

Our third analysis, complementary to the two previous ones, examined whether dissociative symptoms and expectations differed along the clinical trajectories across responder groups.

To this end, patients were grouped into low responders (<25% reduction in MADRS score), moderate responders (25–75% reduction), and high responders (≥75% reduction). For example, participants showing a 15% improvement were classified as low responders, those showing a 50% improvement as moderate responders, and those showing an 80% improvement as high responders (Figure S7).

Two separate linear mixed-effects models (LME4 and LME5) were fitted to standardized (z-scored) therapeutic expectations (*EXPECT*) and dissociation scores (*CADSS)* as follows:

LME4: Trajectories of therapeutic expectations across responder groups

*EXPECT_ij_ ~ (β₀ + β₁·timeᵢⱼ + β₂·groupⱼ + β_3_ (time:group)ᵢⱼ + β_4_·ageⱼ + β_5_·educationⱼ + β_6_·sexⱼ + (u₀ⱼ + u₁ⱼ·timeᵢⱼ) + εᵢⱼ*

LME5: Trajectories of dissociative symptoms across responder groups

*CADSS_ij_ ~ (β₀ + β₁·timeᵢⱼ + β₂·groupⱼ + β_3_ (time:group)ᵢⱼ + β_4_·ageⱼ + β_5_·educationⱼ + β_6_·sexⱼ + (u₀ⱼ + u₁ⱼ·timeᵢⱼ) + εᵢⱼ*

The models included the *group × time* interaction as a fixed effect, with random intercepts (*u₀ⱼ*) and time-by-participant slopes (*u₁ⱼ·timeᵢⱼ*). They were adjusted for age, sex, and education level (Tables S6 and S7 in the Supplement).

Both LME4 and LME5 were estimated by restricted maximum likelihood (REML) using the *fitlme* function in MATLAB. Model fits and diagnostics are reported in Methods S10 and Figures S8 and S9.

**Methods S8.** Overall Model Fit Diagnostics LME1 to LME3

The assumptions for the linear mixed-effects models were assessed using Q–Q plots of residuals and scatterplots of residuals versus fitted values (Figures S4 and S5). Residuals were normally distributed and showed no signs of heteroscedasticity. Missing data were handled under the working assumption that data were missing at random (MAR) for the likelihood-based mixed-effects models, which uses all available observations without listwise deletion and provide unbiased estimates when data are missing at random.

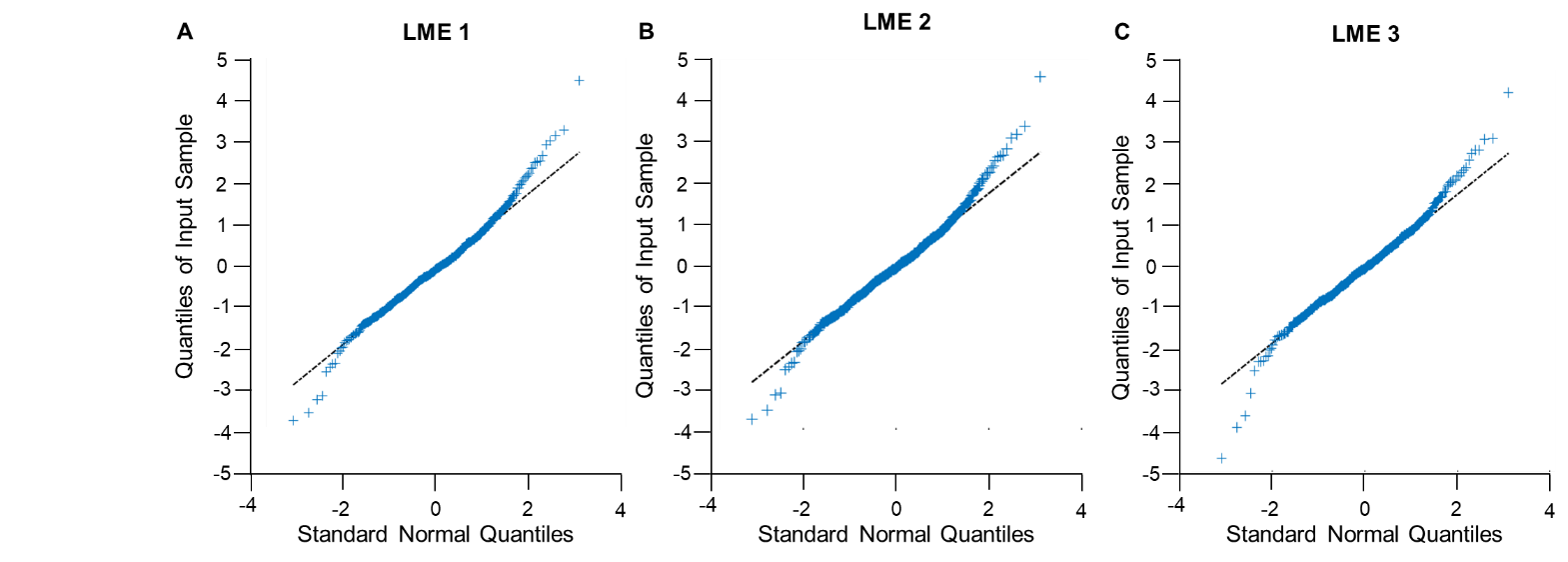

**Figure S4. Quantile–Quantile (Q–Q) plots of standardized residuals for LME1 to LME3.** These plots show the distribution of standardized residuals from the three longitudinal mixed-effects models assessing the relationship between therapeutic expectations, dissociative symptoms, and changes in depressive symptom severity (*MADRS*) across the ketamine treatment course. Model specifications were as follows: **(A)** LME1: *MADRS ~ time x EXPECT x CADSS + covariates*; **(B)** LME2: *MADRS ~ time x EXPECT + covariates*; **(C)** LME3: *MADRS ~ time x CADSS + covariates*. All models included random intercepts and slopes for time and were adjusted for age, sex, and education. Each panel displays standardized residuals (blue points) plotted against the theoretical quantiles of a standard normal distribution. The black diagonal line indicates the expected distribution under normality. Residuals closely follow the theoretical line across all models, supporting the assumption of normally distributed residuals without systematic deviation or influential data points.

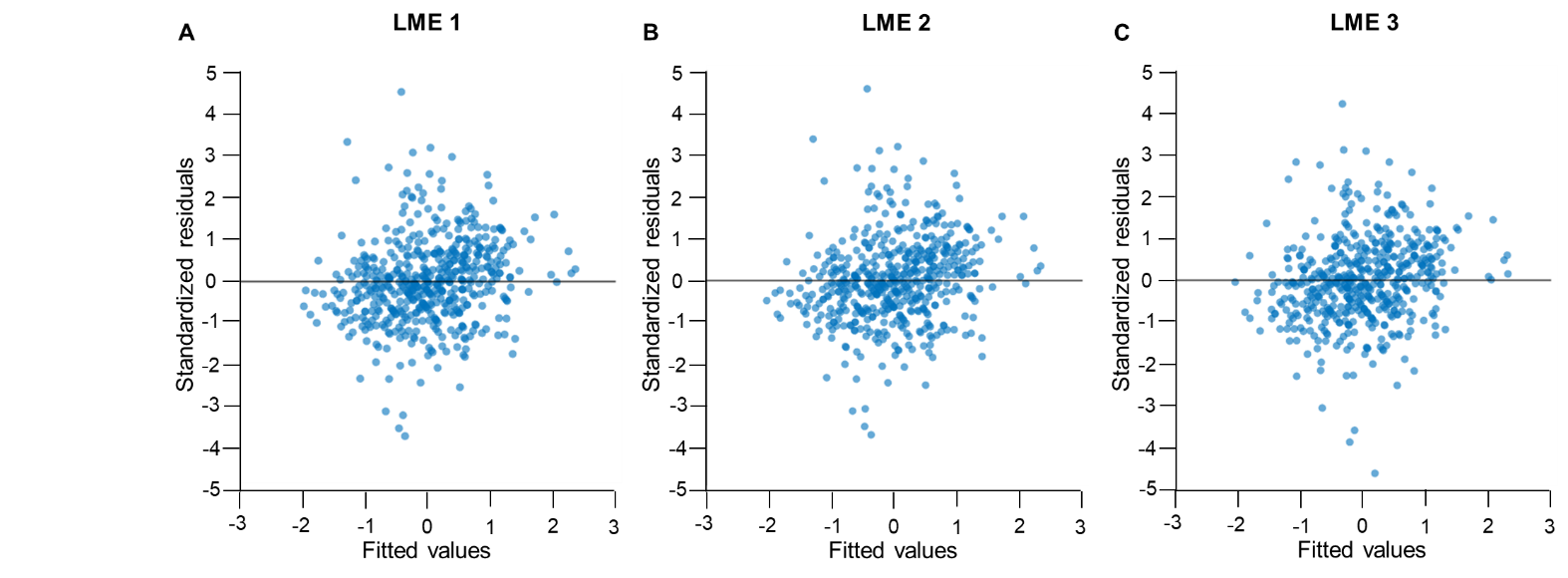

**Figure S5. Residuals versus Fitted Values for Longitudinal LME1 to LME3.** These plots display standardized residuals as a function of fitted values for the three longitudinal mixed-effects models assessing changes in depressive symptoms (*MADRS*) throughout the ketamine treatment course. Model specifications were as follows: **(A)** LME1: *MADRS ~ time x EXPECT x CADSS + covariates*; **(B)** LME2: *MADRS ~ time x EXPECT + covariates*; **(C)** LME3: *MADRS ~ time x CADSS + covariates.* All models included random intercepts and slopes for time, adjusted for age, sex, and education. Standardized residuals (y-axis) are plotted against fitted values (x-axis), with the horizontal black line indicating the zero reference. Residuals were symmetrically distributed around zero with consistent variance across fitted values, indicating homoscedasticity and supporting that model assumptions were met.

**Model Fit and Diagnostics for Longitudinal Analysis of the Trajectory of Depressive Symptoms (LME1–LME3)**

Model performance was evaluated using the Akaike Information Criterion (AIC), the Bayesian Information Criterion (BIC), and log-likelihood values. Fit indices for the final models were:

- **LME1** (Full model: Expectation x Dissociation x Time): *AIC* = 1195.1, *BIC* = 1259.0, *LogLik* = -582.5;
- **LME2** (Expectation x Time): *AIC* = 1189.0, *BIC* = 1235.8, *LogLik* = -583.5;
- **LME3** (Dissociation x Time): *AIC* = 1230.1, *BIC* = 1276.9, *LogLik* = -604.0.

Each model included 524 valid observations (n = 100 patients) and converged successfully. Goodness-of-fit was quantified using marginal R² (variance explained by fixed effects), conditional R² (variance explained by fixed and random effects), and the root mean square error (RMSE):

**•	LME1:** *R²_marginal_* = 0.21, *R²_conditional_* = 0.62, *RMSE* = 0.52

**•	LME2:** *R²_marginal_* = 0.20, *R²_conditional_* = 0.62, *RMSE* = 0.52

**•	LME3:** *R²_marginal_* = 0.17, *R²_conditional_* = 0.63, *RMSE* = 0.48

In addition to the total CADSS dissociation score, the same modeling approach was applied independently to the three CADSS subdimensions—depersonalization, derealization, and amnesia—each analyzed in parallel mixed-effects models with identical structure and covariate adjustment. Fixed-effect estimates, standard errors, and 95% confidence intervals for each model are reported in Tables S1–S3 in the Supplement.

**Methods S9**. Cross-Validation of LME1

To evaluate model generalizability, we used 10-fold group-aware cross-validation, ensuring that all observations from the same participant were placed in the same fold. Patients were randomly divided into ten folds. The model was trained on nine folds and tested on the remaining fold, repeated for all folds. This method offers an empirical estimate of out-of-sample predictive performance while reducing bias from repeated measures or effects of individual participants. Predictive accuracy was measured using root mean squared error (*RMSE*) and the coefficient of determination *R²*, calculated for both training and testing datasets in each fold.

The linear mixed-effects model predicting depressive symptom severity (MADRS) showed good internal fit, with an in-sample *RMSE* of 0.52 and total *R²* of 0.73 (*SD* = 0.01), including *R²_marginal_* = 0.20 and *R²_conditional_* = 0.61. The average test *RMSE* was 0.88 (*SD* = 0.14), with a mean *R²* of 0.19 (*SD* = 0.09). The average difference in RMSE between training and test sets was 0.36 (*SD* = 0.13), indicating a drop in predictive accuracy for unseen patients.

When expressed on the original MADRS scale (*SD* = 9.52), the in-sample RMSE corresponded to 4.96 MADRS points, and the cross-validated test RMSE corresponded to 8.38 MADRS points. The mean difference between training and test performance was 3.42 MADRS points (Figure S8). In standardized units, prediction errors corresponded to approximately 0.52 standard deviations in the training data and 0.88 standard deviations in cross-validation. This pattern is consistent with the known interindividual heterogeneity in clinical response to antidepressant treatments and suggests that the model captured within-subject variance more effectively than between-subject variance.

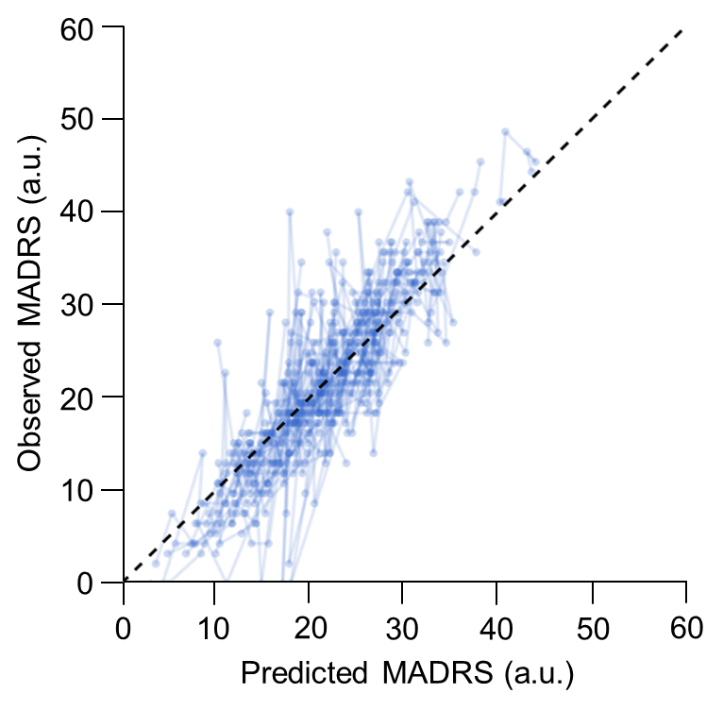

**Figure S6. Correlation of Predicted MADRS Scores from LME1 and Observed MADRS Scores.** This figure shows the relationship between observed and predicted Montgomery-Åsberg Depression Rating Scale (MADRS) scores among patients (n = 100). Each point represents a patient’s observed and model-predicted MADRS values at a given-assessment. The dashed diagonal line indicates perfect agreement between observed and predicted scores. Predicted values were generated from a mixed-effects model using 10-fold group-aware cross-validation and are shown on the original MADRS scale.

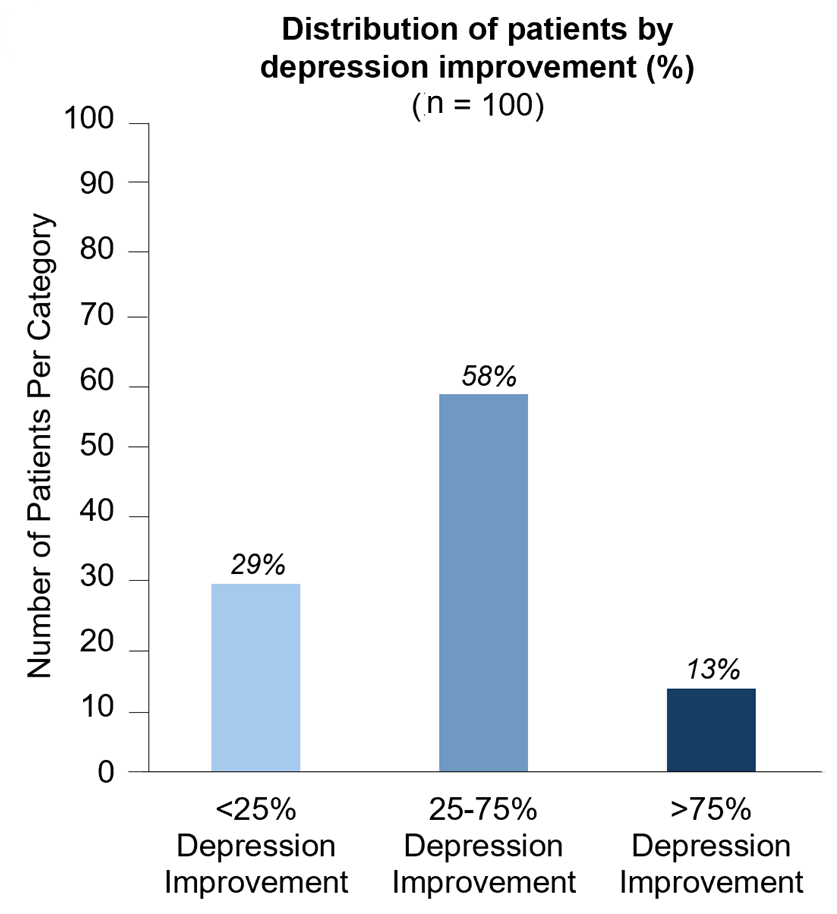

**Figure S7**. **Distribution of treatment responses.** percentages of patients in each of the three MADRS-based improvement categories.

**Methods S10.** Overall Model Fit Diagnostics LME4 and LME5

Complementary analyses examined whether therapeutic expectations and dissociative symptoms evolved differently across levels of clinical response. Patients were categorized as low responders (<25% reduction in MADRS), moderate responders (25–75%), or high responders (≥75%). Two separate models were fitted using restricted maximum likelihood (REML). Fit indices were:

- LME4 (*EXPECT* ~ *time* x *group*): *AIC* = 1528.4, *BIC* = 1592.6, *LogLik* = 749.2.
- LME5 (*CADSS* ~ *time* x *group*): *AIC* = 1572.7, *BIC* = 1636.8, *LogLik* = 771.4.

Each model included 624 valid observations (n = 100 patients), corresponding to repeated measures across six treatment sessions. Missing data were handled under a working assumption that data were missing at random (MAR) within the likelihood-based mixed-model framework, which uses all available observations without listwise deletion.

Goodness-of-fit indices were:

- **LME4:** *R²_marginal_* = 0.19, *R²_conditional_* = 0.64, *RMSE* = 0.48.
- **LME5:** *R²_marginal_* = 0.17, *R²_conditional_* = 0.63, *RMSE* = 0.48.

As with the total CADSS score, the same dimensional analyses were conducted for the three CADSS subdimensions—depersonalization, derealization, and amnesia—using the same model structure, covariate set, and estimation procedure. Fixed-effect estimates, standard errors, and 95% confidence intervals are presented in Tables S6 and S7 in the Supplement.

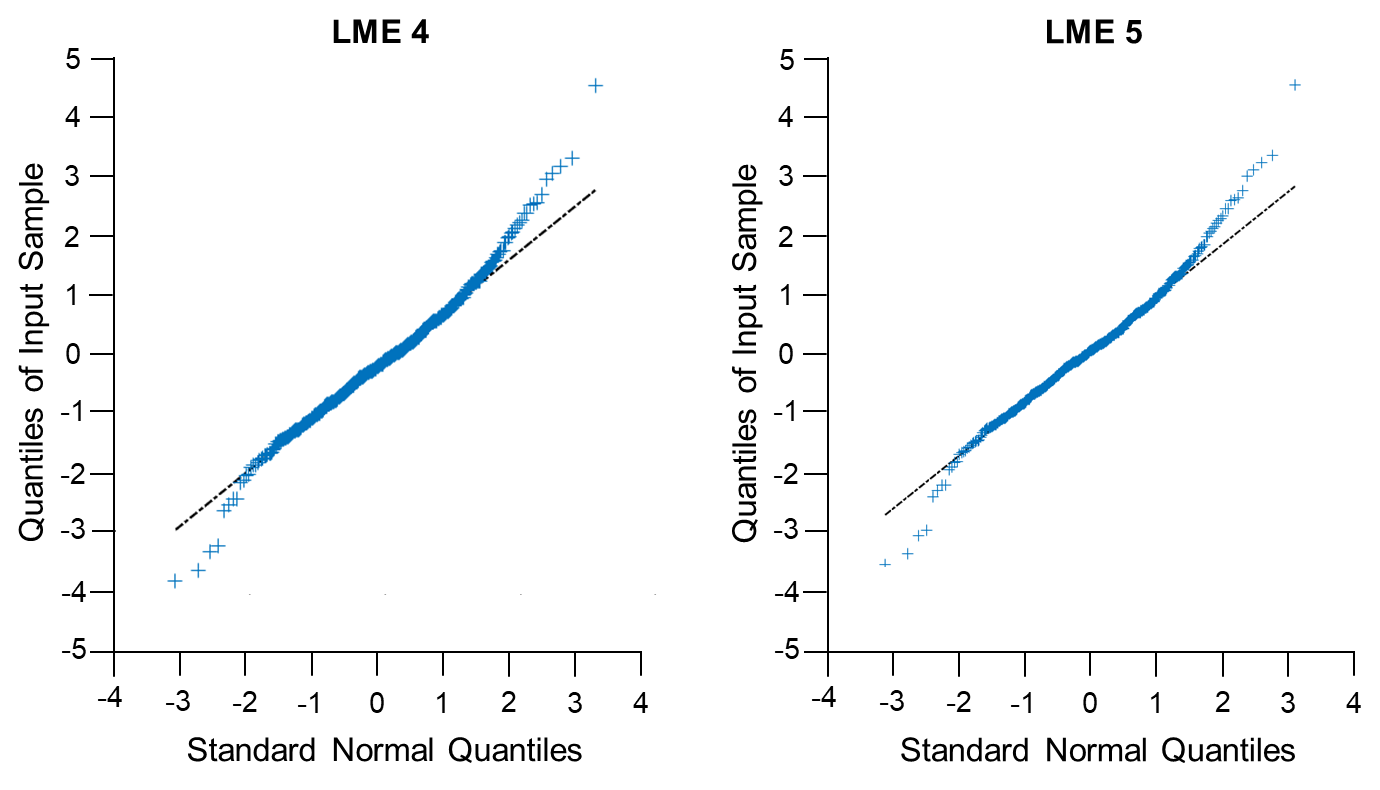

**Figure S8. Quantile–Quantile (Q–Q) plots of standardized residuals for LME4 and LME5.** These plots show the distribution of standardized residuals from the two longitudinal mixed-effects models examining the relationship between depression improvement, therapeutic expectations, and dissociative symptoms over time. Model specifications are as follows: **(A)** LME4: *EXPECT* ~ *time* x *group* + *covariates*; **(B)** LME5: *CADSS* ~ *time* x *group* + *covariates*. All models included random intercepts and random slopes for time and were adjusted for age, sex, and education. Each panel displays standardized residuals (blue points) plotted against the theoretical quantiles of a standard normal distribution. The black diagonal line indicates the expected distribution under normality.

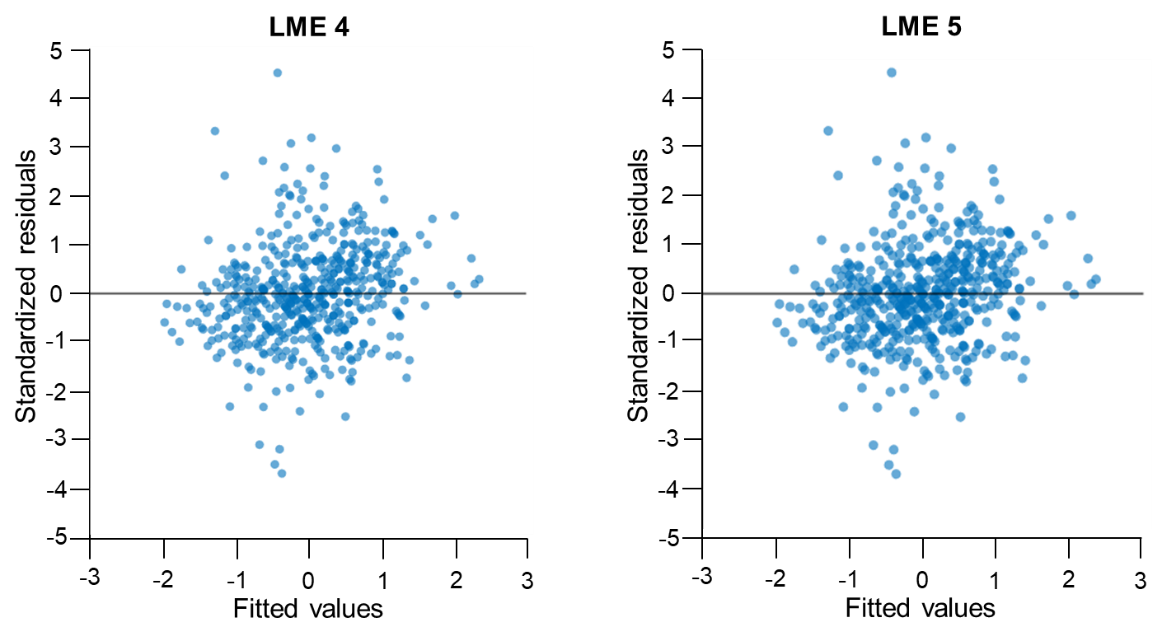

**Figure S9. Residuals versus Fitted Values for LME4 and 5.** These plots show standardized residuals plotted against fitted values for the two longitudinal mixed-effects models examining the relationship between depression improvement, therapeutic expectations, and dissociative symptoms during the ketamine treatment course. Model specifications were as follows: **(A)** LME4: *EXPECT* ~ *time* x *group* + *covariates*; **(B)** LME5: *CADSS* ~ *time* x *group* + *covariates*. All models included random intercepts and slopes for time and were adjusted for age, sex, and education. Standardized residuals (y-axis) are plotted against fitted values (x-axis), with the horizontal black line indicating zero. Residuals were symmetrically distributed around zero with relatively constant variance across fitted values, indicating homoscedasticity and supporting that the model assumptions were adequately met.

**Methods S11.** Random Intercept Cross-Lagged Panel Model (RI-CLPM) Specification

**Rationale for the RI-CLPM**

The standard cross-lagged panel model (CLPM) estimates cross-lagged effects from pooled variance that combines overall between-person differences and within-person fluctuations over time. When individual differences are present, as expected for therapeutic expectations, dissociative symptom propensity, and depressive symptom severity, CLPM estimates cannot be interpreted as purely within-person dynamic effects. The RI-CLPM (10) addresses this limitation by introducing a random intercept for each variable, thereby separating overall between-person variance from session-specific within-person deviations. Cross-lagged and autoregressive paths are then estimated on the within-person (person-mean centered) component (11).

**Model specification**

Each observed score at session T_n_ was decomposed into three components: (i) a between-person random intercept, representing each individual’s average level across all sessions; (ii) a within-person deviation around that individual mean; and (iii) residual variance capturing unexplained within-person variability. All cross-lagged and autoregressive paths were estimated on the within-person deviations. Random intercepts were allowed to covary freely across variables to capture overall between-person associations among variables.

Three models were specified to address distinct questions:

(1) **RI-CLPM1** tested whether therapeutic expectations predict both dissociative symptoms (*β_ec_*) and depressive symptom severity (*β_ed_*) within each session, and whether dissociative symptoms additionally predict depressive symptom severity (*β_cd_*) beyond expectations, while controlling for reverse temporal paths (*β_de_, β_ce_, β_dc_*):

$$\left\{ \begin{aligned} \text{CADSS}(T_{n})=\beta_{ec}\cdot\text{Expectations }(T_{n})+\varepsilon\\ \text{MADRS}(T_{n})=\beta_{ed}\cdot\text{Expectations }(T_{n})+\beta_{cd}\cdot\text{CADSS}(T_{n})+\varepsilon\end{aligned} \right.$$

(2) **RI-CLPM2** tested the robustness of the expectation effect on depressive symptom severity (*β_ed_*) observed in RI-CLPM1, after removing dissociative symptoms from the model, with prior depressive symptom severity predicting subsequent therapeutic expectations (*β_de_*: MADRS→Expectations) as reverse path control:

$$\text{MADRS}\left( T_{n} \right)=\beta_{ed}\cdot\text{Expectations }\left( T_{n} \right)+\varepsilon$$

(3) **RI-CLPM3** assessed whether the absence of a significant effect of dissociative symptoms on depressive symptom severity (*β_cd_*) observed in RI-CLPM1 persisted after removing therapeutic expectations from the model, testing whether this null finding was not due to multicollinearity with therapeutic expectations:

$$\text{MADRS}\left( T_{n} \right)=\beta_{cd}\cdot\text{CADSS}\left( T_{n} \right)+\varepsilon$$

**Within-session directional paths (lag0)**

The assessment sequence was fixed: therapeutic expectations were assessed before infusion (EXPECT); post-infusion, dissociative symptoms were systematically assessed by the patient (CADSS) prior to depressive symptom severity rated by the clinician (MADRS), within a 1–4 hour post-infusion window once acute dissociative symptoms had clinically dissipated. Accordingly, contemporaneous within-session paths were specified as directional rather than bidirectional: *β_ec_* (EXPECT→CADSS), *β_ed_* (EXPECT→MADRS), and *β_cd_* (CADSS→MADRS), consistent with recent methodological recommendations for panel models with protocol-defined temporal ordering (12,13).

**Inter-session paths and constraints**

Inter-session paths captured associations from session T_n_ to session T_n+1_. Autoregressive paths represented temporal stability within each variable. Reverse paths, *β_de_* (MADRS→EXPECT), *β_ce_* (CADSS→EXPECT), and *β_dc_* (MADRS→CADSS), were included to account for the possibility that prior symptom levels influenced subsequent expectations or dissociative symptoms. Autoregressive paths and *β_dc_* were constrained to equality across transitions, whereas *β_de_* and *β_ce_* were freely estimated across transitions. Constraining autoregressive paths to equality is consistent with common RI-CLPM specifications imposing time-invariant lagged effects to improve parsimony and statistical power, and reflects an assumption of temporal stationarity of within-person processes (14). Reverse paths from depressive and dissociative symptoms to subsequent expectations (*β_de_*, *β_ce_*) were freely estimated to allow for potential dynamic feedback effects, as therapeutic expectations are shaped by prior symptom trajectories and treatment-related experiences (15–17). In contrast, the reverse path from depressive symptoms to subsequent dissociative symptoms (*β_dc_*) was constrained to equality across transitions, reflecting its role as a control parameter in the absence of specific theoretical or empirical indications of time-varying effects in this direction. This specification ensures model parsimony while preserving flexibility for theoretically plausible feedback mechanisms.

**Covariates**

Age, sex, and education were regressed on all random intercepts. Baseline MADRS was additionally regressed on the random intercept of depressive symptoms (RId) to account for overall between-person differences in baseline severity.

**Model identification and estimation**

Models were estimated in R using lavaan (version 0.6-21; (18)) with robust maximum likelihood (MLR; Yuan-Bentler correction) and full-information maximum likelihood for missing data under the missing-at-random assumption. Model fit was evaluated using standard fit indices, including the comparative fit index (CFI), root mean square error of approximation (RMSEA) with 90% confidence interval, Akaike information criterion (AIC), and Bayesian information criterion (BIC). CFI values ≥ 0.90 and RMSEA values ≤ 0.08 were considered indicative of adequate fit, with CFI ≥ 0.95 and RMSEA ≤ 0.05 reflecting excellent fit, consistent with conventional cutoffs in structural equation modeling (19,20). AIC and BIC were used for relative model comparison, with lower values indicating better-supported models (21).

**Directional hypothesis testing: GORICA**

Directional hypotheses were pre-specified on theoretical grounds prior to data analysis, based on the hypothesized mechanistic roles of therapeutic expectations and dissociative symptoms in ketamine's antidepressant action (8,22–26). Hypotheses were evaluated using the Generalized Order-Restricted Information Criterion Approximation (GORICA; (27)), an AIC-type information criterion that quantifies the relative support for order-restricted, theory-based hypotheses under structural equation models (28). The GORICA has been validated for RI-CLPM analyses through simulation, demonstrating adequate performance for evaluating directional dominance hypotheses among cross-lagged effects (29).

Each order-constrained hypothesis was evaluated against its complement, representing all alternative orderings, following Vanbrabant (30). This approach allows directional dominance to be evaluated in a manner that is both statistically coherent and substantively interpretable within the GORICA framework. For each hypothesis, a GORICA weight (w, reflecting relative support for the hypothesis compared with its complement, ranging from 0 to 1) and evidence ratio (w[H]/w[complement]) are reported. Loglik weights, which reflect empirical fit without the complexity penalty, are reported alongside GORICA weights to distinguish fit-based support from the parsimony-corrected evidence ratio (Tables S10–S12).

**Pre-specified directional hypotheses**

Directional dominance was tested on the absolute values of the path coefficients, to compare the relative magnitudes of opposing within-session and inter-session effects independently of their sign. This formulation is consistent with the causal dominance framework defined by Hamaker, Kuiper, and Grasman (2015) for the RI-CLPM (10), and has been validated through simulation for RI-CLPM applications (29).

Two hypotheses tested whether the hypothesized effects of therapeutic expectations exceeded their corresponding reverse effects, thereby evaluating directional dominance in the temporal ordering of effects:

*(i)* |*β_ed_|* > |*β_de_|* across all sessions, the effect of therapeutic expectations on depressive symptoms dominates the reverse effect;

*(ii)* |*β_ec_|* > |*β_ce_|* across all session-to-session transitions, the effect of therapeutic expectations on dissociative symptoms dominates the reverse effect.

Hypothesis (i) was tested in RI-CLPM1 and RI-CLPM2; hypothesis (ii) was tested in RI-CLPM1.

**Interpretability criterion**

The confirmatory interpretation of GORICA results is strongest when a genuine directional effect exists at the within-person level. As demonstrated through simulation by Kuiper (28), the GORIC and GORICA asymptotically select the correct inequality-constrained hypothesis when a true directional effect is present. When the directional effect is null or inconsistent, the GORICA weight may be driven more by the penalty structure than by empirical directional signal, and should therefore not be interpreted as confirmatory evidence. Accordingly, interpretation considered both GORICA weights and the consistency of coefficient directionality across sessions. The present sample size (n = 100) meets the minimum sample size recommended for GORICA applications to structural equation models (27).

**Table S1.** Longitudinal Analysis: Linear Mixed-Effects Model 1 of Depression

| **Predictor** | **β** | | **SE** | | **t** | | **P** | | | **95%CI** | | |
| --- | --- | --- | --- | --- | --- | --- | --- | --- | --- | --- | --- | --- |
|  |  |  |  |  |  |  |  |  |  | **Lower** | | **Upper** |
| **Dissociative symptoms** – **Total CADSS score** | | | | | | | | | | | | |
| Intercept | -0.02 | | 0.09 | | -0.28 | | .7804 | | | -0.19 | | 0.15 |
| Time | -0.27 | | 0.05 | | -5.83 | | **9.5787e-09** | | | -0.36 | | -0.18 |
| EXPECT | -0.32 | | 0.05 | | -6.67 | | **6.4548e-11** | | | -0.42 | | -0.23 |
| CADSS | -0.06 | | 0.05 | | -1.29 | | .1972 | | | -0.15 | | 0.03 |
| Time x EXPECT | -0.06 | | 0.03 | | -1.64 | | .1014 | | | -0.13 | | 0.01 |
| Time x CADSS | -0.02 | | 0.04 | | -0.55 | | .5799 | | | -0.09 | | 0.05 |
| EXPECT x CADSS | 0.00 | | 0.04 | | 0.10 | | .9211 | | | -0.08 | | 0.08 |
| Time x EXPECT x CADSS | 0.00 | | 0.04 | | 0.03 | | .9800 | | | -0.07 | | 0.08 |
| Age | -0.02 | | 0.05 | | -0.49 | | .6247 | | | -0.11 | | 0.07 |
| Sex | 0.08 | | 0.14 | | 0.58 | | .5594 | | | -0.19 | | 0.36 |
| Study | -0.00 | | 0.05 | | -0.09 | | .9288 | | | -0.09 | | 0.09 |
| **Depersonalization symptoms** | | | | | | | | | | | | |
| Intercept | -0.02 | | 0.09 | | -0.19 | | .8467 | | | -0.19 | | 0.15 |
| Time | -0.27 | | 0.05 | | -5.81 | | **1.1165e-08** | | | -0.36 | | -0.18 |
| EXPECT | -0.32 | | 0.05 | | -6.52 | | **1.7274e-10** | | | -0.41 | | -0.22 |
| Depersonalization | -0.08 | | 0.05 | | -1.74 | | .0827 | | | -0.17 | | 0.01 |
| EXPECT x Time | -0.05 | | 0.03 | | -1.59 | | .1118 | | | -0.12 | | 0.01 |
| Time * Depersonalization | -0.03 | | 0.03 | | -0.90 | | .3696 | | | -0.10 | | 0.04 |
| EXPECT x Depersonalization | -0.02 | | 0.04 | | -0.53 | | .5966 | | | -0.10 | | 0.06 |
| Time x EXPECT x Depersonalization | 0.01 | | 0.04 | | 0.33 | | .7440 | | | -0.06 | | 0.08 |
| Age | -0.02 | | 0.05 | | -0.46 | | .6438 | | | -0.11 | | 0.07 |
| Sex | 0.08 | | 0.14 | | 0.56 | | .5733 | | | -0.20 | | 0.36 |
| Study | -0.00 | | 0.05 | | -0.07 | | .9412 | | | -0.09 | | 0.09 |
| **Derealization manifestations** | | | | | | | | | | | | |
| Intercept | | -0.02 | | 0.09 | | -0.28 | | .7831 | -0.19 | | 0.15 | |
| Time | | -0.27 | | 0.05 | | -5.80 | | **1.1313e-08** | -0.36 | | -0.18 | |
| EXPECT | | -0.33 | | 0.05 | | -6.86 | | **1.9771e-11** | -0.43 | | -0.24 | |
| Derealization | | -0.05 | | 0.05 | | -1.15 | | .2522 | -0.14 | | 0.04 | |
| EXPECT x Time | | -0.06 | | 0.04 | | -1.61 | | .1084 | -0.13 | | 0.01 | |
| Time x Derealization | | -0.01 | | 0.04 | | -0.30 | | .7639 | -0.08 | | 0.06 | |
| EXPECT x Derealization | | 0.01 | | 0.04 | | 0.18 | | .8606 | -0.07 | | 0.09 | |
| Time x EXPECT x Derealization | | -0.02 | | 0.04 | | -0.52 | | .6036 | -0.09 | | 0.05 | |
| Age | | -0.02 | | 0.05 | | -0.48 | | .6291 | -0.11 | | 0.07 | |
| Sex | | 0.08 | | 0.14 | | 0.56 | | .5779 | -0.20 | | 0.36 | |
| Study | | -0.00 | | 0.05 | | -0.10 | | .9239 | -0.09 | | 0.09 | |
| **Amnesia manifestations** | | | | | | | | | | | | |
| Intercept | | -0.01 | | 0.09 | | -0.15 | | .8769 | -0.18 | | 0.16 | |
| Time | | -0.27 | | 0.05 | | -5.88 | | **7.3868e-09** | -0.36 | | -0.18 | |
| EXPECT | | -0.35 | | 0.05 | | -7.19 | | **2.3017e-12** | -0.44 | | -0.25 | |
| Amnesia | | 0.00 | | 0.04 | | 0.10 | | .9180 | -0.08 | | 0.09 | |
| EXPECT x Time | | -0.06 | | 0.03 | | -1.66 | | .0978 | -0.13 | | 0.01 | |
| Time x Amnesia | | 0.01 | | 0.03 | | 0.24 | | .8117 | -0.06 | | 0.07 | |
| EXPECT x Amnesia | | 0.06 | | 0.04 | | 1.43 | | .1521 | -0.02 | | 0.13 | |
| Time x EXPECT x Amnesia | | 0.03 | | 0.03 | | 0.83 | | .4068 | -0.04 | | 0.09 | |
| Age | | -0.02 | | 0.05 | | -0.45 | | .6511 | -0.11 | | 0.07 | |
| Sex | | 0.05 | | 0.14 | | 0.36 | | .7176 | -0.23 | | 0.33 | |
| Study | | -0.01 | | 0.05 | | -0.13 | | .8974 | -0.10 | | 0.08 | |

**LME1 model fit**: *AIC* = 1195.1, *BIC* = 1259.0, *LogLik* = -582.5; *R²_marginal_* = 0.21, *R²_conditional_* = 0.62, *RMSE* = 0.52. **Random effects**: *SD(intercept)* = 0.61 [0.52 to 0.73], *SD(time)* = 0.20 [0.13 to 0.30], *Residual(SD)* = 0.59 [0.55 to 0.64].

**LME1_depersonalization_ model fit**: *AIC* = 1192.9, *BIC* = 1256.8, *LogLik* = -581.5; *R²_marginal_* = 0.21, *R²_conditional_* = 0.62, *RMSE* = 0.52. **Random effects**: *SD(intercept)* = 0.62 [0.52 to 0.73], *SD(time)* = 0.19 [0.12 to 0.30], *Residual(SD)* = 0.59 [0.55 to 0.64].

**LME1_depersonalization_ model fit**: *AIC* = 1195.3, *BIC* = 1259.2, *LogLik* = -582.7; *R²_marginal_* = 0.21, *R²_conditional_* = 0.619, *RMSE* = 0.52. **Random effects**: *SD(intercept)* = 0.61 [0.51 to 0.73], *SD(time)* = 0.20 [0.14 to 0.31], *Residual(SD)* = 0.59 [0.55 to 0.64].

**LME1_amnesia_** **model fit**: *AIC* = 1193.9, *BIC* = 1257.8, *LogLik* = -582.0; *R²_marginal_* = 0.21, *R²_conditional_* = 0.63, *RMSE* = 0.517. **Random effects**: *SD(intercept)* = 0.62 [0.52 to 0.74], *SD(time)* = 0.20 [0.14 to 0.31], *Residual(SD)* = 0.59 [0.54 to 0.63].

β represents regression coefficients; SE indicates standard error; CI, confidence interval; AIC, Akaike information criterion; BIC, Bayesian information criterion; LogLik, log-likelihood; R²_marginal,_ proportion of variance explained by fixed effects; R²_conditional_, proportion of variance explained by fixed and random effects; RMSE, Root-Mean-Square Error; SD, standard Deviation; z, standardized value; CADSS, Clinician-Administered Dissociative States Scale; MADRS, Montgomery-Åsberg Depression Rating Scale.

**Table S2**. Longitudinal Analysis: Linear Mixed-Effects Model 2 of Depression

| **Predictor** | **β** | **SE** | **t** | **P** | **95%CI** | |
| --- | --- | --- | --- | --- | --- | --- |
|  |  |  |  |  | **Lower** | **Upper** |
| Intercept | -0.02 | 0.09 | -0.20 | .8448 | -0.19 | 0.15 |
| Time | -0.27 | 0.05 | -5.90 | **6.4087e-09** | -0.36 | -0.18 |
| EXPECT | -0.34 | 0.05 | -7.07 | **5.0504e-12** | -0.43 | -0.24 |
| EXPECT * Time | -0.05 | 0.03 | -1.60 | .1103 | -0.12 | 0.01 |
| Age | -0.02 | 0.05 | -0.39 | .6986 | -0.11 | 0.07 |
| Sex | 0.06 | 0.14 | 0.46 | .6493 | -0.21 | 0.34 |
| Study | -0.01 | 0.05 | -0.16 | .8723 | -0.10 | 0.08 |

**LME2** **model fit**: *AIC* = 1189.0, *BIC* = 1235.8, *LogLik* = -583.5; *R²_marginal_* = 0.20, *R²_conditional_* = 0.62, *RMSE* = 0.52. **Random effects**: *SD(intercept)* = 0.62 [0.52 to 0.74], *SD(time)* = 0.20 [0.13 to 0.30], *Residual(SD)* = 0.59 [0.55 to 0.64].

**Table S3**. Longitudinal Analysis: Linear Mixed-Effects Model 3 of Depression

| **Predictor** | **β** | | **SE** | | **t** | **P** | | | **95%CI** | | |
| --- | --- | --- | --- | --- | --- | --- | --- | --- | --- | --- | --- |
|  |  |  |  |  |  |  |  |  | **Lower** | | **Upper** |
| **Dissociative symptoms** – **Total CADSS score** | | | | | | | | | | | |
| Intercept | -0.07 | | 0.10 | | -0.70 | .4864 | | | -0.26 | | 0.12 |
| Time | -0.25 | | 0.05 | | -5.26 | **2.0691e-07** | | | -0.34 | | -0.15 |
| CADSS | -0.11 | | 0.05 | | -2.20 | **.0279** | | | -0.20 | | -0.01 |
| Time x CADSS | -0.04 | | 0.03 | | -1.05 | .2954 | | | -0.10 | | 0.03 |
| Age | -0.04 | | 0.05 | | -0.92 | .3579 | | | -0.14 | | 0.05 |
| Sex | 0.22 | | 0.16 | | 1.40 | .1623 | | | -0.09 | | 0.52 |
| Study | 0.03 | | 0.05 | | 0.55 | .5816 | | | -0.07 | | 0.12 |
| **Depersonalization symptoms** | | | | | | | | | | | |
| Intercept | -0.06 | | 0.10 | | -0.64 | .5195 | | | -0.25 | | 0.13 |
| Time | -0.24 | | 0.05 | | -5.28 | **1.9292e-07** | | | -0.33 | | -0.15 |
| Depersonalization | -0.13 | | 0.05 | | -2.84 | **.0048** | | | -0.22 | | -0.04 |
| Time x Depersonalization | -0.06 | | 0.03 | | -1.64 | .1026 | | | -0.12 | | 0.01 |
| Age | -0.04 | | 0.05 | | -0.92 | .3570 | | | -0.14 | | 0.05 |
| Sex | 0.21 | | 0.16 | | 1.32 | .1872 | | | -0.10 | | 0.51 |
| Study | 0.03 | | 0.05 | | 0.57 | .5689 | | | -0.07 | | 0.12 |
| **Derealization manifestations** | | | | | | | | | | | |
| Intercept | -0.07 | | 0.10 | | -0.70 | .4830 | | | -0.26 | | 0.12 |
| Time | -0.25 | | 0.05 | | -5.29 | **1.7743e-07** | | | -0.34 | | -0.16 |
| Derealization | -0.08 | | 0.05 | | -1.65 | .1001 | | | -0.17 | | 0.02 |
| Time x Derealization | -0.02 | | 0.03 | | -0.65 | .5165 | | | -0.09 | | 0.05 |
| Age | -0.04 | | 0.05 | | -0.86 | .3911 | | | -0.13 | | 0.05 |
| Sex | 0.22 | | 0.16 | | 1.39 | .1649 | | | -0.09 | | 0.52 |
| Study | 0.02 | | 0.05 | | 0.52 | .6044 | | | -0.07 | | 0.12 |
| **Amnesia manifestations** | | | | | | | | | | | |
| Intercept | -0.06 | 0.10 | | -0.64 | | | .5218 | -0.26 | | 0.13 | |
| Time | -0.25 | 0.05 | | -5.26 | | | **2.1544e-07** | -0.34 | | -0.15 | |
| Amnesia | -0.02 | 0.04 | | -0.41 | | | .6823 | -0.11 | | 0.07 | |
| Time x Amnesia | 0.01 | 0.03 | | 0.18 | | | .8564 | -0.06 | | 0.07 | |
| Age | -0.04 | 0.05 | | -0.80 | | | .4214 | -0.13 | | 0.05 | |
| Sex | 0.20 | 0.16 | | 1.28 | | | .1999 | -0.11 | | 0.52 | |
| Study | 0.02 | 0.05 | | 0.48 | | | .6336 | -0.07 | | 0.11 | |

**LME3** **model fit**: *AIC* = 1230.1, *BIC* = 1276.9, *LogLik* = -604.0; *R²_marginal_* = 0.08, *R²_conditional_* = 0.62, *RMSE* = 0.54. **Random effects**: *SD(intercept)* = 0.72 [0.61 to 0.85], *SD(time)* = 0.19 [0.12 to 0.30], *Residual(SD)* = 0.61 [0.56 to 0.66].

**LME3_depersonalization_** **model fit:** *AIC* = 1226.1, *BIC* = 1272.9, *LogLik* = -602.0; *R²_marginal_* = 0.09, *R²_conditional_* = 0.62, *RMSE* = 0.54. **Random-effects**: *SD(intercept)* = 0.72 [0.61 to 0.84], *SD(time)* = 0.18 [0.10 to 0.30], and *Residual(SD)* = 0.61 [0.56 to 0.66].

**LME3_derealization_** **model fit:** *AIC* = 1232.6, *BIC* = 1279.5, *LogLik* = -605.3; *R²_marginal_* = 0.08, *R²_conditional_* = 0.61, *RMSE* = 0.54. **Random-effects**: *SD(intercept)* = 0.72 [0.61 to 0.85], *SD(time)* = 0.20 [0.12 to 0.30], and *Residual(SD)* = 0.61 [0.56 to 0.66].

**LME3_amnesia_** **model fit:** *AIC* = 1235.2, *BIC* = 1282.0, *LogLik* = -606.6; *R²_marginal_* = 0.07, *R²_conditional_* = 0.62, *RMSE* = 0.54. **Random-effects**: *SD(intercept)* = 0.74 [0.63 to 0.87], *SD(time)* = 0.19 [0.12 to 0.31], and *Residual(SD)* = 0.61 [0.56 to 0.66].

**Table S4**. Session-Level Analysis: Linear Regression Model of Depression Predicted by Therapeutic Expectations and Dissociative Symptoms

| **Independant Variable** | **Time** | **β** | **SE** | **P** | **%Δ MADRS** | **R²** |
| --- | --- | --- | --- | --- | --- | --- |
| **Therapeutic expectations**  –  **EXPECT**  **Total score** | T0 | 9.01 | 2.70 | **.0012** | 20.35 | 0.19 |
|  | T1 | 10.73 | 2.71 | **.0001** | 24.38 | 0.22 |
|  | T2 | 10.94 | 2.74 | **.0001** | 24.69 | 0.23 |
|  | T3 | 13.79 | 2.64 | **1.2165e-06** | 30.70 | 0.32 |
|  | T4 | 15.41 | 2.75 | **2.7142e-07** | 34.04 | 0.34 |
|  | T5 | 14.82 | 2.95 | **3.4748e-06** | 31.34 | 0.29 |
|  | T6 | 15.86 | 3.00 | **1.4328e-06** | 34.29 | 0.36 |
| **Dissociative symptoms**  –  **CADSS Total**  **score** | T1 | 8.68 | 2.69 | **.0017** | 19.72 | 0.18 |
|  | T2 | 6.09 | 2.81 | .0330 | 13.76 | 0.14 |
|  | T3 | 9.94 | 2.65 | **.0003** | 22.13 | 0.23 |
|  | T4 | 8.02 | 2.93 | .0076 | 17.72 | 0.16 |
|  | T5 | 7.85 | 3.17 | .0155 | 16.61 | 0.12 |
|  | T6 | 7.35 | 3.15 | .0227 | 15.89 | 0.17 |

ΔMADRS% = % improvement per 1 SD increase in EXPECT (*depersonalization, derealization, amnesia*). Bonferroni (α = .0038) corrections were applied.

**Table S5**. Session-Level Analysis: Linear Regression Model of Depression Predicted by Therapeutic Expectations and Dissociative Symptom Dimensions

| **Independant Variable** | **Time** | **β** | **SE** | **P** | **%Δ MADRS** | **R²** |
| --- | --- | --- | --- | --- | --- | --- |
| **Depersonalization**  **manifestations** | T1 | 8.60 | 2.63 | **.0015** | 19.54 | 0.16 |
|  | T2 | 9.55 | 2.67 | **.0006** | 21.56 | 0.17 |
|  | T3 | 11.44 | 2.62 | **3.3676e-05** | 25.47 | 0.24 |
|  | T4 | 9.51 | 2.87 | **.0013** | 21.02 | 0.16 |
|  | T5 | 8.70 | 3.10 | .0064 | 18.39 | 0.12 |
|  | T6 | 10.37 | 3.12 | **.0014** | 22.41 | 0.17 |
| **Derealization**  **manifestations** | T1 | 8.32 | 2.67 | **.0024** | 18.91 | 0.15 |
|  | T2 | 6.31 | 2.76 | .0248 | 14.24 | 0.11 |
|  | T3 | 8.95 | 2.73 | **.0015** | 19.93 | 0.18 |
|  | T4 | 6.55 | 2.99 | .0311 | 14.48 | 0.10 |
|  | T5 | 8.57 | 3.14 | .0079 | 18.11 | 0.11 |
|  | T6 | 7.96 | 3.29 | .0182 | 17.20 | 0.11 |
| **Amnesia**  **symptoms** | T1 | 4.25 | 2.77 | .1287 | 9.65 | 0.09 |
|  | T2 | 0.63 | 2.85 | .8261 | 1.42 | 0.06 |
|  | T3 | 1.89 | 2.88 | .5133 | 4.21 | 0.08 |
|  | T4 | 0.94 | 3.09 | .7608 | 2.09 | 0.05 |
|  | T5 | 1.53 | 3.28 | .6427 | 3.23 | 0.02 |
|  | T6 | -1.05 | 3.37 | .7574 | -2.26 | 0.03 |

ΔMADRS% = % improvement per 1 SD increase in EXPECT (*depersonalization, derealization, amnesia*). Bonferroni (α = .0028) corrections were applied.

**Table S6.** Dimensional Analysis: Linear Mixed-Effects Model 4 of Therapeutic Expectations

| **Predictor** | **β** | **SE** | **t** | **P** | **95%CI** | |
| --- | --- | --- | --- | --- | --- | --- |
|  |  |  |  |  | **Lower** | **Upper** |
| Intercept | -0.03 | 0.08 | -0.42 | .6725 | -0.19 | 0.12 |
| Time | -0.05 | 0.04 | -1.31 | .1899 | -0.14 | 0.03 |
| Improvement Group | 0.65 | 0.12 | 5.25 | **2.0563e-07** | 0.41 | 0.89 |
| Time x Improvement Group | 0.19 | 0.06 | 3.21 | **.0014** | 0.07 | 0.30 |
| Age | 0.04 | 0.03 | 1.25 | .2105 | -0.02 | 0.10 |
| Sex | 0.14 | 0.08 | 1.76 | .0796 | -0.02 | 0.29 |
| Study | -0.04 | 0.03 | -1.37 | .1705 | -0.10 | 0.02 |

**LME4 model fit**: *AIC* = 1211.9, *BIC* = 1260.5, *LogLik* = -594.9; *R²_marginal_* = 0.24, *R²_conditional_* = 0.84, *RMSE* = 0.38. **Random effects**: *SD(intercept)* = 0.74 [0.63 to 0.86], *SD(time)* = 0.31 [0.25 to 0.39], *Residual(SD)* = 0.44 [0.41 to 0.47].

**Table S7**. Dimensional Analysis: Linear Mixed-Effects Model 5 of Dissociative Symptoms

| **Predictor** | **β** | | **SE** | | | **t** | **P** | **95%CI** | | |
| --- | --- | --- | --- | --- | --- | --- | --- | --- | --- | --- |
|  |  |  |  |  |  |  |  | **Lower** | | **Upper** |
| **Dissociative symptoms** | | | | | | | | | | |
| Intercept | -0.08 | | 0.10 | | | -0.72 | .4711 | -0.28 | | 0.13 |
| Improvement Group | 0.48 | | 0.13 | | | 3.61 | **.0003** | 0.22 | | 0.75 |
| Time | -0.02 | | 0.04 | | | -0.55 | .5809 | -0.10 | | 0.06 |
| Improvement Group x Time | -0.06 | | 0.05 | | | -1.30 | .1933 | -0.16 | | 0.03 |
| Age | -0.10 | | 0.04 | | | -2.45 | .0145 | -0.18 | | -0.02 |
| Study | 0.05 | | 0.04 | | | 1.17 | .2430 | -0.03 | | 0.12 |
| Sex | 0.27 | | 0.17 | | | 1.60 | .1112 | -0.06 | | 0.61 |
| **Depersonalization manifestations** | | | | | | | | | | |
| Intercept | -0.04 | | 0.10 | | | -0.40 | .6882 | -0.24 | | 0.16 |
| Improvement Group | 0.57 | | 0.13 | | | 4.55 | **6.77e-06** | 0.33 | | 0.82 |
| Time | -0.01 | | 0.04 | | | -0.29 | .7726 | -0.10 | | 0.07 |
| Improvement Group x Time | -0.04 | | 0.05 | | | -0.66 | .5080 | -0.14 | | 0.07 |
| Age | -0.10 | | 0.04 | | | -2.41 | **.0163** | -0.19 | | -0.02 |
| Study | 0.05 | | 0.04 | | | 1.23 | .2206 | -0.03 | | 0.14 |
| Sex | 0.15 | | 0.16 | | | 0.90 | .3691 | -0.17 | | 0.47 |
| **Derealization manifestations** | | | | | | | | | | |
| Intercept | | -0.08 | | 0.11 | -0.79 | | .4294 | | -0.29 | -0.12 |
| Improvement Group | | 0.40 | | 0.13 | 2.98 | | **.0031** | | 0.14 | 0.67 |
| Time | | -0.05 | | 0.04 | -1.28 | | .2010 | | -0.13 | 0.03 |
| Improvement Group x Time | | -0.07 | | 0.05 | -1.44 | | .1512 | | -0.16 | 0.02 |
| Age | | -0.08 | | 0.04 | -1.92 | | .0550 | | -0.16 | 0.06 |
| Study | | 0.04 | | 0.04 | 0.95 | | .3444 | | -0.04 | 0.12 |
| Sex | | 0.31 | | 0.17 | 1.79 | | .0747 | | -0.03 | 0.64 |
| **Amnesia manifestations** | | | | | | | | | | |
| Intercept | | -0.07 | | 0.11 | -0.64 | | .5217 | | -0.28 | 0.14 |
| Improvement Group | | 0.13 | | 0.14 | 0.99 | | .3234 | | -0.13 | 0.40 |
| Time | | 0.07 | | 0.04 | 1.59 | | .1135 | | -0.02 | 0.15 |
| Improvement Group x Time | | -0.06 | | 0.05 | -1.33 | | .1848 | | -0.16 | 0.03 |
| Age | | -0.06 | | 0.05 | -1.32 | | .1886 | | -0.15 | 0.03 |
| Study | | 0.03 | | 0.05 | 0.58 | | .5619 | | -0.06 | 0.12 |
| Sex | | 0.21 | | 0.17 | 1.24 | | .2142 | | -0.12 | 0.55 |

**LME5 model fit**: *AIC* = 1120.9; *BIC* = 1167.7; *LogLik* = -549.5; *R²_marginal_* = 0.12, *R²_conditional_* = 0.78, *RMSE* = 0.44. **Random effects**: *SD(intercept)* = 0.79 [0.68 to 0.92], *SD(time)* = 0.22 [0.15 to 0.32], *Residual(SD)* = 0.51 [0.47 to 0.55].

**LME5_depersonalization_ model fit**: *AIC* = 1172.8; *BIC* = 1219.6; *LogLik* = -575.4; *R²_marginal_* = 0.16, *R²_conditional_* = 0.75, *RMSE* = 0.47. **Random effects**: *SD(intercept)* = 0.74 [0.63 to 0.86], *SD(time)* = 0.22 [0.15 to 0.33], *Residual(SD)* = 0.54 [0.50 to 0.58].

**LME5_derealization_ model fit**: *AIC* = 1143.6; *BIC* = 1190.3; *LogLik* = -560.8; *R²_marginal_* = 0.09, *R²_conditional_* = 0.75, *RMSE* = 0.46. **Random effects**: *SD(intercept)* = 0.79 [0.68 to 0.93], *SD(time)* = 0.16 [0.10 to 0.27], *Residual(SD)* = 0.53 [0.49 to 0.57].

**LME5_amnesia_ model fit**: *AIC* = 1234.6; *BIC* = 1281.4; *LogLik* = -606.3; *R²_marginal_* = 0.03, *R²_conditional_* = 0.67, *RMSE* = 0.53. **Random effects**: *SD(intercept)* = 0.79 [0.67 to 0.93], *SD(time)* = 0.13 [0.06 to 0.29], *Residual(SD)* = 0.60 [0.55 to 0.65].

**Table S8.** Three-Path Mediation Model with Average Therapeutic Expectations, and Dissociative Symptom Scores

| **Path** | **β** | **SE** | **t** | **Z** | **P** |
| --- | --- | --- | --- | --- | --- |
| a1  X: EXPECT_T0_ → M1: CADSS_T1_ | 0.20 | 0.09 | 2.14 | 2.05 | **.0404** |
| a2  M1: CADSS_T1_ →  M2: MADRS_T0–T1_ | 0.19 | 0.10 | 1.91 | 1.96 | **.0497** |
| b  M2: MADRS_T0–T1_→  Y: MADRS_T0–T6_ | 0.33 | 0.10 | 3.46 | 2.96 | **.0030** |
| c’ (direct) | 0.24 | 0.11 | 2.30 | 2.29 | **.0220** |
| c (total) | 0.39 | 0.09 | 4.27 | **3.54** | **.0004** |
| Indirect Effect  a1·a2·b | 0.01 | 0.01 | 1.23 | **2.09** | **.0365** |

Results from three-path mediation analysis testing whether early dissociative symptoms (M1) and early improvement (M2) sequentially mediated the association between therapeutic expectations (X) and final depression improvement (Y). CADSS indicates Clinician-Administered Dissociative States Scale; EXPECT, average therapeutic expectations score; and MADRS, Montgomery-Åsberg Depression Rating Scale.

**Table S9**. Three-Path Mediation Model with Dissociative Symptom Dimensions

| **Path** | **β** | **SE** | **t** | **Z** | **P** |
| --- | --- | --- | --- | --- | --- |
| **Depersonalization manifestations** | | | | | |
| a1  X: EXPECT_T0_ →  M1: Depersonalization_T1_ | 0.12 | 0.10 | 1.25 | 1.28 | .2010 |
| a2  M1: Depersonalization_T1_ → M2: MADRS_T0–T1_ | 0.18 | 0.10 | 1.89 | 1.83 | .0666 |
| b  M2: MADRS _T0–T1_ →  Y: MADRS_T0–T6_ | 0.33 | 0.10 | 3.45 | 2.96 | **.0031** |
| c’ (direct) | 0.25 | 0.10 | 2.43 | 2.44 | **.0148** |
| c (total) | 0.39 | 0.09 | 4.25 | 3.53 | **.0004** |
| Indirect Triple Effect  a1·a2·b | 0.01 | 0.01 | 0.89 | 1.59 | .1126 |
| **Derealization manifestations** | | | | | |
| a1  X: EXPECT_T0_ →  M1: Derealization_T1_ | 0.20 | 0.10 | 2.03 | 2.04 | **.0409** |
| a2  M1: Derealization_T1_ → M2: MADRS_T0–T1_ | 0.17 | 0.10 | 1.72 | 1.78 | .0745 |
| b  M2: MADRS_T0–T1_ →  Y: MADRS_T0–T6_ | 0.34 | 0.10 | 3.42 | 3.22 | **.0013** |
| c’ (direct) | 0.24 | 0.11 | 2.24 | 2.29 | **.0217** |
| c (total) | 0.39 | 0.09 | 4.20 | 3.55 | **.0004** |
| Indirect Triple Effect  a1·a2·b | 0.01 | 0.01 | 1.12 | 1.97 | **.0483** |
| **Amnesia manifestations** | | | | | |
| a1  X: EXPECT_T0_ →  M1: Amnesia_T1_ | 0.19 | 0.10 | 1.84 | 1.75 | .0804 |
| a2  M1: Amnesia_T1_ →  M2: MADRS_T0–T1_ | 0.10 | 0.10 | 0.99 | 1.04 | .2982 |
| b  M2: MADRS_T0–T1_→  Y: MADRS_T0–T6_ | 0.36 | 0.10 | 3.77 | 3.20 | **.0014** |
| c’ (direct) | 0.26 | 0.11 | 2.37 | 2.36 | **.0184** |
| c (total) | 0.39 | 0.09 | 4.23 | 3.57 | **.0004** |
| Indirect Triple Effect  a1·a2·b | 0.01 | 0.01 | 0.77 | 1.32 | .1855 |

**Table S10.** Multivariate Cross-Lagged Analysis: Trivariate RI-CLPM of Therapeutic Expectations, Dissociative Symptoms, and Depressive Symptoms

RI-CLPM1: Trivariate RI-CLPM including EXPECT, CADSS total score, and MADRS. n = 100; 108 parameters; 18 equality constraints; MLR estimator; full-information maximum likelihood for missing data. Age, sex, and education were regressed on all random intercepts; baseline MADRS was additionally regressed on RId. w = GORICA weight; ratio = w(H)/w(complement or unconstrained). we0~~wd0 = covariance of within-person deviations at baseline.

**Model fit**

| **Index** | **Standard** |
| --- | --- |
| χ²(200) | 330.79 |
| P | <.001 |
| CFI | 0.91 |
| RMSEA [90% CI] | 0.08 [0.065 ; 0.096] |
| AIC | 13218.85 |
| BIC | 13453.32 |

**Panel A. Within-session directional paths (lag0)**

| **Path ec: EXPECT(T_n_) → CADSS(T_n_)** | | | | | |
| --- | --- | --- | --- | --- | --- |
| **Session** | **β** | **SE** | **z** | **P** | **βstd** |
| T1 | 0.26 | 0.10 | 2.50 | .012 | 0.39 |
| T2 | 0.11 | 0.09 | 1.19 | .233 | 0.16 |
| T3 | 0.31 | 0.09 | 3.65 | <.001 | 0.49 |
| T4 | 0.16 | 0.09 | 1.70 | .089 | 0.30 |
| T5 | 0.02 | 0.07 | 0.32 | .752 | 0.06 |
| T6 | 0.04 | 0.09 | 0.48 | .632 | 0.09 |
| **Path ed: EXPECT(T_n_) → MADRS(T_n_)** | | | | | |
| **Session** | **β** | **SE** | **z** | **P** | **βstd** |
| T1 | -0.06 | 0.06 | -0.94 | .346 | -0.16 |
| T2 | -0.15 | 0.06 | -2.61 | .009 | -0.42 |
| T3 | -0.10 | 0.05 | -2.02 | .044 | -0.32 |
| T4 | -0.11 | 0.05 | -2.34 | .019 | -0.40 |
| T5 | -0.09 | 0.05 | -2.06 | .040 | -0.32 |
| T6 | -0.18 | 0.04 | -4.28 | <.001 | -0.55 |
| **Path cd: CADSS(T_n_) → MADRS(T_n_)** | | | | | |
| **Session** | **β** | **SE** | **z** | **P** | **βstd** |
| T1 | -0.06 | 0.09 | -0.63 | .528 | -0.09 |
| T2 | 0.09 | 0.09 | 1.11 | .269 | 0.17 |
| T3 | -0.13 | 0.08 | -1.62 | .105 | -0.27 |
| T4 | -0.04 | 0.07 | -0.55 | .584 | -0.07 |
| T5 | 0.01 | 0.10 | 0.06 | .950 | 0.01 |
| T6 | 0.09 | 0.10 | 0.86 | .391 | 0.12 |

**Panel B. Inter-session reverse paths**

| **Path de: MADRS(T_n_) → EXPECT(T_n+1_)** | | | | | |
| --- | --- | --- | --- | --- | --- |
| **Transition** | **β** | **SE** | **z** | **P** | **βstd** |
| T0→T1 | 0.14 | 0.22 | 0.62 | .534 | 0.05 |
| T1→T2 | 0.03 | 0.30 | 0.11 | .913 | 0.01 |
| T2→T3 | 0.10 | 0.36 | 0.27 | .786 | 0.03 |
| T3→T4 | 0.43 | 0.50 | 0.87 | .387 | 0.13 |
| T4→T5 | -0.29 | 0.36 | -0.82 | .415 | -0.08 |
| T5→T6 | -0.63 | 0.28 | -2.23 | .025 | -0.19 |
| **Path ce: CADSS(T_n_) → EXPECT(T_n+1_)** | | | | | |
| **Transition** | **β** | **SE** | **z** | **P** | **βstd** |
| T1→T2 | 0.07 | 0.15 | 0.50 | .620 | 0.04 |
| T2→T3 | 0.39 | 0.23 | 1.68 | .093 | 0.22 |
| T3→T4 | 0.29 | 0.18 | 1.61 | .107 | 0.18 |
| T4→T5 | -0.12 | 0.19 | -0.62 | .532 | -0.06 |
| T5→T6 | 0.00 | 0.20 | 0.02 | .987 | 0.00 |

**Panel C. Constrained paths and GORICA results**

***Constrained paths (equality across transitions)***

| **Parameter** | **Direction** | **β** | **SE** | **z** | **P** | **βstd range** |
| --- | --- | --- | --- | --- | --- | --- |
| ar_e | EXPECT(T_n_) → EXPECT(T_n+1_) | 0.78 | 0.06 | 12.57 | <.001 | 0.65–0.81 |
| ar_d | MADRS(T_n_) → MADRS(T_n+1_) | 0.25 | 0.09 | 2.77 | .006 | 0.23–0.26 |
| ar_c | CADSS(T_n_) → CADSS(T_n+1_) | 0.32 | 0.17 | 1.96 | .051 | 0.28–0.39 |
| dc | MADRS(T_n_) → CADSS(T_n+1_) | 0.01 | 0.16 | 0.05 | .961 | 0.00–0.01 |

***GORICA directional hypotheses***

| **Hypothesis** | **Loglik**  **(H)** | **Loglik**  **(complement)** | **Loglik weight** | **Penalty** | **w** | **Ratio** |
| --- | --- | --- | --- | --- | --- | --- |
| \|*β_ed_*\| > \|*β_de_*\| | 13.41 | 15.61 | 0.10 | 9.06 | 0.66 | 1.92× |
| \|*β_ec_*\| > \|*β_ce_*\| | 12.49 | 12.99 | 0.38 | 7.35 | 0.89 | 7.71× |

**Panel D. Between-person effects**

***Random intercept covariances***

| **Pair** | **Estimate** | **SE** | **z** | **P** | **r** |
| --- | --- | --- | --- | --- | --- |
| RIe~~RId | -7.20 | 17.43 | -0.41 | .680 | -0.09 |
| RIe~~RIc | 1.21 | 26.81 | 0.05 | .964 | 0.01 |
| RId~~RIc | -30.27 | 13.91 | -2.18 | .030 | -0.29 |

***Baseline within-person covariance***

| **Pair** | **Estimate** | **SE** | **z** | **P** | **r** |
| --- | --- | --- | --- | --- | --- |
| we0~~wd0 | 39.47 | 15.05 | 2.62 | .009 | 0.45 |

***Residual random intercept variances (after covariates)***

| **Random intercept** | **Variance** | **SE** | **z** | **P** | **Std** |
| --- | --- | --- | --- | --- | --- |
| RIe | 108.47 | 46.66 | 2.33 | .020 | 0.81 |
| RId | 62.78 | 37.01 | 1.70 | .090 | 2.27 |
| RIc | 180.01 | 34.96 | 5.15 | <.001 | 0.92 |

***Covariate effects on random intercepts***

| **Random Intercept** | **Covariate** | **β** | **SE** | **z** | **P** | **βstd** |
| --- | --- | --- | --- | --- | --- | --- |
| RIe | Age | 0.25 | 0.10 | 2.53 | .011 | 0.33 |
| RIe | Sex | -2.26 | 3.33 | -0.68 | .496 | -0.10 |
| RIe | Study | -3.20 | 1.49 | -2.14 | .032 | -0.28 |
| RId | Age | -0.01 | 0.05 | -0.23 | .817 | -0.04 |
| RId | Sex | -2.45 | 2.24 | -1.09 | .274 | -0.23 |
| RId | Study | 1.67 | 0.73 | 2.28 | .022 | 0.32 |
| RId | Baseline MADRS | -0.83 | 0.45 | -1.84 | .065 | -0.88 |
| RIc | Age | 0.19 | 0.10 | 1.85 | .065 | 0.20 |
| RIc | Sex | 6.38 | 3.23 | 1.98 | .048 | 0.22 |
| RIc | Study | -0.52 | 1.57 | -0.33 | .740 | -0.04 |

**Panel E. Within-person residual variances**

| **Variable** | **T0** | **T1** | **T2** | **T3** | **T4** | **T5** | **T6** |
| --- | --- | --- | --- | --- | --- | --- | --- |
| we (EXPECT) | 200.00 | 114.02 | 161.38 | 215.70 | 145.93 | 202.61 | 83.88 |
| wd (MADRS) | 38.30 | 34.09 | 32.54 | 26.03 | 24.05 | 32.84 | 26.09 |
| wc (CADSS) | — | 91.21 | 122.55 | 107.42 | 83.56 | 74.01 | 83.28 |

| **Variable** | **T0** | **T1** | **T2** | **T3** | **T4** | **T5** | **T6** |
| --- | --- | --- | --- | --- | --- | --- | --- |
| we (std) | 1.00 | 0.47 | 0.51 | 0.47 | 0.31 | 0.41 | 0.18 |
| wd (std) | 1.00 | 0.93 | 0.77 | 0.61 | 0.64 | 0.78 | 0.52 |
| wc (std) | — | 0.85 | 0.87 | 0.58 | 0.64 | 0.83 | 0.88 |

**Table S11.** Multivariate Cross-Lagged Analysis: Bivariate RI-CLPM of Therapeutic Expectations and Depressive Symptoms

RI-CLPM2: Bivariate RI-CLPM of EXPECT and MADRS. n = 100; MLR estimator; full-information maximum likelihood. In Model 2, *β_de_* was freely estimated per transition; *ar_e* and *ar_d* were constrained to equality across transitions. Age, sex, and education were regressed on all random intercepts; baseline MADRS was additionally regressed on RId. *w* = GORICA weight; *ratio* = w(H)/w(complement).

**Model fit**

| χ²(108) | 144.99 |
| --- | --- |
| P | .010 |
| CFI | 0.96 |
| RMSEA [90% CI] | 0.06 [0.030 ; 0.082] |
| AIC | 9112.63 |
| BIC | 9250.70 |

**Panel A. Within-session directional paths (lag0)**

| **Path ed: EXPECT(T_n_) → MADRS(T_n_)** | | | | | |
| --- | --- | --- | --- | --- | --- |
| **Session** | **β** | **SE** | **z** | **P** | **βstd** |
| T1 | -0.08 | 0.06 | -1.50 | .134 | -0.22 |
| T2 | -0.14 | 0.05 | -2.84 | .005 | -0.38 |
| T3 | -0.16 | 0.04 | -3.65 | <.001 | -0.49 |
| T4 | -0.13 | 0.05 | -2.97 | .003 | -0.44 |
| T5 | -0.10 | 0.04 | -2.45 | .014 | -0.34 |
| T6 | -0.18 | 0.05 | -4.00 | <.001 | -0.54 |

**Panel B. Inter-session reverse paths**

| **Path de: MADRS(T_n_) → EXPECT(T_n+1_)** | | | | | |
| --- | --- | --- | --- | --- | --- |
| **Transition** | **β** | **SE** | **z** | **P** | **βstd** |
| T0→T1 | 0.12 | 0.20 | 0.61 | .539 | 0.05 |
| T1→T2 | -0.00 | 0.29 | -0.01 | .991 | -0.00 |
| T2→T3 | 0.15 | 0.37 | 0.39 | .694 | 0.05 |
| T3→T4 | 0.15 | 0.39 | 0.39 | .697 | 0.05 |
| T4→T5 | -0.26 | 0.34 | -0.76 | .450 | -0.07 |
| T5→T6 | -0.57 | 0.26 | -2.21 | .027 | -0.17 |

**Panel C. Constrained paths and GORICA results**

***Constrained paths***

| **Parameter** | **Direction** | **β** | **SE** | **z** | **P** |
| --- | --- | --- | --- | --- | --- |
| ar_e | EXPECT(T_n_)→ EXPECT(T_n+1_) | 0.80 | 0.07 | 12.00 | <.001 |
| ar_d | MADRS(T_n_)→ MADRS(T_n+1_) | 0.26 | 0.08 | 3.06 | .002 |

***GORICA directional hypotheses***

| **Hypothesis** | **Loglik**  **(H)** | **Loglik**  **(complement)** | **Loglik weight** | **Penalty** | **Gorica weight (w)** | **Ratio** |
| --- | --- | --- | --- | --- | --- | --- |
| \|*β_ed_*\| > \|*β_de_*\| | 14.94 | 16.42 | 0.19 | 8.75 | 0.85 | 5.57× |

**Panel D. Between-person effects**

***Random intercept covariances***

| **Pair** | **Estimate** | **SE** | **z** | **P** | **r** |
| --- | --- | --- | --- | --- | --- |
| *RIe~~RId* | -9.35 | 17.29 | -0.54 | .589 | -0.11 |

**Baseline within-person covariance**

| **Pair** | **Estimate** | **SE** | **z** | **P** | **r** |
| --- | --- | --- | --- | --- | --- |
| *we0~~wd0* | 39.69 | 14.61 | 2.72 | .007 | 0.47 |

**Table S12.** Multivariate Cross-Lagged Analysis: Bivariate RI-CLPM of Dissociative Symptoms and Depressive Symptoms

RI-CLPM3: Bivariate RI-CLPM of CADSS and MADRS. n = 100; MLR estimator; full-information maximum likelihood. *β_cd_* was freely estimated per transition; *β_dc_*, *ar_c* and *ar_d* were constrained to equality across transitions. Age, sex, and education were regressed on all random intercepts; baseline MADRS was additionally regressed on RId.

**Model fit**

| **Index** | **Standard** |
| --- | --- |
| χ²(98) | 160.65 |
| P | <.001 |
| CFI | 0.92 |
| RMSEA [90% CI] | 0.08 [0.057 ; 0.102] |
| AIC | 8218.18 |
| BIC | 8335.41 |

**Panel A. Within-session directional paths (lag0)**

| **Path cd: CADSS(T_n_) → MADRS(T_n_)** | | | | | |
| --- | --- | --- | --- | --- | --- |
| **Session** | **β** | **SE** | **z** | **P** | **βstd** |
| T1 | -0.06 | 0.08 | -0.73 | .463 | -0.10 |
| T2 | 0.15 | 0.08 | 1.82 | .069 | 0.28 |
| T3 | -0.19 | 0.09 | -2.19 | .028 | -0.40 |
| T4 | -0.07 | 0.07 | -0.99 | .320 | -0.15 |
| T5 | 0.02 | 0.09 | 0.19 | .853 | 0.03 |
| T6 | 0.15 | 0.10 | 1.43 | .154 | 0.26 |

**Panel B. Inter-session reverse paths**

| **Path dc: MADRS(T_n_) → CADSS(T_n+1_)** | | | | |
| --- | --- | --- | --- | --- |
| **β** | **SE** | **z** | **P** | **βstd** |
| 0.08 | 0.14 | 0.60 | .548 | 0.06 |

**Panel C. Constrained paths**

| **Parameter** | **Direction** | **β** | **SE** | **z** | **P** |
| --- | --- | --- | --- | --- | --- |
| ar_d | MADRS(T_n_) → MADRS(T_n+1_) | 0.27 | 0.08 | 3.58 | <.001 |
| ar_c | CADSS(T_n_) → CADSS(T_n+1_) | 0.44 | 0.19 | 2.37 | .018 |

**Panel D. Between-person effects**

*Random intercept covariance*

| **Pair** | **Estimate** | **SE** | **z** | **P** | **r** |
| --- | --- | --- | --- | --- | --- |
| RIc~~RId | -45.46 | 16.86 | -2.70 | .007 | -0.34 |

**All analyses at constant ketamine dosage of 0.5 mg/kg:**

**Table S13.** Linear Mixed Effects Model 1 of Depression

| **Predictor** | **β** | **SE** | **t** | **P** | **95%CI** | |
| --- | --- | --- | --- | --- | --- | --- |
|  |  |  |  |  | **Lower** | **Upper** |
| **Dissociative symptoms - CADSS** | | | | | | |
| Intercept | 0.01 | 0.08 | 0.07 | .9435 | -0.15 | 0.17 |
| EXPECT | -0.36 | 0.05 | -6.64 | **1.09e-10** | -0.47 | -0.25 |
| CADSS | -0.08 | 0.05 | -1.58 | .1146 | -0.18 | 0.02 |
| Time | -0.30 | 0.04 | -7.09 | **6.71e-12** | -0.38 | -0.22 |
| CADSS x EXPECT | 0.04 | 0.05 | 0.74 | .4589 | -0.06 | 0.13 |
| CADSS x Time | -0.02 | 0.04 | -0.35 | .7297 | -0.10 | 0.07 |
| EXPECT x Time | -0.04 | 0.04 | -0.95 | .3448 | -0.12 | 0.04 |
| CADSS x EXPECT x Time | 0.05 | 0.05 | 1.07 | .2832 | -0.04 | 0.14 |
| Age | 0.05 | 0.07 | 0.75 | .4516 | -0.08 | 0.18 |
| Sex | 0.07 | 0.14 | 0.49 | .6248 | -0.20 | 0.33 |
| Study | -0.07 | 0.07 | -1.14 | .2560 | -0.20 | 0.05 |
| **Depersonalization symptoms** | | | | | | |
| Intercept | 0.02 | 0.08 | 0.19 | .8507 | -0.15 | 0.18 |
| EXPECT | -0.36 | 0.05 | -6.59 | **1.47e-10** | -0.46 | -0.25 |
| Depersonalization | -0.10 | 0.05 | -1.92 | .0560 | -0.19 | 0.00 |
| Time | -0.29 | 0.04 | -7.03 | **1.00e-11** | -0.37 | -0.21 |
| Depersonalization x EXPECT | 0.00 | 0.05 | -0.01 | .9901 | -0.09 | 0.09 |
| Depersonalization x Time | -0.02 | 0.04 | -0.36 | .7184 | -0.10 | 0.07 |
| EXPECT x Time | -0.04 | 0.04 | -0.96 | .3384 | -0.12 | 0.04 |
| Depersonalization x EXPECT x Time | 0.04 | 0.04 | 0.87 | .3849 | -0.05 | 0.12 |
| Age | 0.06 | 0.07 | 0.85 | .3949 | -0.07 | 0.19 |
| Sex | 0.06 | 0.14 | 0.44 | .6592 | -0.21 | 0.33 |
| Study | -0.07 | 0.07 | -1.11 | .2675 | -0.20 | 0.06 |
| **Derealization manifestations** | | | | | | |
| Intercept | 0.00 | 0.08 | 0.02 | .9804 | -0.16 | 0.16 |
| EXPECT | -0.36 | 0.05 | -6.69 | **8.41e-11** | -0.46 | -0.25 |
| Derealization | -0.09 | 0.05 | -1.69 | .0923 | -0.19 | 0.01 |
| Time | -0.30 | 0.04 | -7.06 | **8.25e-12** | -0.38 | -0.22 |
| Derealization x EXPECT | 0.05 | 0.05 | 0.94 | .3495 | -0.05 | 0.14 |
| Derealization x Time | -0.03 | 0.04 | -0.57 | .5695 | -0.11 | 0.06 |
| EXPECT x Time | -0.04 | 0.04 | -0.89 | .3743 | -0.12 | 0.05 |
| Derealization x EXPECT x Time | 0.04 | 0.05 | 0.77 | .4428 | -0.06 | 0.13 |
| Age | 0.06 | 0.07 | 0.85 | .3949 | -0.07 | 0.19 |
| Sex | 0.07 | 0.14 | 0.51 | .6093 | -0.20 | 0.34 |
| Study | -0.07 | 0.06 | -1.13 | .2589 | -0.20 | 0.05 |
| **Amnesia manifestations** | | | | | | |
| Intercept | 0.02 | 0.08 | 0.20 | .8437 | -0.15 | 0.18 |
| EXPECT | -0.38 | 0.05 | -7.11 | **6.08e-12** | -0.49 | -0.27 |
| Amnesia | 0.02 | 0.05 | 0.34 | .7342 | -0.08 | 0.11 |
| Time | -0.28 | 0.04 | -6.83 | **3.52e-11** | -0.37 | -0.20 |
| Amnesia x EXPECT | 0.06 | 0.04 | 1.39 | .1660 | -0.03 | 0.15 |
| Amnesia x Time | 0.05 | 0.04 | 1.29 | .1976 | -0.03 | 0.13 |
| EXPECT x Time | -0.03 | 0.04 | -0.76 | .4455 | -0.12 | 0.05 |
| Amnesia x EXPECT x Time | 0.04 | 0.04 | 0.99 | .3228 | -0.04 | 0.11 |
| Age | 0.03 | 0.07 | 0.43 | .6651 | -0.10 | 0.16 |
| Sex | 0.05 | 0.14 | 0.35 | .7230 | -0.22 | 0.32 |
| Study | -0.08 | 0.07 | -1.19 | .2338 | -0.21 | 0.05 |

**LME1** **model fit:** *AIC* = 892.2; *BIC* = 951.3; *LogLik* = -431.1; *R²_marginal_* = 0.29, *R²_conditional_* = 0.59, *RMSE* = 0.54. **Random effects**: *SD(intercept)* = 0.52 [0.42 to 0.65], *SD(time)* = 0.18 [0.09 to 0.34], *Residual(SD)* = 0.62 [0.56 to 0.68].

**LME1_depersonalization_** **model fit***: AIC* = 891.8; *BIC* = 951.0; *LogLik* = -430.9; *R²_marginal_* = 0.29, *R²_conditional_* = 0.59, *RMSE* = 0.54. **Random effects**: *SD(intercept)* = 0.52 [0.42 to 0.65], *SD(time)* = 0.19 [0.10 to 0.34], *Residual(SD)* = 0.62 [0.56 to 0.68].

**LME1_derealization_** **model fit***: AIC* = 892.0; *BIC* = 951.1; *LogLik* = -431.0; *R²_marginal_* = 0.29, *R²_conditional_* = 0.61, *RMSE* = 0.53. **Random effects**: *SD(intercept)* = 0.52 [0.42 to 0.65], *SD(time)* = 0.19 [0.10 to 0.34], *Residual(SD)* = 0.62 [0.56 to 0.68].

**LME1_amnesia_** **model fit***: AIC* = 889.6; *BIC* = 948.8; *LogLik* = -429.8; *R²_marginal_* = 0.29, *R²_conditional_* = 0.61, *RMSE* = 0.53. **Random effects**: *SD(intercept)* = 0.54 [0.44 to 0.67], *SD(time)* = 0.18 [0.10 to 0.34], *Residual(SD)* = 0.61 [0.55 to 0.67].

β correspond to regression coefficients; SE indicates standard error; CI, confidence interval; AIC, Akaike information criterion; BIC, Bayesian information criterion; LogLik, log-likelihood; R²_marginal,_ proportion of variance explained by fixed effects; R²_conditional_, proportion of variance explained by fixed and random effects; RMSE, Root-Mean-Square Error; SD, standard Deviation; z, standardized value; CADSS, Clinician-Administered Dissociative States Scale; MADRS, Montgomery-Åsberg Depression Rating Scale.

**Table S14**. Linear Mixed Effects Model 2 of Depression

| **Predictor** | **β** | **SE** | **t** | **P** | **95%CI** | |
| --- | --- | --- | --- | --- | --- | --- |
|  |  |  |  |  | **Lower** | **Upper** |
| Intercept | 0.02 | 0.08 | 0.20 | .8431 | -0.15 | 0.18 |
| EXPECT | -0.37 | 0.05 | -7.02 | **1.07e-11** | -0.48 | -0.27 |
| Time | -0.29 | 0.04 | -6.77 | **5.11e-11** | -0.37 | -0.20 |
| EXPECT * Time | -0.04 | 0.04 | -0.88 | .3801 | -0.12 | 0.05 |
| Age | 0.04 | 0.07 | 0.65 | .5175 | -0.09 | 0.18 |
| Sex | 0.06 | 0.14 | 0.43 | .6655 | -0.21 | 0.33 |
| Study | -0.08 | 0.07 | -1.24 | .2175 | -0.21 | 0.05 |

**LME2** **model fit***:* *AIC* = 887.8.0, *BIC* = 931.2, *LogLik* = -432.9; *R²_marginal_* = 0.28, *R²_conditional_* = 0.60, *RMSE* = 0.54. **Random effects**: *SD(intercept)* = 0.54 [0.44 to 0.67], *SD(time)* = 0.19 [0.11 to 0.34], *Residual(SD)* = 0.61 [0.56 to 0.67].

**Table S15**. Linear Mixed Effects Model 3 of Depression

| **Predictor** | **β** | | **SE** | | **t** | **P** | **95%CI** | |
| --- | --- | --- | --- | --- | --- | --- | --- | --- |
|  |  |  |  |  |  |  | **Lower** | **Upper** |
| **Dissociative symptoms - Total CADSS score** | | | | | | | | |
| Intercept | -0.01 | | 0.09 | | -0.10 | .9167 | -0.19 | 0.17 |
| CADSS | -0.13 | | 0.05 | | -2.47 | **.0138** | -0.24 | -0.03 |
| Time | -0.29 | | 0.04 | | -6.76 | **5.36e-11** | -0.38 | -0.21 |
| CADSS * Time | 0.00 | | 0.04 | | 0.00 | .9990 | -0.09 | 0.09 |
| Age | -0.01 | | 0.08 | | -0.12 | .9075 | -0.16 | 0.14 |
| Sex | 0.22 | | 0.15 | | 1.44 | .1521 | -0.08 | 0.52 |
| Study | -0.01 | | 0.07 | | -0.15 | .8814 | -0.16 | 0.13 |
| **Depersonalization symptoms** | | | | | | | | |
| Intercept | 0.00 | | 0.09 | | -0.04 | .9653 | -0.19 | 0.18 |
| Depersonalization | -0.15 | | 0.05 | | -2.90 | **.0039** | -0.26 | -0.05 |
| Time | -0.29 | | 0.04 | | -6.87 | **2.76e-11** | -0.37 | -0.20 |
| Depersonalization * Time | -0.02 | | 0.04 | | -0.53 | .5966 | -0.11 | 0.06 |
| Age | -0.01 | | 0.08 | | -0.11 | .9151 | -0.16 | 0.14 |
| Sex | 0.20 | | 0.15 | | 1.30 | .1945 | -0.10 | 0.50 |
| Study | -0.00 | | 0.07 | | -0.05 | .9567 | -0.15 | 0.14 |
| **Derealization manifestations** | | | | | | | | |
| Intercept | -0.01 | | 0.09 | | -0.14 | .8863 | -0.20 | 0.17 |
| Derealization | -0.12 | | 0.05 | | -2.27 | **.0240** | -0.23 | -0.02 |
| Time | -0.30 | | 0.04 | | -6.76 | **5.24e-11** | -0.38 | -0.21 |
| Derealization * Time | 0.00 | | 0.04 | | -0.05 | .9602 | -0.09 | 0.08 |
| Age | -0.01 | | 0.08 | | -0.11 | .9149 | -0.16 | 0.14 |
| Sex | 0.23 | | 0.15 | | 1.49 | .1375 | -0.07 | 0.53 |
| Study | -0.01 | | 0.07 | | -0.17 | .8641 | -0.16 | 0.13 |
| **Amnesia manifestations** | | | | | | | | |
| Intercept | 0.00 | 0.10 | | -0.02 | | .9853 | -0.19 | 0.19 |
| Amnesia | -0.01 | 0.05 | | -0.13 | | .8971 | -0.11 | 0.09 |
| Time | -0.28 | 0.04 | | -6.64 | | **1.11e-10** | -0.36 | -0.20 |
| Amnesia * Time | 0.06 | 0.04 | | 1.43 | | .1547 | -0.02 | 0.14 |
| Age | -0.04 | 0.08 | | -0.45 | | .6499 | -0.19 | 0.12 |
| Sex | 0.21 | 0.16 | | 1.34 | | .1824 | -0.10 | 0.52 |
| Study | -0.01 | 0.08 | | -0.17 | | .8651 | -0.16 | 0.14 |

**LME3** **model fit**: *AIC* = 926.5, *BIC* = 969.9, *LogLik* = -452.3; *R²_marginal_* = 0.13, *R²_conditional_* = 0.56, *RMSE* = 0.56. **Random effects**: *SD(intercept)* = 0.63 [0.52 to 0.77], *SD(time)* = 0.17 [0.09 to 0.34], *Residual(SD)* = 0.64 [0.58 to 0.70].

**LME3_depersonalization_** **model fit:** *AIC* = 924.1, *BIC* = 967.4, *LogLik* = -451.0; *R²_marginal_* = 0.14, *R²_conditional_* = 0.57, *RMSE* = 0.56. **Random-effects**: *SD(intercept)* = 0.64 [0.53 to 0.78], *SD(time)* = 0.15 [0.07 to 0.35], and *Residual(SD)* = 0.64 [0.58 to 0.70].

**LME3_derealization_** **model fit:** *AIC* = 927.6, *BIC* = 971.0, *LogLik* = -452.8; *R²_marginal_* = 0.13, *R²_conditional_* = 0.56, *RMSE* = 0.56. **Random-effects**: *SD(intercept)* = 0.63 [0.51 to 0.77], *SD(time)* = 0.18 [0.09 to 0.35], and *Residual(SD)* = 0.64 [0.58 to 0.70].

**LME3_amnesia_** **model fit:** *AIC* = 930.3, *BIC* = 973.7, *LogLik* = -454.1; *R²_marginal_* = 0.11, *R²_conditional_* = 0.57, *RMSE* = 0.56. **Random-effects**: *SD(intercept)* = 0.66 [0.54 to 0.80], *SD(time)* = 0.16 [0.07 to 0.35], and *Residual(SD)* = 0.64 [0.58 to 0.70].

**Table S16**. Session-Level Regressions of Therapeutic Expectations and Dissociative Symptoms

| **Independant Variable** | **Time** | **β** | **SE** | **P** | **%Δ MADRS** | **R²** |
| --- | --- | --- | --- | --- | --- | --- |
| **Therapeutic expectations** | T0 | 10.19 | 2.90 | **.0007** | 23.21 | 0.18 |
|  | T1 | 11.51 | 2.83 | **9.909e-05** | 26.40 | 0.21 |
|  | T2 | 12.00 | 2.90 | **8.2074e-05** | 27.01 | 0.23 |
|  | T3 | 15.10 | 2.93 | **1.9321e-06** | 33.45 | 0.32 |
|  | T4 | 14.70 | 3.27 | **5.2873e-05** | 26.67 | 0.36 |
|  | T5 | 7.64 | 4.72 | .1165 | 13.05 | 0.18 |
|  | T6 | 9.43 | 4.67 | .0538 | 16.78 | 0.29 |
| **Dissociative symptoms** | T1 | 9.41 | 2.75 | **.0009** | 21.57 | 0.17 |
|  | T2 | 7.67 | 2.90 | .0098 | 17.27 | 0.14 |
|  | T3 | 10.12 | 2.99 | **.0011** | 22.44 | 0.20 |
|  | T4 | 6.07 | 3.85 | .1217 | 11.02 | 0.11 |
|  | T5 | 2.44 | 4.96 | .6272 | 4.16 | 0.11 |
|  | T6 | –0.08 | 4.50 | .9861 | –0.14 | 0.19 |

ΔMADRS% = % improvement per 1 SD increase in EXPECT (*depersonalization*, *derealization*, *amnesia*). Bonferroni (α = .0038) corrections were applied.

**Table S17**. Session-Level Regressions of Dissociative Symptom Dimensions

| **Independant Variable** | **Time** | **β** | **SE** | **P** | **%Δ MADRS** | **R²** |
| --- | --- | --- | --- | --- | --- | --- |
| **Depersonalization**  **symptoms** | T1 | 8.96 | 2.76 | **.0016** | 20.55 | 0.16 |
|  | T2 | 9.77 | 2.83 | **.0009** | 22.01 | 0.19 |
|  | T3 | 12.16 | 2.87 | **.0001** | 26.96 | 0.26 |
|  | T4 | 9.04 | 3.76 | .0206 | 16.41 | 0.17 |
|  | T5 | 2.95 | 4.78 | .5424 | 5.03 | 0.11 |
|  | T6 | 2.10 | 4.50 | .6452 | 3.73 | 0.19 |
| **Derealization**  **symptoms** | T1 | 8.57 | 2.80 | .1497 | 19.65 | 0.15 |
|  | T2 | 6.71 | 2.94 | .0251 | 15.12 | 0.13 |
|  | T3 | 9.09 | 3.07 | .0041 | 20.15 | 0.18 |
|  | T4 | 4.24 | 3.99 | .2945 | 7.69 | 0.09 |
|  | T5 | 6.45 | 4.66 | .1767 | 11.01 | 0.16 |
|  | T6 | 2.22 | 4.51 | .6271 | 3.95 | 0.20 |
| **Amnesia**  **symptoms** | T1 | 4.42 | 2.83 | .1212 | 10.14 | 0.09 |
|  | T2 | 1.10 | 2.93 | .7086 | 2.48 | 0.07 |

| **Independant Variable** | **Time** | **β** | **SE** | **P** | **%Δ MADRS** | **R²** |
| --- | --- | --- | --- | --- | --- | --- |
| **Amnesia**  **symptoms** | T4 | 0.37 | 3.69 | .9213 | 0.67 | 0.06 |
|  | T5 | -10.23 | 4.03 | .0168 | -17.46 | 0.27 |
|  | T6 | -8.42 | 3.94 | .0420 | -14.99 | 0.30 |

MADRS% = % improvement per 1 SD increase in CADSS (*depersonalization*, *derealization*, *amnesia*). Bonferroni (α = .0024) corrections were applied.

**Table S18**. Linear Mixed-Effects Model 4 of Therapeutic Expectations

| **Predictor** | **β** | **SE** | **t** | **P** | **95%CI** | |
| --- | --- | --- | --- | --- | --- | --- |
|  |  |  |  |  | **Lower** | **Upper** |
| Intercept | -0.10 | 0.08 | -1.18 | .2396 | -0.26 | 0.06 |
| Improvement Group | 0.64 | 0.13 | 5.04 | **6.58e-07** | 0.39 | 0.89 |
| Time | -0.05 | 0.04 | -1.27 | .2043 | -0.13 | 0.03 |
| Improvement Group * Time | 0.20 | 0.06 | 3.14 | **.0018** | 0.08 | 0.33 |
| Age | 0.28 | 0.08 | 3.64 | **.0003** | 0.13 | 0.43 |
| Study | -0.16 | 0.08 | -2.13 | **.0333** | -0.31 | -0.01 |
| Sex | 0.09 | 0.08 | 1.17 | .2446 | -0.06 | 0.25 |

**LME4 model fit**: *AIC* = 969.7, *BIC* = 1015.4, *LogLik* = -473.8; *R²_marginal_* = 0.34, *R²_conditional_* = 0.84, *RMSE* = 0.39. **Random effects**: *SD(intercept)* = 0.74 [0.63 to 0.87], *SD(time)* = 0.28 [0.21 to 0.39], *Residual(SD)* = 0.46 [0.42 to 0.50].

**Table S19.** Linear Mixed-Effects Model 5 of Dissociative Symptoms

| **Predictor** | **β** | | **SE** | | | **t** | **P** | **95%CI** | | |
| --- | --- | --- | --- | --- | --- | --- | --- | --- | --- | --- |
|  |  |  |  |  |  |  |  | **Lower** | | **Upper** |
| **Dissociative symptoms** | | | | | | | | | | |
| Intercept | -0.10 | | 0.11 | | | -0.90 | .3711 | -0.32 | | 0.12 |
| Improvement Group | 0.54 | | 0.14 | | | 3.85 | **1.38e-04** | 0.27 | | 0.82 |
| Time | -0.12 | | 0.05 | | | -2.62 | **.0092** | -0.21 | | -0.03 |
| Improvement Group x Time | -0.02 | | 0.07 | | | -0.33 | .7408 | -0.17 | | 0.12 |
| Age | 0.17 | | 0.08 | | | 2.01 | **.0448** | 0.00 | | 0.34 |
| Study | 0.02 | | 0.08 | | | 0.24 | .8112 | -0.14 | | 0.18 |
| Sex | 0.26 | | 0.17 | | | 1.48 | .1384 | -0.08 | | 0.60 |
| **Depersonalization symptoms** | | | | | | | | | | |
| Intercept | -0.06 | | 0.10 | | | -0.59 | .5571 | -0.27 | | 0.14 |
| Improvement Group | 0.64 | | 0.13 | | | 4.80 | **2.34e-06** | 0.38 | | 0.90 |
| Time | -0.10 | | 0.05 | | | -1.93 | .0539 | -0.21 | | 0.00 |
| Improvement Group x Time | 0.04 | | 0.09 | | | 0.47 | .6367 | -0.13 | | 0.21 |
| Age | 0.16 | | 0.08 | | | 1.95 | .0522 | 0.00 | | 0.32 |
| Study | 0.06 | | 0.08 | | | 0.81 | .4162 | -0.09 | | 0.22 |
| Sex | 0.11 | | 0.17 | | | 0.68 | .4983 | -0.21 | | 0.44 |
| **Derealization manifestations** | | | | | | | | | | |
| Intercept | | -0.12 | | 0.11 | -1.06 | | .2905 | | -0.33 | 0.10 |
| Improvement Group | | 0.45 | | 0.14 | 3.27 | | **.0012** | | 0.18 | 0.73 |
| Time | | -0.14 | | 0.04 | -3.33 | | **.0009** | | -0.22 | -0.06 |
| Improvement Group x Time | | -0.04 | | 0.07 | -0.55 | | .5819 | | -0.17 | 0.10 |
| Age | | 0.20 | | 0.08 | 2.33 | | **.0206** | | 0.03 | 0.36 |
| Study | | -0.01 | | 0.08 | -0.13 | | .8931 | | -0.17 | 0.15 |
| Sex | | 0.31 | | 0.17 | 1.80 | | .0735 | | -0.03 | 0.65 |
| **Amnesia manifestations** | | | | | | | | | | |
| Intercept | | -0.01 | | 0.11 | -0.10 | | .9194 | | -0.23 | 0.21 |
| Improvement Group | | 0.12 | | 0.14 | 0.83 | | .4045 | | -0.16 | 0.39 |
| Time | | 0.02 | | 0.04 | 0.45 | | .6556 | | -0.06 | 0.10 |
| Improvement Group x Time | | -0.13 | | 0.07 | -1.96 | | .0505 | | -0.26 | 0.00 |
| Age | | -0.01 | | 0.09 | -0.11 | | .9106 | | -0.18 | 0.16 |
| Study | | -0.01 | | 0.09 | -0.06 | | .9513 | | -0.17 | 0.16 |
| Sex | | 0.12 | | 0.18 | 0.67 | | .5030 | | -0.23 | 0.47 |

**LME5 model fit**: *AIC* = 868.2; *BIC* = 911.3; *LogLik* = -423.1; *R²_marginal_* = 0.18, *R²_conditional_* = 0.80, *RMSE* = 0.42. **Random effects**: *SD(intercept)* = 0.80 [0.68 to 0.95], *SD(time)* = 0.28 [0.15 to 0.50], *Residual(SD)* = 0.51 [0.45 to 0.57].

**LME5_depersonalization_ model fit**: *AIC* = 889.5; *BIC* = 932.6; *LogLik* = -433.7; *R²_marginal_* = 0.23, *R²_conditional_* = 0.79, *RMSE* = 0.42. **Random effects**: *SD(intercept)* = 0.74 [0.62 to 0.87], *SD(time)* = 0.36 [0.25 to 0.53], *Residual(SD)* = 0.52 [0.46 to 0.57].

**LME5_derealization_ model fit**: *AIC* = 874.5; *BIC* = 917.6; *LogLik* = -426.2; *R²_marginal_* = 0.16, *R²_conditional_* = 0.77, *RMSE* = 0.45. **Random effects**: *SD(intercept)* = 0.79 [0.66 to 0.94], *SD(time)* = 0.22 [0.11 to 0.41], *Residual(SD)* = 0.53 [0.48 to 0.59].

**LME5_amnesia_ model fit**: *AIC* = 947.0; *BIC* = 990.1; *LogLik* = -462.5; *R²_marginal_* = 0.02, *R²_conditional_* = 0.65, *RMSE* = 0.54. **Random effects**: *SD(intercept)* = 0.78 [0.65 to 0.92], *SD(time)* = 0.15 [0.05 to 0.44], *Residual(SD)* = 0.62 [0.56 to 0.68].

**Table S20.** Three-Path Mediation Model with Average Therapeutic Expectations, and Dissociative Symptom Scores

| **Path** | **β** | **SE** | **t** | **Z** | **P** |
| --- | --- | --- | --- | --- | --- |
| a1  X: EXPECT_T0_ → M1: CADSS_T1_ | 0.19 | 0.10 | 1.98 | 2.05 | **.0405** |
| a2  M1: CADSS_T1_ →  M2: MADRS_T0-T1_ | 0.20 | 0.10 | 1.92 | 1.92 | .0543 |
| b  M2: MADRS_T0-T1_ →  Y: MADRS_T0-T6_ | 0.39 | 0.09 | 4.31 | 3.37 | **.0008** |
| c’ (direct) | 0.23 | 0.09 | 2.67 | 2.61 | **.0090** |
| c (total) | 0.38 | 0.09 | 4.24 | 3.69 | **.0002** |
| Indirect Triple Effect  a1·a2·b | 0.02 | 0.01 | 1.16 | 2.01 | **.0449** |

Three-paths mediation results testing whether early dissociative symptoms (M1) and early improvement (M2) sequentially mediate the association between therapeutic expectations (X) and end-of-course depression improvement (Y). CADSS, Clinician-Administered Dissociative States Scale; EXPECT, Therapeutic expectations score; MADRS, Montgomery-Åsberg Depression Rating Scale.

**Table S21.** Three-Path Mediation Model with Dissociative Symptom Dimensions

| **Path** | **β** | **SE** | **t** | **Z** | **P** |
| --- | --- | --- | --- | --- | --- |
| **Depersonalization manifestations** | | | | | |
| a1  X: EXPECT_T0_→  M1: Depersonalization_T1_ | 0.10 | 0.10 | 1.04 | 1.10 | .2722 |
| a2  M1: Depersonalization_T1_ →  M2: MADRS_T0-T1_ | 0.18 | 0.10 | 1.82 | 1.84 | .0664 |
| b  M2: MADRS_T0-T1_ →  Y: MADRS_T0-T6_ | 0.39 | 0.09 | 4.43 | 3.40 | **.0007** |
| c’ (direct) | 0.24 | 0.09 | 2.70 | 2.60 | **.0094** |
| c (total) | 0.37 | 0.09 | 4.24 | 3.68 | **.0002** |
| Indirect Triple Effect  a1·a2·b | 0.01 | 0.01 | 0.80 | 1.37 | .1695 |
| **Derealization manifestations** | | | | | |
| a1  X: EXPECT_T0_ →  M1: Derealization_T1_ | 0.19 | 0.10 | 1.90 | 1.84 | .0655 |
| a2  M1: Derealization_T1_ → M2: MADRS_T0-T1_ | 0.17 | 0.10 | 1.71 | 1.76 | .0784 |
| b  M2: MADRS_T0-T1_ →  Y: MADRS_T0-T6_ | 0.39 | 0.09 | 4.28 | 3.37 | **.0007** |
| c’ (direct) | 0.22 | 0.09 | 2.58 | 2.61 | **.0090** |
| c (total) | 0.37 | 0.09 | 4.16 | 3.69 | **.0002** |
| Indirect Triple Effect  a1·a2·b | 0.01 | 0.01 | 1.07 | 1.82 | **.0690** |
| **Amnesia manifestations** | | | | | |
| a1  X: EXPECT_T0_ →  M1: Amnesia_T1_ | 0.20 | 0.10 | 2.00 | 1.94 | .0524 |
| a2  M1: Amnesia_T1_ →  M2: MADRS_T0-T1_ | 0.11 | 0.10 | 1.09 | 1.10 | .2692 |
| b  M2: MADRS_T0-T1_ →  Y: MADRS_T0-T6_ | 0.41 | 0.09 | 4.77 | 3.45 | **.0006** |
| c’ (direct) | 0.25 | 0.09 | 2.80 | 2.84 | **.0045** |
| c (total) | 0.37 | 0.09 | 4.20 | 3.71 | **.0002** |
| Indirect Triple Effect  a1·a2·b | 0.01 | 0.01 | 0.85 | 1.39 | .1656 |

M1: EXPECT(Tn) → CADSS(Tn); M2: CADSS(Tn) → MADRS(Tn+1); M3: EXPECT(Tn) → MADRS(Tn+1). WI, within-person; BW, between-person. *See Methods S11 for full model specification.* CADSS, Clinician-Administered Dissociative States Scale; EXPECT, therapeutic expectations; MADRS, Montgomery-Åsberg Depression Rating Scale.
